# Language comprehension developmental milestones in typically developing children assessed by the new Language Phenotype Assessment (LPA)

**DOI:** 10.1101/2024.12.30.24318918

**Authors:** Andrey Vyshedskiy, Ariella Pevzner, Brigid Mack, Eva Shrayer, Miranda Zea, Sasha Bunner, Nicole Wong, Elena Baskina, Amira Sheikh, Alessandro Tagliavia, Andriane Schmiedel Fucks, Andressa Schmiedel Sanches Santos, Lucas Ernesto Pavoski Poloni, Elielton Fucks, Yudit Bolotovsky, Sung Jin (Sam) Kang

## Abstract

We recently identified three distinct phenotypes of language comprehension in 31,000 autistic individuals ^1^: 1) individuals with the Command Phenotype were limited to comprehension of simple commands; 2) individuals with the Modifier Phenotype demonstrated additional comprehension of color, size, and number modifiers; and 3) individuals with the Syntactic Phenotype added comprehension of spatial prepositions, verb tenses, flexible syntax, possessive pronouns, complex explanations, and fairytales. We hypothesized that typically developing children progress through the same three language comprehension phenotypes and aimed to investigate the typical age at which each phenotype emerged. To evaluate comprehension in young children, we developed a new assessment, the 15-item Language Phenotype Assessment (LPA), which utilizes toy-animal manipulatives to avoid reliance on picture interpretation and includes short instructions to reduce auditory memory load. The LPA was administered to 116 typically-developing children aged 1.5 to 7 years. Results revealed a developmental pattern in line with the three previously described phenotypes: 50% of typically developing children attained the Command Phenotype by 1.5-years, the Modifier Phenotype by 3.0-years, and the Syntactic Phenotype by 3.7-years-of-age. Future research should focus on establishing normative data for the LPA to enable earlier identification of language comprehension challenges, facilitating timely initiation of language interventions.

## Introduction

Language comprehension development is a complex process marked by distinct milestones at various stages of early childhood ^2,3^. In infancy, children begin discerning subtle nuances in speech by distinguishing phonetic sounds. As they enter toddlerhood, comprehension expands to include vocabulary growth and understanding of grammatical structures. Through interactions with caregivers and exposure to a range of linguistic stimuli, children refine their comprehension skills, learning to interpret contextual cues and infer meaning from conversations and narratives. This developmental journey is driven by a dynamic interplay between genetic predispositions and nurture ^4–6^. Both suboptimal genes and language deprivation can hinder language acquisition ^7^. Deleterious genetic variations, for example, are the leading cause of Autism Spectrum Disorder (ASD) and as many as 40% of individuals diagnosed with ASD do not attain the Syntactic Language Comprehension Phenotype ^8^. At the same time, genetically-typical individuals who were not engaged in syntactic conversations in early childhood, also exhibit lifelong deficits in syntactic language comprehension ^7,9–17^. Early language therapy interventions can often mitigate the impact of suboptimal genes and language deprivation ^18–23^. Consequently, there is strong interest in the timely identification of language deficits in young children, to ensure early and effective intervention ^24–27^.

Recent studies have identified three distinct language comprehension phenotypes in autistic individuals. This presents an opportunity to improve the detection and characterization of language deficits in young children ^1^. Analysis of language comprehension abilities of varying complexity in over 31,000 autistic individuals identified three distinct phenotypes that remained consistent across different age groups: 1) individuals in the Command Phenotype were limited to comprehension of simple commands; 2) individuals in the Modifier Phenotype showed additional comprehension of color, size, and number modifiers; and 3) individuals in the Syntactic Phenotype added comprehension of spatial prepositions, verb tenses, flexible syntax, possessive pronouns, complex explanations, and fairytales. The existence of the three distinct language phenotypes was later confirmed in an expanded pool of participants that included other conditions linked to language impairments: mild language delay, apraxia (a motor speech disorder where individuals struggle to plan and coordinate the movements needed for speech despite normal muscle function), Specific Language Impairment, Sensory Processing Disorder, Social Communication Disorder, Down Syndrome, and Attention-Deficit/Hyperactivity Disorder (ADHD) ^28^. We hypothesized that typically developing children progress through these same stages of language comprehension. In this case, establishing normative data for the acquisition of these language comprehension phenotypes could aid in the early diagnosis and monitoring of language development in children.

Accordingly, we reviewed all available assessments that could potentially characterize an individual’s language comprehension phenotype. The ideal instrument would be capable of assessing a range of linguistic abilities with gradually increasing complexity: from the Command level to the Modifier level, and ultimately to the Syntactic level ^1^.

Previous research suggested that typically developing children acquire the Command Phenotype by 2 years and the Syntactic Phenotype by 4 years-of-age ^29,30^. Hence, we focused our search on language comprehension assessments for children aged 2 to 4. To our surprise, most available tools for early language learners, such as the Peabody Picture Vocabulary Test (PPVT-4) ^31^ and the Expressive Vocabulary Test (EVT-2) ^32^, rely exclusively on children’s vocabulary assessment. This approach fails to directly evaluate a language comprehension phenotype, potentially leading to inaccurate language assessments since atypical individuals with any language comprehension phenotype can learn an unlimited number of words ^33,34^.

Four language comprehension assessments were identified that focused on evaluating language comprehension at the sentence level: the Preschool Language Scales (PLS-5) ^35^, the Token Test for children ^36,37^, the Clinical Evaluation of Language Fundamentals (CELF-5) ^38^, and the Test for Reception Of Grammar (TROG) ^39^. All these assessments, however, had significant drawbacks.

The PLS-5 is normed from birth to 8-years-of-age. However, most items targeted at younger children are based on vocabulary comprehension. Sentence level comprehension is assessed by overly complex sentences. For example, the easiest syntactic language comprehension task is Question 39: “Put the Mr. Bear in back of you,” “Put the Mr. Bear in front of me.” This task uses the combination of spatial prepositions (in back of, in front of) with pronouns (you, me) in the same sentence. The spatial prepositions and the pronouns present a significant challenge on their own. Combining the two into a single instruction makes the instruction significantly more difficult than each separately. Sensibly, the PLS-5 manual specifies that Q39 is targeted to 4-and-a-half-year-old children. The next syntactic language question after Q39 is Q46, which tests the understanding of nested sentences and is targeted to 6-year-old children. As a result, PLS-5 is not suited for evaluating language comprehension phenotypes in young children in the age range between 2 and 4 years.

The Token Test is normed 3- to 14-years of age. However, it also suffers from overly complex instructions. The simplest Token Test spatial preposition task instructions are: “Put the red circle on the green square” and “Put the green triangle on the red circle” ^36,37^. The spatial prepositions and modifier domains both present a significant challenge to young children on their own. Combining them in a single sentence makes its comprehension even more daunting. The simplest possible spatial preposition domain instruction should only include a verb, a subject, an object, and a spatial preposition: e.g., “Put the lion on the giraffe.” The simplest possible modifier domain instruction should include a single modifier (size or color) and a noun. The longer-than-necessary instructions of the Token Test make it suboptimal for the assessment of language comprehension phenotypes in young language learners in the age range between 2 and 4 years.

The Clinical Evaluation of Language Fundamentals (CELF-5) ^38^ is normed for students 5 to 21 years of age and the Test for Reception of Grammar (TROG) ^39^ is normed for students 4 to 18+ years of age making them unsuitable for assessment of language comprehension phenotypes in children 2- and 4-years-of-age. Both CELF-5 and TROG have an additional problem of requiring participants to answer by pointing to a correct picture, rather than showing the answer with tangible objects (manipulatives). Children with ASD and attention deficit disorder often fail to interpret picture-answers ^40^.

Thus, none of the existing language comprehension tests were suitable for assessing progression over the Command, Modifier, and Syntactic Phenotypes acquired between 2- and 4-years-of-age. Accordingly, it was necessary to develop a new assessment that could evaluate the trajectory of language comprehension toward the Syntactic Phenotype. In the future, such a tool could enable tracking syntactic language acquisition in individuals with impairments, thereby facilitating a timely language therapy intervention. The resulting 15-item Language Phenotype Assessment (LPA) uses toy-animal manipulatives and brief language instructions to minimize auditory memory demands. The LPA was tested in a convenience sample of 116 typically developing children.

## Methods

The Language Phenotype Assessment (LPA) is rooted in a set of common language comprehension items whereby the participants are required to follow verbal commands of increasing difficulty. The LPA consists of 15 items: three items at the command language levels, five items at the modifier language level, and seven items at the syntactic level (Table 1). All items are scored as either *1:* participant demonstrated an understanding of the item, or *0*: participant did not demonstrate an understanding of the item. The LPA total score was calculated based on the number of items completed correctly. A total score of 15 indicated that a participant demonstrated an understanding of all items. Similarly, a participant who demonstrated an understanding of seven items would receive a total score of 7; a participant who demonstrated an understanding of no items would receive a total score of 0, and so on.

**Table 1.**
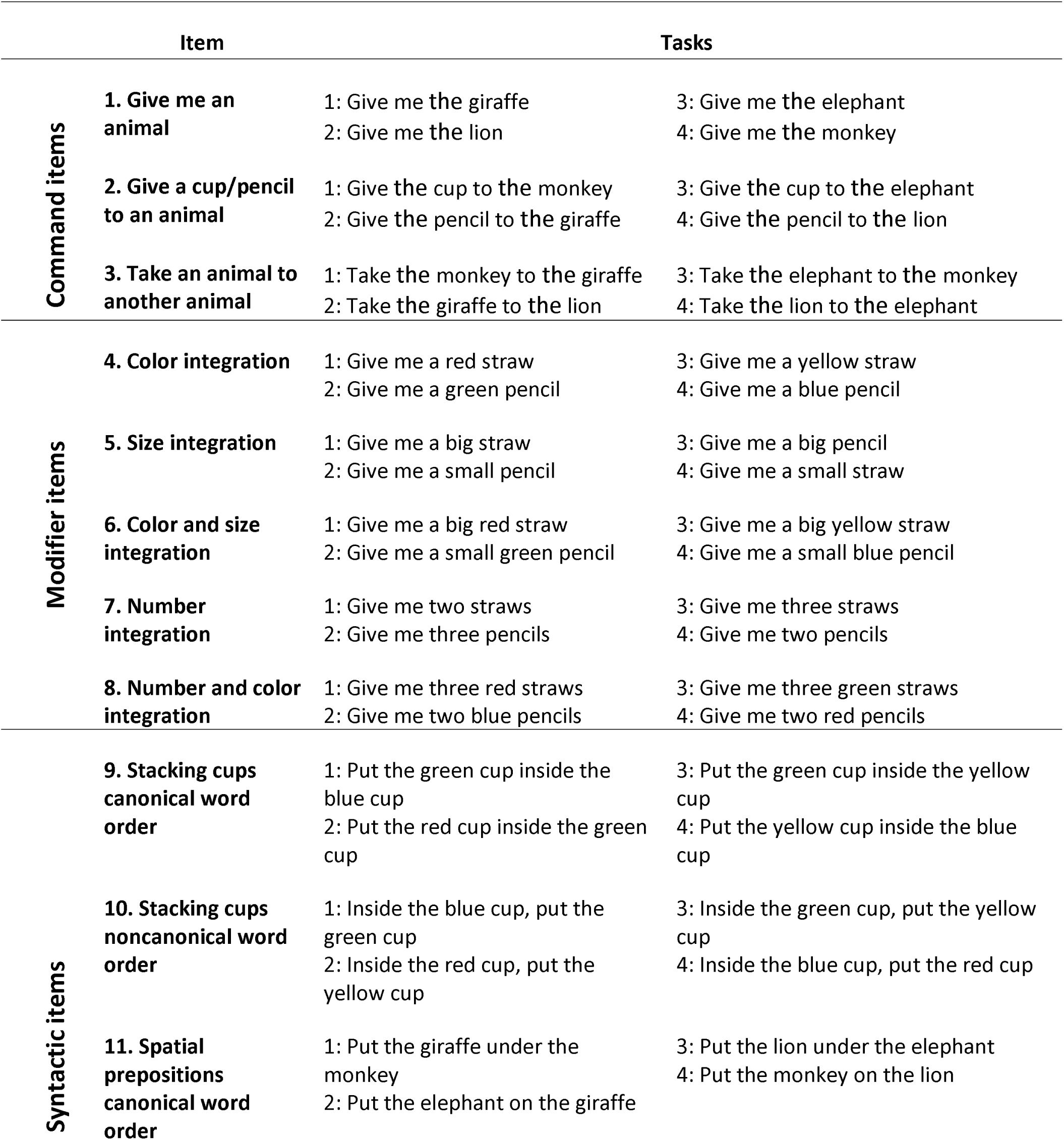
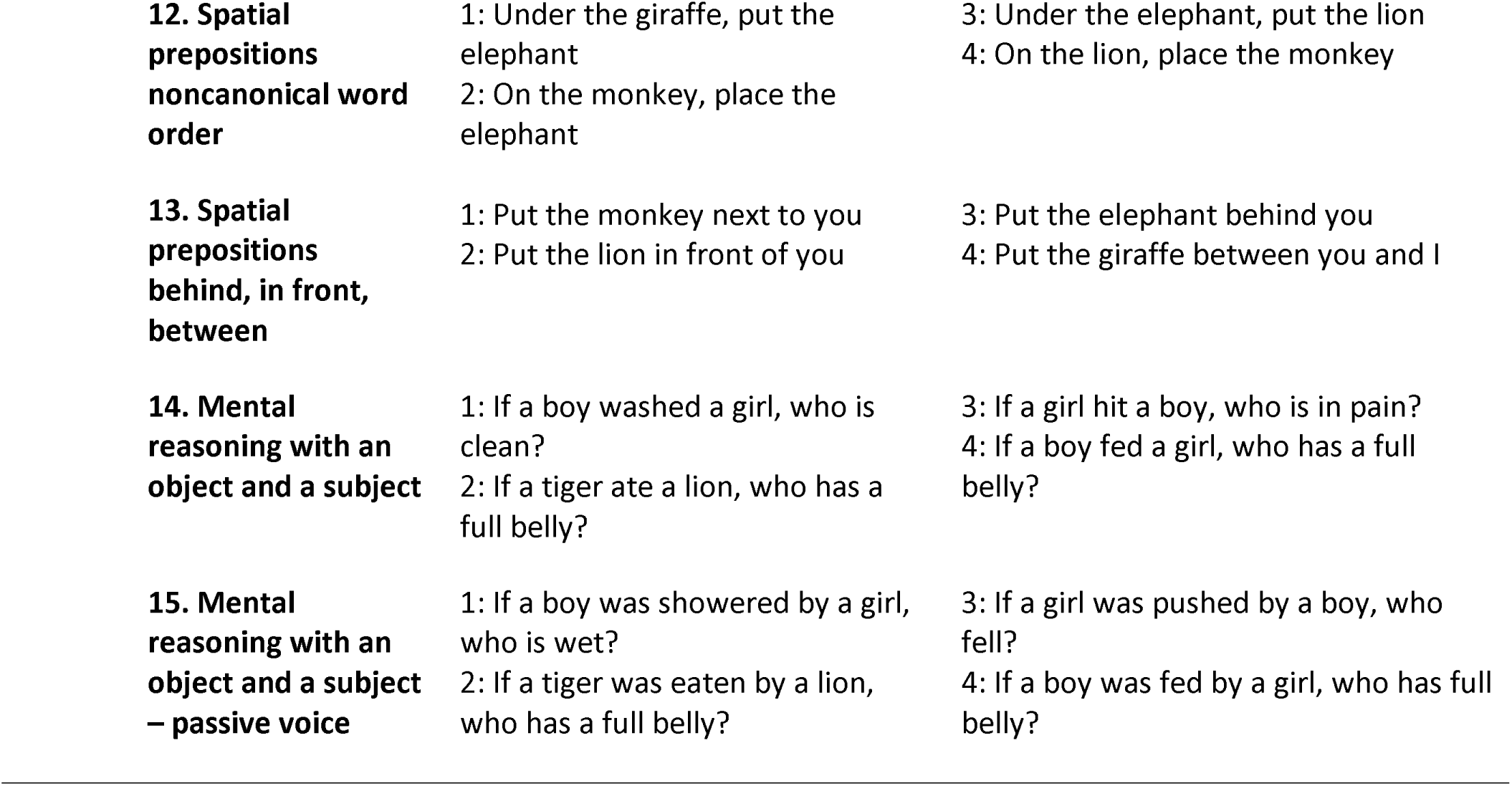
Clinician-observed Language Phenotype Assessment (LPA) items and associated tasks. When a child successfully completes at least three out of the four tasks, the item is scored as 1. The total score is then used to determine the phenotype.

| Item |  | Tasks |  |
| --- | --- | --- | --- |
| Command items | **1. Give me an animal** | 1: Give me the giraffe<br>2: Give me the lion | 3: Give me the elephant<br>4: Give me the monkey |
|  | **2. Give a cup/pencil to an animal** | 1: Give the cup to the monkey<br>2: Give the pencil to the giraffe | 3: Give the cup to the elephant<br>4: Give the pencil to the lion |
|  | **3. Take an animal to another animal** | 1: Take the monkey to the giraffe<br>2: Take the giraffe to the lion | 3: Take the elephant to the monkey<br>4: Take the lion to the elephant |
| Modifier items | **4. Color integration** | 1: Give me a red straw<br>2: Give me a green pencil | 3: Give me a yellow straw<br>4: Give me a blue pencil |
|  | **5. Size integration** | 1: Give me a big straw<br>2: Give me a small pencil | 3: Give me a big pencil<br>4: Give me a small straw |
|  | **6. Color and size integration** | 1: Give me a big red straw<br>2: Give me a small green pencil | 3: Give me a big yellow straw<br>4: Give me a small blue pencil |
|  | **7. Number integration** | 1: Give me two straws<br>2: Give me three pencils | 3: Give me three straws<br>4: Give me two pencils |
|  | **8. Number and color integration** | 1: Give me three red straws<br>2: Give me two blue pencils | 3: Give me three green straws<br>4: Give me two red pencils |
| Syntactic items | **9. Stacking cups canonical word order** | 1: Put the green cup inside the blue cup<br>2: Put the red cup inside the green cup | 3: Put the green cup inside the yellow cup<br>4: Put the yellow cup inside the blue cup |
|  | **10. Stacking cups noncanonical word order** | 1: Inside the blue cup, put the green cup<br>2: Inside the red cup, put the yellow cup | 3: Inside the green cup, put the yellow cup<br>4: Inside the blue cup, put the red cup |
|  | **11. Spatial prepositions canonical word order** | 1: Put the giraffe under the monkey<br>2: Put the elephant on the giraffe | 3: Put the lion under the elephant<br>4: Put the monkey on the lion |

|  |  |  |
| --- | --- | --- |
| **12. Spatial prepositions noncanonical word order** | 1: Under the giraffe, put the elephant<br>2: On the monkey, place the elephant | 3: Under the elephant, put the lion<br>4: On the lion, place the monkey |
| **13. Spatial prepositions behind, in front, between** | 1: Put the monkey next to you<br>2: Put the lion in front of you | 3: Put the elephant behind you<br>4: Put the giraffe between you and I |
| **14. Mental reasoning with an object and a subject** | 1: If a boy washed a girl, who is clean?<br>2: If a tiger ate a lion, who has a full belly? | 3: If a girl hit a boy, who is in pain?<br>4: If a boy fed a girl, who has a full belly? |
| **15. Mental reasoning with an object and a subject – passive voice** | 1: If a boy was showered by a girl, who is wet?<br>2: If a tiger was eaten by a lion, who has a full belly? | 3: If a girl was pushed by a boy, who fell?<br>4: If a boy was fed by a girl, who has full belly? |

The entire test was designed to take approximately 10 minutes to complete. A detailed description of each LPA test item is provided in Table 1. Each item consists of four tasks. To demonstrate their understanding of an item, a child must correctly complete at least three out of the four tasks.

### Manipulatives

The LPA uses the following manipulatives (Figure S1): 1) Large pencils of 4 colors: red, blue, green, and yellow. 2) Small pencils (1/3 length of large pencils) of 4 colors: red, blue, green, and yellow. 3) Large straws of 4 colors: red, blue, green, and yellow (three of each color). 4) Small straws (1/3 length of large straws) of 4 colors: red, blue, green, and yellow (one of each color). 5) A set of puppet-like plush animals: giraffe, lion, elephant, and monkey. 6) A set of four colored cups: red, blue, green, and yellow.

### 1. Command item – give me an animal

For this item, puppet-like plush animals (giraffe, lion, elephant, and monkey, Figure S1) were placed on a flat surface. Each participant was asked to identify the animals to confirm basic knowledge of animal names. If they didn’t know the name, the name was repeated three times. At least 75% accuracy was required to earn a score of *1.* This 75% accuracy threshold was chosen to accommodate possible lapses in attention. With four animals, the probability of answering 75% of tasks correctly by chance is 4.7%.

### 2. Command item – give a cup or a pencil to an animal

Again, the giraffe, lion, elephant, and monkey were laid on a flat surface, each placed as far apart from the others as possible. A cup and a pencil were positioned nearby. Participants were asked to give a cup or a pencil to an animal. After each task, the tester encouraged the child by saying “Good job,” but no feedback was given concerning the correctness of the answer to prevent the child from memorizing the answers. At least 75% accuracy was required to earn a score of *1.* The probability of answering 75% of tasks correctly by chance is 0.68%

### 3. Command item – take an animal to another animal

As before, the giraffe, lion, elephant, and monkey were laid on a flat surface, each placed as far apart from the others as possible. Each participant was asked to bring one animal to another animal. At least 75% accuracy was required to earn a score of *1.* With four animals, the probability of answering 75% of tasks correctly by chance is 1.5%.

### 4. Modifier item – color integration

Integration of color requires the participants to integrate a noun and an adjective. Participants were asked to select an object (e.g., *red* straw) placed among 24 objects (4 large pencils of different colors, 4 small pencils of different colors, 12 large straws of different colors, 4 small straws of different colors), thus forcing the participant to notice and integrate both color and object. Prior to completing this item, participants were asked to point to and name the color of straws and pencils to confirm that they understood the word for specific colors. Participants needed to answer correctly at least 3 out of 4 tasks (75% accuracy) to receive a score of *1* for this item. The probability of answering 75% of tasks correctly by chance is <1.5%.

### 5. Modifier item – size integration

Integration of size requires the participants to integrate a noun and an adjective. Participants were asked to select an object (e.g. *big* straw) placed among 24 objects listed in item 4, thus forcing the participant to notice and integrate size and object. Prior to completing this item, participants were asked to point to and name the size of various objects to confirm that they understood the words *big* and *small*. Participants needed to answer correctly at least 3 out of 4 tasks (75% accuracy) to receive a score of *1* for this item. The probability of answering 75% of tasks correctly by chance is <1.5%.

### 6. Modifier item – color and size integration

Participants were asked to select an object (e.g., *long red* straw) placed among 24 objects listed in item 4, thus forcing the participant to notice and integrate color, size and object. Participants needed to answer correctly at least 3 out of 4 tasks (75% accuracy) to receive a score of *1* for this item. The probability of answering 75% of tasks correctly by chance is 0.68%.

### 7. Modifier item – number integration

Participants were asked to select two or three objects (e.g., *three* straw) placed among 24 objects listed in item 4. Participants needed to answer correctly at least 3 out of 4 tasks (75% accuracy) to receive a score of *1* for this item. The probability of answering 75% of tasks correctly by chance is <1%.

### 8. Modifier item – number and color integration

Participants were asked to select two or three objects of a specific color (e.g., *three red* straws) placed among 24 objects listed in item 4. Participants needed to answer correctly at least 3 out of 4 tasks (75% accuracy) to receive a score of *1* for this item. The probability of answering 75% of tasks correctly by chance is <1%.

### 9. Syntactic item – stacking cups canonical word order

A set of four colored cups (Figure S1) was used for this test. The purpose of this task was to determine whether participants could properly arrange two cups, based on verbal instructions. Before the test, participants were given a demonstration of how to “put the *blue* cup inside the *red* cup” and, if necessary, were helped to stack the cups correctly. This training session with the *blue* and *red* cups was repeated while randomly switching the cup order until the participant was able to stack the correct cups on their own with no errors. Once participants were comfortable stacking the two training cups, they were asked to stack four cups of various color combinations (Table 1, ‘Tasks’ column). Once the cups were stacked, each task was recorded as correct or incorrect. Participants needed to answer correctly at least 3 out of 4 tasks (75% accuracy) to receive a score of *1* for this item. With four cup colors, the probability of answering 75% of tasks correctly by chance is 0.2%.

### 10. Syntactic item – stacking cups noncanonical word order

The directions for stacking cups were varied syntactically from the previous item. E. g., participants were instructed: “inside the blue cup, put the green cup” or “inside the red cup, put the green cup.” This was intended to be more difficult than a canonical instruction such as “put the green cup inside the blue cup.” Participants needed to answer correctly at least 3 out of 4 tasks (75% accuracy) to receive a score of *1* for this item. With four cup colors, the probability of answering 75% of tasks correctly by chance is 0.2%.

### 11. Syntactic item – spatial prepositions canonical word order

In this item, participants were instructed to maneuver the plush animals according to the spatial prepositions *on top of* and *under*. Before the test, participants were given a demonstration of how to “put the monkey *on top of* and *under* the lion.” This training session with the monkey and lion was repeated while randomly switching the order of animals until the participant was able to stack the animals on their own with no errors. Once subjects were comfortable stacking the two training animals, participants were asked to show “the giraffe under the monkey” or “the elephant on top of the giraffe.” The pair containing monkey and lion was not used in the actual test. The spatial prepositions *behind* and *in front of* were not used to avoid confusion about whether the perspective was from the experimenter or the participant. Participants needed to answer correctly at least 3 out of 4 tasks (75% accuracy) to receive a score of 1 for this item. With four animals, the probability of answering 75% of tasks correctly by chance is 0.2%.

### 12. Syntactic item – spatial prepositions noncanonical word order

The directions for spatial prepositions were varied syntactically from the previous item. E.g., participants were instructed: “under the monkey, put the giraffe.” This was intended to be more difficult than a canonical instruction such as “put the giraffe under the monkey.” Identically to all other items, at least 75% accuracy was required to earn a score of *1.* With four animals, the probability of answering 75% of tasks correctly by chance is 0.2%.

### 13. Syntactic item – spatial prepositions next to, behind, in front, between

Participants were instructed in the following way: “Put the monkey next to you,” “Put the lion in front of you,” “Put the elephant behind you,” and “Put the giraffe between you and I.” Identically to all other items, at least 75% accuracy was required to earn a score of *1.* With four animals, the probability of answering 75% of tasks correctly by chance is <0.1%.

### 14. Syntactic item – mental reasoning with an object and a subject

In the final two items, participants were asked to synthesize multiple pieces of information to solve simple mental reasoning tasks. For example, the prompt could be: “If a boy washed a girl, who is clean?” or “If a tiger ate a lion, who has a full belly”? The four tasks are mixed in such a manner that always picking the first object or always picking the second object would result in two correct responses. Identically to all other items, at least 75% accuracy was required to earn a score of *1.* The probability of answering 75% of tasks correctly by chance is 25%.

### 15. Syntactic item – mental reasoning with an object and a subject – passive voice

In the final item, participants were asked to synthesize multiple pieces of information to solve simple mental reasoning tasks given in passive voice. No tangible objects were used as representation. For example, the prompt could be: “if the boy showered the girl, who is wet?” or “if a tiger was eaten by a lion, who has a full belly”? The four tasks are mixed in such a manner that always picking the first object or always picking the second object would result in two correct responses. Identically to all other items, at least 75% accuracy was required to earn a score of *1.* The probability of answering 75% of tasks correctly by chance is 25%.

### Parent-reported Language Phenotype Evaluation Checklist (LPEC)

While children were evaluated with LPA, we asked parents to evaluate their children’s language comprehension abilities using the Language Phenotype Evaluation Checklist (LPEC), shown in Table 2. To enhance compatibility, the LPEC and LPA have the same structure: three command items, five modifier items, and seven syntactic items. The modifier and syntactic items are identical to those found in the validated parent-reported language comprehension tool, Mental Synthesis Evaluation Checklist (MSEC) ^30,41,42^. The command items (1 to 3) are newly introduced. Response options were: ‘very true’ (2 points), ‘somewhat true’ (1 point), and ‘not true’ (0 points). A higher score indicates better language comprehension abilities.

**Table 2.**
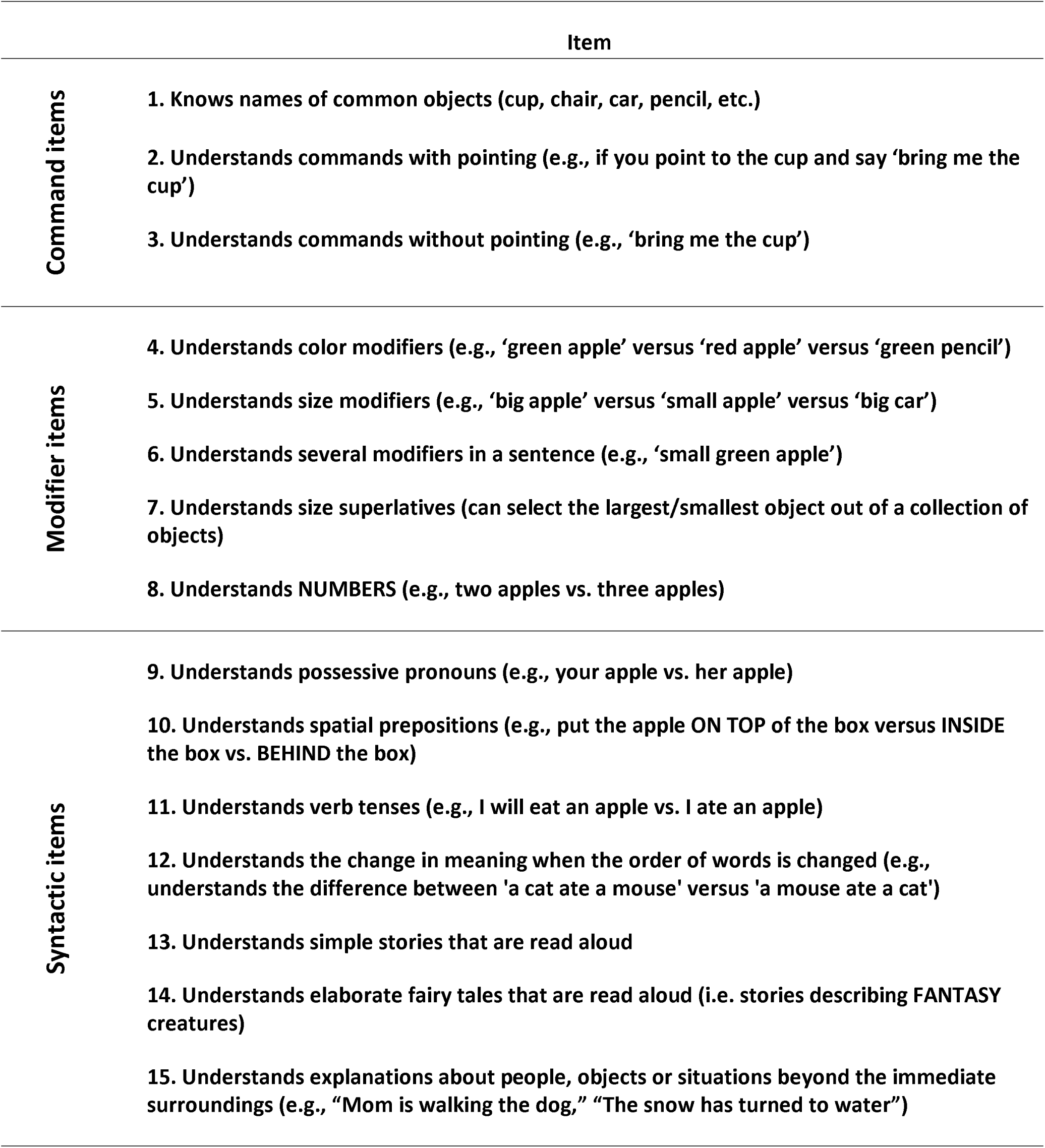
Parent-reported Language Phenotype Evaluation Checklist (LPEC) items. Answer choices were as follows: very true (2 points), somewhat true (1 point), and not true (0 points). A higher score indicates better language comprehension ability.

### Neurotypical participants

Participants for this study were recruited by approaching parents of young children in local parks and inviting them to allow a researcher to administer a language test to their child. This study aimed to capture patterns of language comprehension development patterns in children who do not have language-affecting medical conditions or genetic disorders. The data presented in this manuscript include everyone who agreed to be tested, except two children whose parents indicated that they had a developmental delay. All participants’ caregivers consented to anonymized data analysis and publication of the results. The final sample consisted of 116 neurotypical participants, with a mean age of 4.34 ± 1.4 years (range: 1.5–7 years), of whom 46% were male.

### Participants diagnosed with ASD

Participants with a diagnosis of ASD were attending Soulmare - Autism Clinic, Foz do Iguaçu, Brazil. Participants were recruited by contacting their parents. All children whose parents agreed to participate in the study were included in the study (N=79). After obtaining informed consent from a parent, each child was evaluated by a trained clinician. Participant age range was 2 to 8 years (mean 5.0±1.2 years); 72% were males.

### Statistical analysis

The LPA total score over age and the corresponding percentile curves were generated using Quantile Generalized Additive Models (Quantile GAMs). While traditional linear models assume a constant relationship between variables, Quantile GAMs allow for non-linear modeling that adapt to the data’s structure. This is particularly helpful when modeling developmental data like language acquisition, where growth trajectories often follow non-linear patterns ^43^. By directly estimating specific percentiles (e.g., 5th, 10th, 50th), Quantile GAMs offer insights into how different points in the distribution can shift with age. This approach allowed us to represent growth curves at multiple levels of the distribution, providing a more comprehensive understanding of developmental variation across individuals ^44^. Additionally, by fitting separate curves for each percentile, Quantile GAMs account for skewed distributions and changes in variance over time, both of which are common in developmental data ^45^. Therefore, Quantile GAMs provide a reliable and versatile approach for mapping out age-specific growth patterns without imposing restrictive assumptions, making them highly suitable for percentile-based growth analyses.

Age-related acquisition of each language phenotype and each item was modeled using the sigmoidal function: *100/(1+e((t_c_- t)/τ),* where *t_c_* represents the midpoint of the curve (50%) and *τ* controls the slope. This model was optimized using the R function ‘nlsLM’ (Nonlinear Least-Squares) from the minpack.lm package ^46^.

## Results

### Clinician-observed Language Phenotype Assessment (LPA)

A total of 116 neurotypical participants completed the Language Phenotype Assessment (LPA), Table 1. Each participant was asked to complete 15 items, with each item consisting of four distinct tasks. An item was scored as ‘1’ if at least three out of four tasks were completed correctly, indicating the participant’s understanding; otherwise, it was scored as ‘0.’ Figure 1 illustrates the total LPA score as a function of age, with markers representing individual children’s scores. The near-linear increase in LPA scores between the ages 1.5 and 5 years is followed by a plateau caused by the ceiling effect. Females scored slightly higher than males, although this difference was not statistically significant (Figure S2, Mann-Whitney U test: *p* = 0.58).

**Figure 1.**
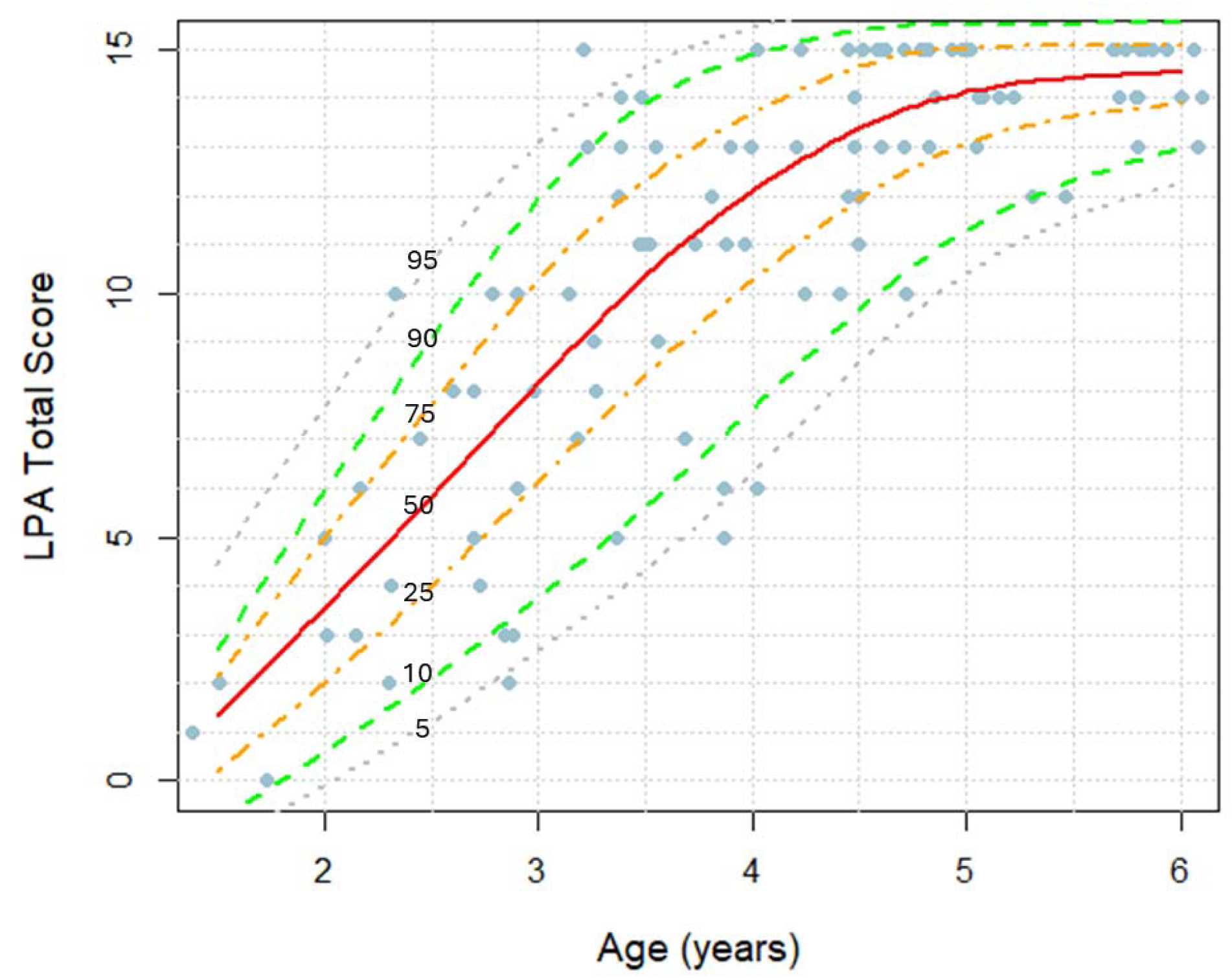
Clinician-observed LPA total score as a function of age in neurotypical children, with percentile lines indicated. Markers represent LPA scores in individual children.

To determine the acquisition age for the Command, Modifier, and Syntactic Phenotypes, we applied the following criteria: 1) the Command Phenotype was assigned if a participant demonstrated understanding of at least two out of three command items (items 1 to 3, Table 1); 2) the Modifier Phenotype was assigned if a participant demonstrated understanding of at least three out of five modifier items (items 4 to 8); and 3) the Syntactic Phenotype was assigned if a participant demonstrated understanding of at least four out of seven syntactic items (items 9 to 15). Figure 2 displays sigmoidal curves illustrating the percentage of children acquiring each phenotype as a function of age. 50% of children attained the Command Phenotype by 1.6 years, the Modifier Phenotype by 3.0 years, and the Syntactic Phenotype by 3.7 years-of-age. All children acquired the Command Phenotype by 3 years, the Modifier Phenotype by 4 years, and the Syntactic Phenotype by 5 years-of-age.

**Figure 2.**
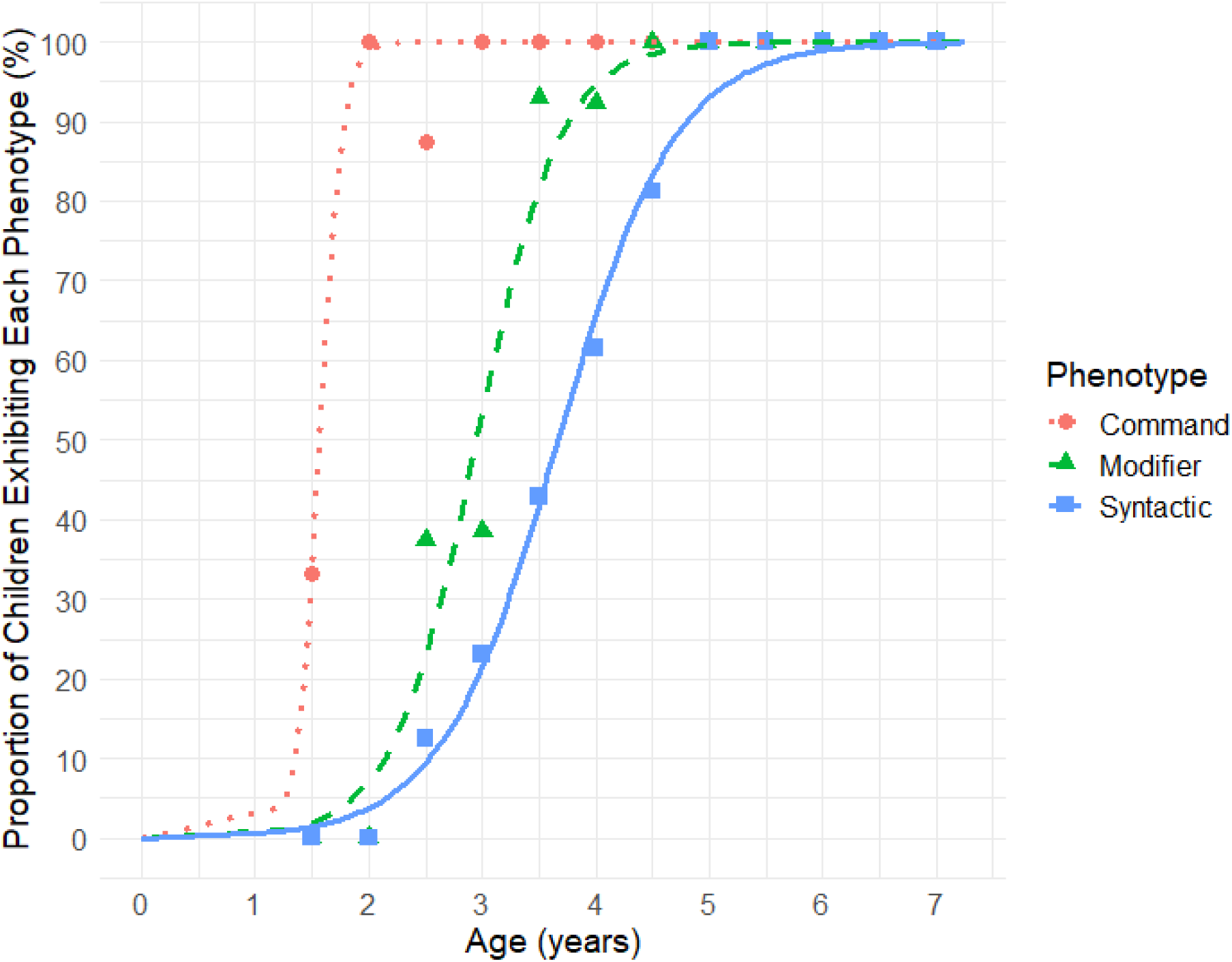
Age-related acquisition of the Command, Modifier, and Syntactic comprehension phenotypes based on clinician-observed Language Phenotype Assessment (LPA) in neurotypical children. Markers represent the proportion of children exhibiting each phenotype, calculated in 0.5-year bins: from 1.25 to 1.75 years, 1.75 to 2.25 years, and so on.

Figures S3 to S18 present sigmoidal curves depicting the percentage of children acquiring each LPA item as a function of age. Table S1 provides a summary of the median age of attainment for each item, offering a concise overview of the developmental milestones median.

### Psychometric Characteristics of LPA

Internal consistency was excellent (Cronbach’s alpha equals to 0.92), suggesting high reliability. The item-total correlations for items 4 to 15 ranged from 0.54 to 0.79, indicating that these items contribute meaningfully to the overall scale. The item-total correlations for items 1 to 3 ranged from 0.24 to 0.45, which is expected since the majority of participants had already attained an understanding of these items. The LPA test-retest reliability was evaluated by calculating a Pearson’s Correlation between the first administration of the LPA and the re-administration of the LPA to the same participants approximately 4 months (average: 111 ± 59 days, range: 29 – 234 days) later (20 participants). The 4-month test-retest correlation coefficient for LPA was *r* = 0.96 (*p* < 0.0001), revealing excellent LPA long-term stability. The inter-observer agreement was very high as indicated by Kappa analysis with Kappa = 0.97 and the percent overall agreement = 98.7%.

### Parent-reported Language Phenotype Evaluation Checklist (LPEC)

In addition to the clinician-administered LPA, we asked parents to evaluate their children’s language comprehension abilities using the Language Phenotype Evaluation Checklist (LPEC), shown in Table 2. To enhance compatibility, the LPEC and LPA share a similar structure: three command items, five modifier items, and seven syntactic items. Figure 3 illustrates total LPEC score by age, with markers representing individual children’s scores. A near-linear increase in LPEC scores between ages 1.5 and 5 is followed by a plateau caused by the ceiling effect. Females scored slightly higher than males, although this difference was not statistically significant (Figure S19, Mann-Whitney U test: *p* = 0.60).

**Figure 3.**
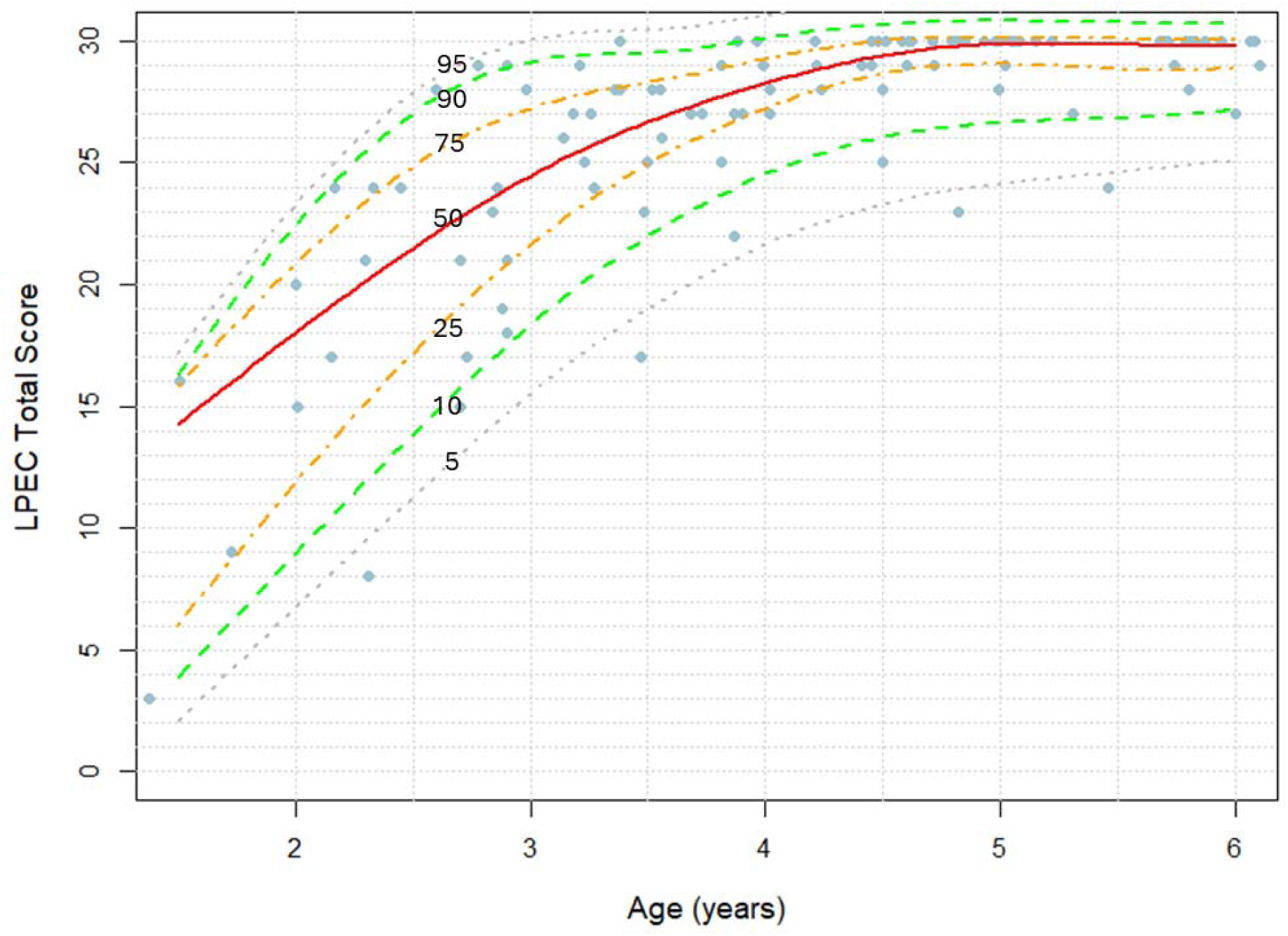
Parent-reported LPEC total score as a function of age in neurotypical children, with percentile lines indicated. Markers represent LPEC scores in individual children.

To evaluate the relationship between clinician-reported LPA scores and parent-reported LPEC scores, Pearson’s correlation analysis was performed. A strong positive correlation was observed (*r* = 0.78, *p* < 0.0001), which demonstrates a significant alignment between the two assessment methods (Figure 4).

**Figure 4.**
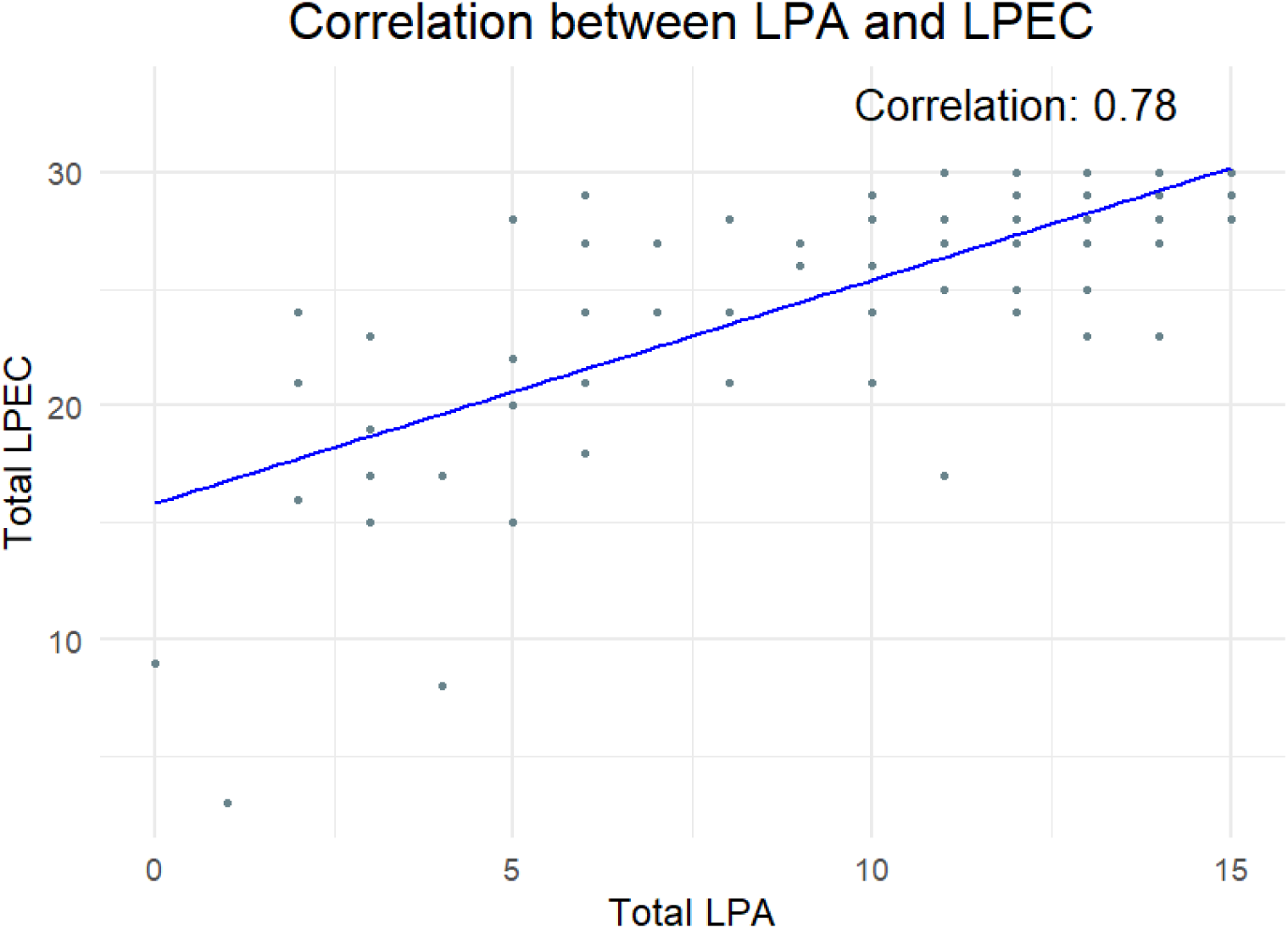
Correlation between clinician-observed LPA score and parent-reported LPEC score in neurotypical children. Each marker represents an individual child score.

To identify the age at which parents perceive their children as having acquired the Command, Modifier, and Syntactic Phenotypes, we applied the same criteria used in the clinician-administered LPA assessment: 1) the Command Phenotype was assigned if a child demonstrated understanding of at least two out of three command items (items 1 to 3, Table 2); 2) the Modifier Phenotype was assigned if a child demonstrated understanding of at least three out of five modifier items (items 4 to 8); and 3) the Syntactic Phenotype was assigned if a child demonstrated understanding of at least four out of seven syntactic items (items 9 to 15). Figure 5 displays sigmoidal curves showing the percentage of children attaining each phenotype. Median acquisition ages reported by parents were 1.6 years for the Command Phenotype, 2.7 years for the Modifier Phenotype, and 3.0 years for the Syntactic Phenotype. According to parent reports, all children acquired the Command Phenotype by age 3, the Modifier Phenotype by age 4, and all but one child achieved the Syntactic Phenotype by age 5.

**Figure 5.**
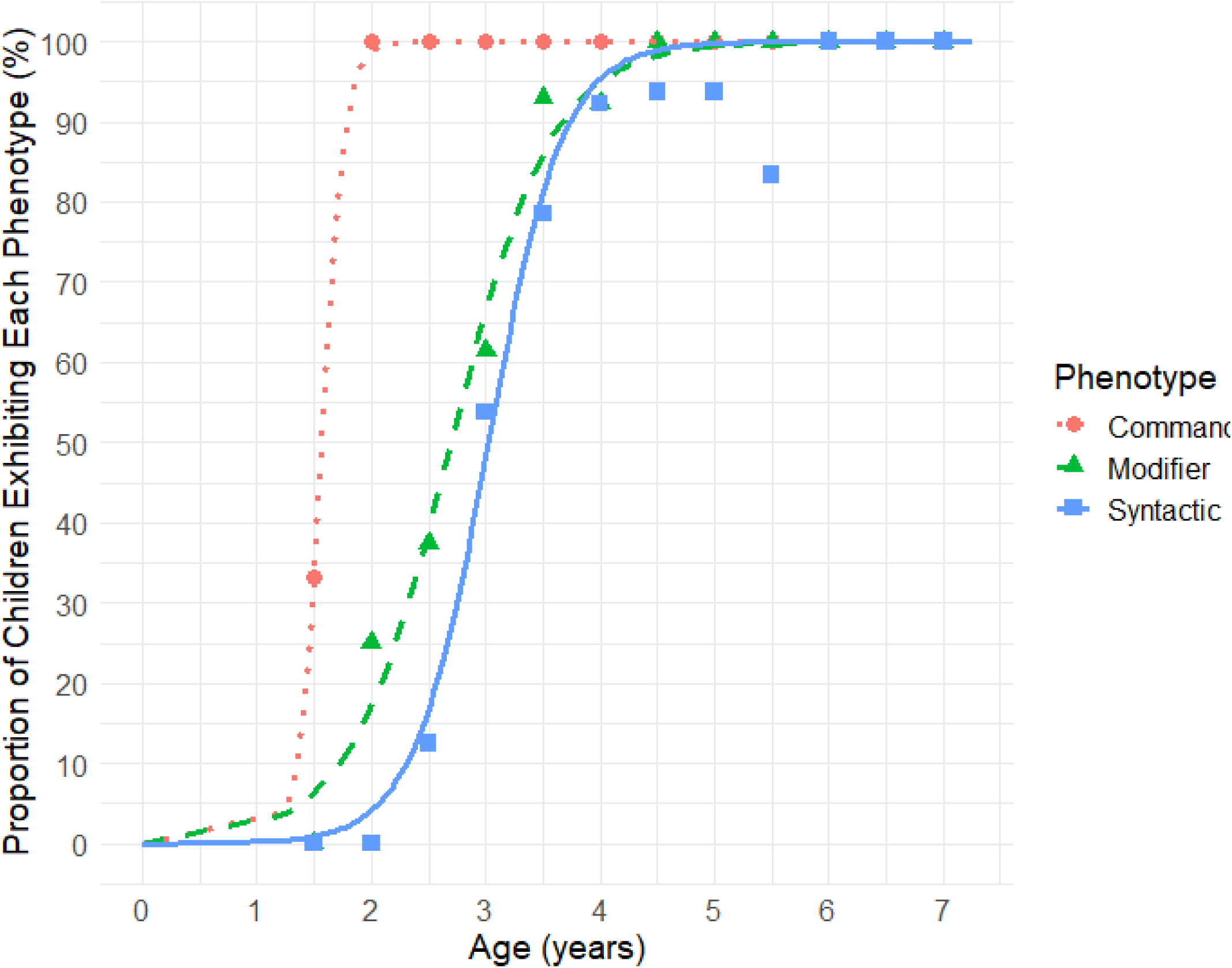
Age-related acquisition of the Command, Modifier, and Syntactic comprehension phenotypes based on parent-reported Language Phenotype Evaluation Checklist (LPEC) neurotypical children. Markers represent the proportion of children exhibiting each phenotype, calculated in 0.5-year bins.

Figures S20 to S34 illustrate sigmoidal curves showing the percentage of children acquiring each LPA item as a function of age. Table S2 summarizes the median age at which each item is acquired according to parents.

### Psychometric Characteristics of parent-reported LPEC

Internal consistency was excellent (Cronbach’s alpha equals to 0.91), suggesting high reliability. The item-total correlations for items 4 to 15 ranged from 0.57 to 0.79, indicating that these items contribute meaningfully to the overall scale. The item-total correlations for items 1 to 3 ranged from 0.29 to 0.50, which is expected since the majority of participants had already attained an understanding of these items. The LPEC test-retest reliability was evaluated by calculating a Pearson’s Correlation between the first administration of the LPEC and the re-administration of the LPEC to the same participants approximately 4 months (average: 111 ± 59 days, range: 29 – 234 days) later (20 participants). The 4-month test-retest correlation coefficient for LPEC was *r* = 0.85 (*p* < 0.0001), revealing excellent LPEC long-term stability.

### LPA generalizability to children diagnosed with Autism Spectrum Disorder (ASD)

A key goal of the LPA is to monitor and optimize language therapy for children diagnosed with ASD. Therefore, it was critical to establish the generalizability of LPA for this population. For this purpose, we administered LPA to 79 children 2 to 8 years of age (mean 5.0 ± 1.2 years) diagnosed with ASD. While all neurotypical children 5 years of age or older attained the Syntactic Phenotype, only 39% of participants diagnosed with ASD reached this milestone (Figure S46). When assessed using the parent-reported LPEC, all but one neurotypical children 5 years of age or older attained the Syntactic Phenotype, compared to only 45% of their peers diagnosed with ASD (Figure S47).

## Discussion

The importance of early language development is well-established ^47–50^. Numerous laws and regulations aim to identify vulnerable individuals, such as those with congenital deafness and ASD, and to provide language therapy as soon as possible. In 1999, the U.S. Congress enacted the “Newborn and Infant Hearing Screening and Intervention Act,” which provides grants to help states establish hearing screening programs for newborns. Otoacoustic Emissions Testing is typically performed at birth, followed by an Auditory Brainstem Response assessment if the initial results suggest potential hearing loss. These screenings enable parents to introduce formal sign language to deaf children soon after birth, thus preventing delays in their exposure to syntactic language. Congenitally deaf children who are exposed to formal sign language early typically exhibit no impairment in their syntactic language comprehension ^51^. This highlights the pressing need to expand these programs to support more children with language comprehension difficulties. A significant challenge in this endeavor is the early identification of vulnerable children.

It is widely recognized that children’s development follows a predetermined time course with a predictable sequence of developmental stages ^52^. However, milestones for language comprehension have not been defined with the granularity required for clinical assessment of young children. This study represents a crucial first step toward establishing developmental norms for syntactic language comprehension. Once these norms have been established, assessing children against these benchmarks could become a part of a standard approach for the early identification of language deficits. Early diagnosis can, in turn, facilitate timely intervention therapy, significantly enhancing outcomes for children at risk.

An additional benefit of this assessment tool is its potential to improve the classification of language developmental stages. Currently, individuals’ communication levels are often categorized as *nonverbal*, *minimally-verbal*, or *verbal*. This one-dimensional framework is insufficient for accurately characterizing communication abilities. For example, a nonverbal individual with a Syntactic Language Comprehension Phenotype may possess normal communication skills, albeit in a nonverbal format. In contrast, a verbal individual with only a Command Language Comprehension Phenotype lacks effective communication abilities across any modality. A two-dimensional classification that considers both verbal abilities and language comprehension levels offers a more nuanced understanding of an individual’s communication skills. The identification of three distinct language comprehension phenotypes—Command, Modifier, and Syntactic ^1,28^—presents an opportunity to refine the characterization and monitoring of language comprehension abilities. However, this refinement necessitates a sensitive and robust assessment instrument.

The conventional approach of assessing language comprehension by focusing solely on a child’s vocabulary fails to accurately capture their language comprehension phenotype. Atypically developing children, for instance, may acquire an extensive vocabulary without ever reaching the Syntactic Phenotype. Moreover, this vocabulary-centric approach may reduce the effectiveness of language therapy by shifting the focus to vocabulary-building rather than on essential syntactic exercises needed for comprehensive language development.

A clinician-administered assessment for the three language comprehension phenotypes will enhance the characterization of language deficits, facilitate early diagnosis of language delay, and improve monitoring of syntactic language acquisition in children receiving language therapy. To address this need, we developed the Language Phenotype Assessment (LPA), a 15-item tool specifically designed to evaluate language comprehension phenotypes in very young children. The LPA was administered to 116 neurotypically developing children aged 1.5 to 7 years. Results showed a linear increase in LPA scores from ages 1.5 to 5 (Figure 1), confirming its sensitivity to language comprehension development in early childhood. The progression of language comprehension phenotypes in typically developing children paralleled that of previously reported progression in autistic individuals ^1^: 50% of children attained the Command Phenotype by 1.6 years, the Modifier Phenotype by 3.0 years, and the Syntactic Phenotype by 3.7 years (Figure 2). All participants who acquired the Modifier Phenotype also acquired the Command Phenotype. Furthermore, 97% of participants who acquired the Syntactic Phenotype, also acquired the Modifier phenotype. The remaining 3% of participants likely experienced lapses in attention while evaluating modifier items.

Alongside the clinician-administered LPA, we invited parents to evaluate their children using the Language Phenotype Evaluation Checklist (LPEC). This 15-item LPEC mirrors the structure of the LPA, with three items assessing command abilities, five items assessing modifier abilities, and seven items assessing syntactic abilities. LPEC scores also demonstrated a linear increase from ages 1.5 to 5 (Figure 3), supporting the effectiveness of the LPEC in tracking language comprehension development in young children. The progression of language comprehension phenotypes observed in typically developing children evaluated by parent-reported LPEC was the following: 50% attained the Command Phenotype by 1.6 years, the Modifier Phenotype by 2.7 years, and the Syntactic Phenotype by 3.0 years (Figure 5). The slight differences observed between the clinician- and parent-reported phenotype trajectories (Figures 2 and 5) are likely due to item variation between the two assessments. Items in the LPA are not designed for parent reporting, nor are LPEC items suited for clinical evaluation, precluding the use of identical items across both tools.

Importantly, the developmental progression of language comprehension—advancing from the Command to the Modifier to the Syntactic Phenotype—is consistent across both clinician-assessments and parent-reports. According to the parent-reported LPEC, all participants who acquired the Modifier Phenotype also acquired the Command Phenotype, and all of the participants who acquired the Syntactic Phenotype, also acquired the Modifier phenotype.

The strong correlation between LPA and LPEC scores (*r* = 0.78, *p* < 0.0001, Figure 4) further suggests that both clinicians and parents can reliably assess a child’s language comprehension phenotype.

### Acquisition of the Syntactic Phenotype by participants diagnosed with ASD according to the clinician-administered LPA

Based on the clinician-administered LPA, all neurotypical children aged 5 years and older achieved the Syntactic Phenotype. In contrast, only 39% of their peers with ASD reached this milestone (Figure S46 and Table S7). Among the remaining autistic participants aged 5 years and older, 37% attained only the Modifier Phenotype, 13% achieved the Command Phenotype, and 11% exhibited the pre-Command Phenotype.

All neurotypical children aged 4 years and older achieved the Modifier Phenotype. In contrast, only 66% of their peers with ASD reached this milestone (Table S8). Among the remaining autistic participants aged 4 years and older, 11% attained only the Command Phenotype, and 23% exhibited the pre-Command Phenotype.

Finally, all neurotypical children aged 3 years and older achieved the Command Phenotype. In contrast, only 74% of their peers with ASD reached this milestone (the remaining 26% exhibited the pre-Command Phenotype).

These observations suggest that the clinician-administered LPA may be useful to diagnose and monitor language comprehension deficits in individuals with ASD.

### Acquisition of the Syntactic Phenotype by participants diagnosed with ASD according to the parent-reported LPEC

A similar pattern was identified by the parent-reported LPEC. While all but one of the neurotypical children aged 5 years and older achieved the Syntactic Phenotype according to parent-reported LPEC, only 48% of their peers with ASD reached this milestone (Figure S47 and Table S7). Among the remaining autistic participants aged 5 years and older, 30% attained only the Modifier Phenotype, 9% achieved the Command Phenotype, and 13% exhibited the pre-Command Phenotype.

According to the parent-reported LPEC, all neurotypical children aged 4 years and older achieved the Modifier Phenotype. In contrast, only 68% of their peers with ASD reached this milestone (Table S8). Among the remaining autistic participants aged 4 years and older, 14% attained only the Command Phenotype, and 18% exhibited the pre-Command Phenotype.

According to the parent-reported LPEC, all neurotypical children aged 3 years and older achieved the Command Phenotype. In contrast, only 81% of their peers with ASD reached this milestone (the remaining 19% exhibited the pre-Command Phenotype).

### Using LPA to monitor success of language therapy

The American Academy of Pediatrics (AAP) recommends universal screening of 18- and 24-month-old children for ASD, and also that individuals *diagnosed* with ASD begin to receive no less than 25 hours per week of treatment within 60 days of identification ^53^. However, language therapy delivery often faces three major challenges: insufficient availability, delayed intervention, and misplaced focus.

First, many children receive too little therapy. Two-thirds of US children on the autism spectrum under the age of 8 fail to get even the AAP-recommended minimum treatment hours ^54^. The problems range from the availability to general funding for early intervention programs ^55–57^. Since the AAP’s 2007 recommendation of universal early screening, there has been a sharp increase in demand for ASD-related services ^58^. However, according to a recent study, most states have reported an enormous shortage of ASD-trained personnel, including behavioral therapists (89%), speech-language pathologists (82%), and occupational therapists (79%) ^58^. In many areas, children receive less than 5 hours per week, falling far short of the recommended care levels ^58^.

Second, therapy often begins too late. Although ASD symptoms typically emerge in early development, the average age of diagnosis is around 4 years ^59^. Current clinical guidelines ^60,61^ highlight diagnosis as a catalyst in the clinical pathway to initiate therapeutic intervention. Applying for an early intervention program and waiting for a therapist’s availability adds several additional months before therapy starts. As a result, most children usually do not begin therapy until 4.5 years of age. The scientific consensus, however, is that earlier interventions lead to a greater impact on developmental outcomes. In a recent study, Whitehouse *et al.* determined the efficacy of such preemptive intervention for ASD beginning during the prodromal period ^23^. In the rater-blinded randomized clinical trial, the investigators compared preemptive intervention with usual care. Using community sampling, the investigators identified 104 one-year-old infants showing early behaviors associated with autism spectrum disorder. Each participant was randomized to receive either a preemptive intervention to go along with usual care or just usual care. The preemptive intervention regimen consisted of a 10-session social communication intervention. Each infant was assessed for a number of metrics at baseline (around age 1 year) and age 3 years. The three-year-old reassessment included 89 participants, 45 of whom were in the preemptive intervention group. The intervention led to a reduction in ASD symptom severity, as well as reduced odds of ASD diagnosis. In the intervention group 6.7% of participants were diagnosed with ASD compared to 20.5% in the usual care group. The number needed to treat to reduce the ASD diagnosis was 7.2 individuals. Other studies have also demonstrated that early intervention, particularly language exercises, significantly improves children’s outcomes ^21,62–65^ and is the greatest tool available to reduce the societal cost of treating ASD.

Third, language therapy for autistic children frequently emphasizes vocabulary acquisition and articulation, which are more straightforward goals. Vocabulary training is 1) quicker to implement, 2) highly valued by parents, 3) intuitive—since typically developing children amass extensive vocabularies before mastering syntactic language, and 4) reinforced by conventional language assessments, such as Peabody Picture Vocabulary Test (PPVT-4) ^31^ and Expressive Vocabulary Test (EVT-2) ^32^. However, this heavy emphasis on vocabulary exercises often diverts attention from crucial syntactic language development. Widespread adoption of the LPA as a benchmark for language therapy success could shift the focus back toward syntactic-language training, allowing more children to reach their full linguistic potential.

### Using LPA to fine-tune language therapy

Previously reported language comprehension phenotype analysis ^1,28^ and the results of this study suggest that the Modifier Phenotype, which includes color, size, and number modifiers, precedes the more complex Syntactic Phenotype. Children find it easier to grasp these simpler modifiers before advancing to syntactic structures such as spatial prepositions, verb tenses, flexible syntax, possessive pronouns, and complex explanations. Moreover, mastery of the Modifier Phenotype may be the crucial steppingstone for acquiring the Syntactic Phenotype.

For optimal language development, a structured educational sequence tailored to each child’s current abilities is essential, especially for psychodiverse learners ^66^. Language therapy programs can use the LPA score to guide curriculum design. Children scoring below 2 on the LPA would likely benefit most from command language exercises, while those scoring between 2 and 6 would be best served by focusing on modifier language exercises (Table S6). This finely tuned, stepwise approach can maximize language comprehension progress and readiness for more advanced syntactic training.

### Canonical versus noncanonical word order

Interpretation of any syntactic structure can be routinized. Consider the instruction, “put the green cup inside the blue cup.” This task can be completed through a simple algorithm: (1) lift the first-mentioned cup, and (2) place it into the second-mentioned cup. This type of stepwise response is an example of automatic, routinized action—similar to stopping at a red light (Movie 1). However, routinized responses do not equate to the Syntactic Language Comprehension Phenotype, as they fail to generalize across the wide variety of sentence structures encountered in everyday language. In essence, reducing syntactic interpretation to an algorithmic routine does not translate to one’s ability to understand complex narratives, such as stories and novels, where flexible and context-dependent comprehension is essential.

**Movie 1: https://youtu.be/Hh7pkZB4ETU. Reprinted with permission from ^67^. Authors have obtained written parental consent to publish the video. Most autistic participants (18 to 21 years of age) were able to complete the canonical stacking cups task (e.g., “put the red cup inside the green cup”), but were unable to complete the same task under the condition of non-canonical word order (“inside the green cup, put the red cup”). Failing participants usually selected the correct cups, but assembled them randomly. Most autistic participants have received over 15 years of intensive language therapy and it is likely that their stacking cups routine has been automated through frequent training using canonical word order only.**

Naturally, in designing a test for language comprehension, we aimed to minimize opportunities for participants to rely on rote, algorithmic responses. Ideally, if we had prior knowledge of the specific tasks each individual had been trained on, we could have excluded those tasks. However, in a standardized test, it is impractical to avoid every task for which a participant might have a memorized solution. An alternative approach is to increase item complexity; the more intricate the items, the less likely participants will have been explicitly trained on those sentence structures, reducing reliance on pre-learned algorithms. Yet, we also needed to avoid overly complex grammar that could overwhelm young children’s attention or working memory. Our chosen solution was to use noncanonical word order, creating a balance between discouraging rote responses and maintaining accessibility for all test-takers.

The results of this study corroborate our earlier findings that typically developing children aged 4 years and older comprehend canonical and noncanonical instructions equally well ^29^. This study examined two sets of noncanonical instructions: stacking cups (item 10, Table 1) and spatial prepositions (item 12). Among neurotypical children who successfully followed the stacking cups instruction presented in canonical word order (item 9: “put the red cup inside the green cup”), 92% understood the instruction given in noncanonical word order (item 10: “inside the green cup, put the red cup”). Similarly, among neurotypical participants who followed the spatial prepositions instruction in canonical order (item 11: “put the giraffe under the monkey”), 88% understood the noncanonical version (item 12: “under the giraffe, put the elephant”). These findings indicate that, for the majority of typically developing children aged four and older, there is minimal difference in understanding between canonical and noncanonical word orders.

In contrast, among children aged 4 years and older diagnosed with ASD who successfully followed the canonical stacking cups instruction, only 76% comprehended the noncanonical version. Similarly, among those who successfully followed the canonical “spatial prepositions” instruction, only 60% comprehended the instruction given in noncanonical word order. These asymmetric results are similar to our previous research, which found that nearly half of autistic participants aged 18 to 21 years who successfully followed instructions in canonical word order failed the noncanonical version, suggesting a greater reliance on routinized responses (Movie 1).

### The Modifier Phenotype

Learning words for colors, sizes, and numbers does not equate to acquiring the Modifier Phenotype. Research has demonstrated that chimpanzees and some other animals can learn and even name chips of various colors ^68–70^ and remember Arabic numbers up to nine ^71–74^. However, no non-human animal has shown the ability to combine modifiers—such as color, size, and number—with different nouns ^75^. When faced with tasks that include distractors varying in colors, number, and shapes, such as items 4 to 8 in Table 1, animals consistently perform at chance levels, despite being familiar with individual words. In other words, the Modifier Phenotype is uniquely human. Remarkably, the median age of attainment of this uniquely human phenotype is 3.0 years of age.

## Limitations

Learning words for colors and sizes alone does not equate to the acquisition of the Modifier Phenotype; however, without this foundational vocabulary, it becomes challenging to assess a child’s ability to integrate modifiers with nouns. Therefore, the LPA includes specific steps designed to demonstrate the meanings of individual words to a child prior to testing. Children who are unfamiliar with certain terms typically grasp their meanings quickly: each object used in the LPA is explicitly named, along with the relevant colors, sizes, and numbers. Additionally, spatial prepositions are demonstrated and explained beforehand to ensure that the assessment focuses on language comprehension rather than vocabulary knowledge.

The use of manipulatives in the LPA poses a logistical challenge for clinicians, as picture-based tests are generally easier to store and transport. However, picture-based assessments can present additional difficulties for individuals with attentional issues. For instance, we previously described a case study of Peter, a 7-year-and-7-month-old fully verbal child with ADHD, whose performance on paper-based tests differed markedly from his performance with physical toys ^29^. Peter received a standardized score of 74 on the Fluid Reasoning Index of the WPPSI-IV, placing him below 96% of the population, with scores of 6 on Matrix Reasoning and 5 on Picture Concepts. This IQ test required him to select the picture that best represented the correct answer. To further explore this discrepancy, we tested Peter with our proprietary paper-based assessment. He demonstrated an understanding of matrix analogies by successfully completing simpler items that involved “finding the same objects” and “integrating color, size, and number modifiers.” However, when asked to point to a picture representing “the man ate the whale” or “the whale ate the man,” Peter answered randomly. His performance on the paper-based test was below chance level, but his accuracy jumped to 100% when allowed to respond using physical objects. This suggests that the manipulatives were more engaging for Peter than paper-based options. A follow-up visit four months later, during which Peter had been taking 30 mg of Ritalin daily, yielded different results: he answered all paper-based items correctly. Since the goal of the LPA is to assess language comprehension rather than attention, we aimed to minimize the attentional component. Drawing from our experience with Peter and various studies indicating that manipulatives enhance attention more effectively than pictures ^40,76,77^, we developed the LPA to rely solely on manipulatives, completely eliminating pictures from the assessment.

## Conclusions

We present a 15-item Language Phenotype Assessment (LPA), a 10-minute test designed specifically for evaluating syntactic language comprehension skills in children aged 2 to 5 years. The LPA items incorporate elements such as colors, sizes, numbers, spatial prepositions, and non-canonical syntax, posing a set of novel questions that participants have not encountered before. The total score for the LPA ranges from 0 to 15. The internal consistency of the LPA is high, with a Cronbach’s alpha of 0.92, indicating reliable measurement. Additionally, the assessment demonstrates excellent test-retest reliability and very high inter-observer agreement. Because the LPA does not depend on productive language skills, it serves as a particularly valuable tool for assessing language comprehension development in minimally verbal-children. Using the LPA, we tracked the progression of language comprehension phenotype acquisition—Command, Modifier, and Syntactic—and identified the typical ages at which these are acquired. Assessing children against these benchmarks could become a part of a standard approach for early identification of language deficits.

## Acknowledgments

We wish to thank all participants and their parents who found time to complete assessments.

## Author contributions

AV designed the study. AP, BM, ES, MZ, SB, NW, EB, AS, AT, ASF, ASSS, LEPP, EF, YB collected data. SJK and AV analyzed the data. AV wrote the paper. All authors edited the paper.

## Informed Consent

Caregivers have provided informed consent to anonymized data analysis and publication of the results. The study was conducted in compliance with the Declaration of Helsinki ^78^.

## Compliance with Ethical Standards

The study adhered to all local regulations. Using the Department of Health and Human Services regulations found at 45 CFR 46.101(b)(4), the Biomedical Research Alliance of New York LLC Institutional Review Board (IRB) determined that this research project is exempt from IRB oversight. In Brazil, the study received IRB approval from Centro Universitário Dinâmica das Cataratas (Foz do Iguaçu, Brazil).

## Data Availability

De-identified raw data from this manuscript are available from the corresponding author upon reasonable request.

## Code availability statement

Code is available from the corresponding author upon reasonable request.

## References

1. Vyshedskiy, A., Venkatesh, R. & Khokhlovich, E. Are there distinct levels of language comprehension in autistic individuals – cluster analysis. Npj Ment. Health Res. 3, (2024).

2. Visser-Bochane, M. I., Reijneveld, S. A., Krijnen, W. P., Van der Schans, C. P. & Luinge, M. R. Identifying milestones in language development for young children ages 1 to 6 years. Acad. Pediatr. 20, 421–429 (2020).

3. Luinge, M. R., Post, W. J., Wit, H. P. & Goorhuis-Brouwer, S. M. The Ordering of Milestones in Language Development for Children From 1 to 6 Years of Age. J. Speech Lang. Hear. Res. 49, 923–940 (2006).

4. Dale, P. et al. Genetic influence on language delay in two-year-old children. Nat. Neurosci. 1, 324–328 (1998).

5. Newbury, D. F., Fisher, S. E. & Monaco, A. P. Recent advances in the genetics of language impairment. Genome Med. 2, 6 (2010).

6. Romeo, R. R. et al. Language exposure relates to structural neural connectivity in childhood. J. Neurosci. 38, 7870–7877 (2018).

7. Grimshaw, G. M., Adelstein, A., Bryden, M. P. & MacKinnon, G. E. First-language acquisition in adolescence: Evidence for a critical period for verbal language development. Brain Lang. 63, 237–255 (1998).

8. Fombonne, E. Epidemiological surveys of autism and other pervasive developmental disorders: an update. J. Autism Dev. Disord. 33, 365–382 (2003).

9. Curtiss, S. Abnormal language acquisition and the modularity of language. Linguist. Camb. Surv. 2, 96–116 (1988).

10. Curtiss, S. The case of Chelsea: The effects of late age at exposure to language on language performance and evidence for the modularity of language and mind. UCLA Work. Pap. Linguist. 18, 115–146 (2014).

11. Fromkin, V., Krashen, S., Curtiss, S., Rigler, D. & Rigler, M. The development of language in genie: a case of language acquisition beyond the “critical period”. Brain Lang. 1, 81–107 (1974).

12. Hyde, D. C. et al. Spatial and numerical abilities without a complete natural language. Neuropsychologia 49, 924–936 (2011).

13. Lenneberg, E. H. The biological foundations of language. Hosp. Pract. 2, 59–67 (1967).

14. Morford, J. P. Grammatical development in adolescent first-language learners. Linguistics 41, 681–722 (2003).

15. Morford, J. P. & Hänel-Faulhaber, B. Homesigners as late learners: connecting the dots from delayed acquisition in childhood to sign language processing in adulthood. Lang. Linguist. Compass 5, 525–537 (2011).

16. Ramírez, N. F., Lieberman, A. M. & Mayberry, R. I. The initial stages of first-language acquisition begun in adolescence: when late looks early. J. Child Lang. 40, 391–414 (2013).

17. Vyshedskiy, A., Mahapatra, S. & Dunn, R. Linguistically deprived children: meta-analysis of published research underlines the importance of early syntactic language use for normal brain development. Res. Ideas Outcomes (2017) doi:10.3897/rio.3.e20696.

18. Dawson, G. et al. Randomized, controlled trial of an intervention for toddlers with autism: the Early Start Denver Model. Pediatrics 125, e17–e23 (2010).

19. French, L. & Kennedy, E. M. M. Annual Research Review: Early intervention for infants and young children with, or at-risk of, autism spectrum disorder: a systematic review. J. Child Psychol. Psychiatry 59, 444–456 (2018).

20. Pickles, A. et al. Parent-mediated social communication therapy for young children with autism (PACT): long-term follow-up of a randomised controlled trial. The Lancet 388, 2501–2509 (2016).

21. Rogers, S. J. et al. Autism treatment in the first year of life: a pilot study of infant start, a parent-implemented intervention for symptomatic infants. J. Autism Dev. Disord. 44, 2981–2995 (2014).

22. Shire, S. Y. et al. Hybrid implementation model of community-partnered early intervention for toddlers with autism: A randomized trial. J. Child Psychol. Psychiatry 58, 612–622 (2017).

23. Whitehouse, A. J. O. et al. Effect of Preemptive Intervention on Developmental Outcomes Among Infants Showing Early Signs of Autism: A Randomized Clinical Trial of Outcomes to Diagnosis. JAMA Pediatr. e213298 (2021) doi:10.1001/jamapediatrics.2021.3298.

24. Foster-Cohen, S. H. & Van Bysterveldt, A. K. Assessing the communication development of children with language delay through parent multi-questionnaire reporting. Speech Lang. Hear. 19, 79–86 (2016).

25. Frizelle, P., Thompson, P., Duta, M. & Bishop, D. V. M. Assessing Children’s Understanding of Complex Syntax: A Comparison of Two Methods. Lang. Learn. 69, 255–291 (2019).

26. Jullien, S. Screening for language and speech delay in children under five years. BMC Pediatr. 21, 362 (2021).

27. Sachse, S. & Von Suchodoletz, W. Early identification of language delay by direct language assessment or parent report? J. Dev. Behav. Pediatr. 29, 34–41 (2008).

28. Vyshedskiy, A., Venkatesh, R., Khokhlovich, E. & Satik, D. Three mechanisms of language comprehension are revealed through cluster analysis of individuals with language deficits. Npj Sci. Learn. 9, 1–12 (2024).

29. Vyshedskiy, A. et al. Novel Linguistic Evaluation of Prefrontal Synthesis (LEPS) test measures prefrontal synthesis acquisition in neurotypical children and predicts high-functioning versus low-functioning class assignment in individuals with autism. Appl. Neuropsychol. Child (2020) 10.1080/21622965.2020.1758700.

30. Arnold, M. & Vyshedskiy, A. Combinatorial language parent-report score differs significantly between typically developing children and those with Autism Spectrum Disorders. J. Autism Dev. Disord. (2022) doi:10.1007/s10803-022-05769-8.

31. Dunn, L. M. & Dunn, D. M. PPVT-4: Peabody Picture Vocabulary Test. (Pearson Assessments, 2007).

32. Williams, K. T. Expressive vocabulary test second edition (EVT^TM^ 2). J Am Acad Child Adolesc Psychiatry 42, 864–872 (1997).

33. Fromkin, V., Krashen, S., Curtiss, S., Rigler, D. & Rigler, M. The development of language in genie: a case of language acquisition beyond the “critical period”. Brain Lang. 1, 81–107 (1974).

34. Ramírez, N. F., Lieberman, A. M. & Mayberry, R. I. The initial stages of first-language acquisition begun in adolescence: when late looks early. J. Child Lang. 40, 391–414 (2013).

35. Zimmerman, I. L., Steiner, V. G. & Pond, R. E. PLS-5: Preschool language scale-5 [measurement instrument]. San Antonio TX Psychol. Corp. (2011).

36. De Renzi, A. & Vignolo, L. A. Token test: A sensitive test to detect receptive disturbances in aphasics. Brain J. Neurol. (1962).

37. De Renzi, E. & Faglioni, P. Normative data and screening power of a shortened version of the Token Test. Cortex 14, 41–49 (1978).

38. Wiig, E. H., Secord, W. A. & Semel, E. Clinical Evaluation of Language Fundamentals: CELF-5. (Pearson, 2013).

39. Bishop, D. V. M. Test for reception of grammar—electronic. Lond. Psychol. Corp. (2005).

40. Gomez, M. A., Skiba, R. M. & Snow, J. C. Graspable objects grab attention more than images do. Psychol. Sci. 29, 206–218 (2018).

41. Braverman, J., Dunn, R. & Vyshedskiy, A. Development of the Mental Synthesis Evaluation Checklist (MSEC): A Parent-Report Tool for Mental Synthesis Ability Assessment in Children with Language Delay. Children 5, 62 (2018).

42. Netson, R. et al. A Comparison of Parent Reports, the Mental Synthesis Evaluation Checklist (MSEC) and the Autism Treatment Evaluation Checklist (ATEC), with the Childhood Autism Rating Scale (CARS). Pediatr. Rep. 16, 174–189 (2024).

43. Wood, S. N. Generalized Additive Models: An Introduction with R. (chapman and hall/CRC, 2017).

44. Koenker, R. Quantile regression. Camb. Univ Pr (2005).

45. Yee, T. W. Vector Generalized Linear and Additive Models: With an Implementation in R. (Springer New York, New York, NY, 2015). doi:10.1007/978-1-4939-2818-7.

46. Elzhov, T. V., Mullen, K. M., Spiess, A. & Bolker, B. R interface to the Levenberg-Marquardt nonlinear least-squares algorithm found in MINPACK. Plus Support Bounds 1–2 (2010).

47. Eldevik, S. et al. Using participant data to extend the evidence base for intensive behavioral intervention for children with autism. Am. J. Intellect. Dev. Disabil. 115, 381–405 (2010).

48. Law, J. & Levickis, P. Early language development must be a public health priority. J. Health Visit. 6, 586–589 (2018).

49. Peters-Scheffer, N., Didden, R., Korzilius, H. & Sturmey, P. A meta-analytic study on the effectiveness of comprehensive ABA-based early intervention programs for children with autism spectrum disorders. Res. Autism Spectr. Disord. 5, 60–69 (2011).

50. Virués-Ortega, J. Applied behavior analytic intervention for autism in early childhood: Meta-analysis, meta-regression and dose–response meta-analysis of multiple outcomes. Clin. Psychol. Rev. 30, 387–399 (2010).

51. Mayberry, R. I. Cognitive development in deaf children: The interface of language and perception in neuropsychology. Handb. Neuropsychol. 8, 71–107 (2002).

52. Piaget, J. The stages of the intellectual development of the child. Educ. Psychol. Context Read. Future Teach. 63, 98–106 (1965).

53. Maglione, M. A. et al. Nonmedical interventions for children with ASD: Recommended guidelines and further research needs. Pediatrics 130, S169–S178 (2012).

54. Hume, K., Bellini, S. & Pratt, C. The usage and perceived outcomes of early intervention and early childhood programs for young children with autism spectrum disorder. Top. Early Child. Spec. Educ. 25, 195–207 (2005).

55. Johnson, E. & Hastings, R. P. Facilitating factors and barriers to the implementation of intensive home-based behavioural intervention for young children with autism. Child Care Health Dev. 28, 123–129 (2002).

56. Bibby, P., Eikeseth, S., Martin, N. T., Mudford, O. C. & Reeves, D. Progress and outcomes for children with autism receiving parent-managed intensive interventions. Res. Dev. Disabil. 23, 81–104 (2002).

57. Jacobson, J. W. Early intensive behavioral intervention: Emergence of a consumer-driven service model. Behav. Anal. 23, 149 (2000).

58. Wise, M. D., Little, A. A., Holliman, J. B., Wise, P. H. & Wang, C. J. Can state early intervention programs meet the increased demand of children suspected of having autism spectrum disorders? J. Dev. Behav. Pediatr. 31, 469–476 (2010).

59. van’t Hof, M., et al. Age at autism spectrum disorder diagnosis: A systematic review and meta-analysis from 2012 to 2019. Autism 25, 862–873 (2021).

60. Hyman, S. L., Levy, S. E. & Myers, S. M. Identification, evaluation, and management of children with autism spectrum disorder. Pediatrics 145, (2020).

61. Sandbank, M., Bottema-Beutel, K. & Woynaroski, T. Intervention recommendations for children with autism in light of a changing evidence base. JAMA Pediatr. (2020).

62. Tamis-LeMonda, C. S., Bornstein, M. H. & Baumwell, L. Maternal responsiveness and children’s achievement of language milestones. Child Dev. 72, 748–767 (2001).

63. Siller, M. & Sigman, M. The behaviors of parents of children with autism predict the subsequent development of their children’s communication. J. Autism Dev. Disord. 32, 77–89 (2002).

64. Wan, M. W. et al. Quality of interaction between at-risk infants and caregiver at 12–15 months is associated with 3-year autism outcome. J. Child Psychol. Psychiatry 54, 763–771 (2013).

65. Wetherby, A. M. et al. Parent-implemented social intervention for toddlers with autism: An RCT. Pediatrics 134, 1084–1093 (2014).

66. Vyshedskiy, A. & Dunn, R. Mental Imagery Therapy for Autism (MITA)-An Early Intervention Computerized Brain Training Program for Children with ASD. Autism Open Access 5, 2 (2015).

67. Vyshedskiy, A. Imagination in Autism: A Chance to Improve Early Language Therapy. Healthcare 9, 63 (2021).

68. Matsuzawa, T. Colour naming and classification in a chimpanzee (Pan troglodytes). J. Hum. Evol. 14, 283–291 (1985).

69. Matsuno, T., Kawai, N. & Matsuzawa, T. Color Recognition in Chimpanzees (Pan troglodytes). in Cognitive Development in Chimpanzees (eds. Matsuzawa, T., Tomonaga, M. & Tanaka, M.) 317–329 (Springer-Verlag, Tokyo, 2006). doi:10.1007/4-431-30248-4_20.

70. Pepperberg, I. M. Cognitive and communicative abilities of Grey parrots. Appl. Anim. Behav. Sci. 100, 77–86 (2006).

71. Matsuzawa, T. Use of numbers by a chimpanzee. Nature 315, 57–59 (1985).

72. Biro, D. & Matsuzawa, T. Use of numerical symbols by the chimpanzee (Pan troglodytes): Cardinals, ordinals, and the introduction of zero. Anim. Cogn. 4, 193–199 (2001).

73. Dooley, G. B. & Gill, T. V. Acquisition and use of mathematical skills by a linguistic chimpanzee. in Language learning by a chimpanzee 247–260 (Elsevier, 1977).

74. Pepperberg, I. M. & Carey, S. Grey parrot number acquisition: The inference of cardinal value from ordinal position on the numeral list. Cognition 125, 219–232 (2012).

75. Ape Language. in Why Chimpanzees Can’t Learn Language and Only Humans Can 33–74 (Columbia University Press, 2019). doi:10.7312/terr17110-004.

76. Lin, Y.-H. & Chiang, H.-M. Language comprehension of children with Asperger’s disorder and children with autistic disorder. Res. Autism Spectr. Disord. 8, 767–774 (2014).

77. Maljaars, J., Noens, I., Scholte, E. & van Berckelaer-Onnes, I. Language in low-functioning children with autistic disorder: Differences between receptive and expressive skills and concurrent predictors of language. J. Autism Dev. Disord. 42, 2181–2191 (2012).

78. World Medical Association. World Medical Association Declaration of Helsinki: ethical principles for medical research involving human subjects. JAMA 310, 2191–2194 (2013).

