## Supplemental material for "Language comprehension developmental milestones in typically developing children assessed by the new Language Phenotype Assessment (LPA)"

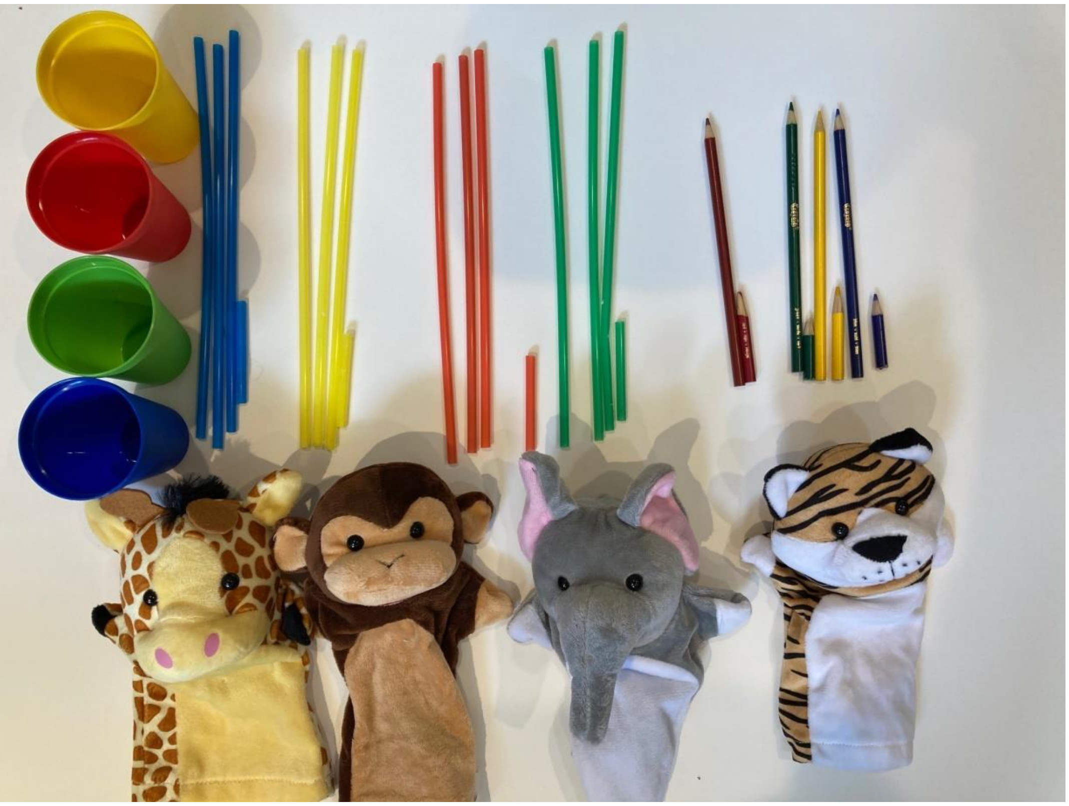

**Figure S1. Manipulatives used by Language Phenotype Assessment (LPA).**

**The Straws and Pencils Set Used in Items 4 to 8:**
-Large pencils of 4 colors: red, blue, green, yellow.
-Small pencils (1/3 length of large pencils) of 4 colors: red, blue, green, yellow.
-Large straws of 4 colors: red, blue, green, yellow (three of each color).
-Small straws (1/3 length of large straws) of 4 colors: red, blue, green, yellow (one of each color).

### Language Phenotype Assessment (LPA) Form

Developed by Andrey Vyshedskiy, Ph.D, Boston University and ImagiRation.com

LPA consists of 15 items. For each item, enter the score of 1 if an examinee demonstrated an understanding of the instructions by successfully completing at least three out of four item's tasks. Otherwise, enter a score of 0. To calculate the total LPA score, add the scores for each item. Highest possible score is 15. Video tutorial: https://youtu.be/SU_6cZEVyTo

Name of Child:_______________ _______________ ☐ Female Age: _________

Last First Date of Birth: _________

Form completed by: _________________ ☐ Male Today's Date:_________

Language spoken at home: _________________

✓/×

**1. Command language – give me an animal (demonstration first)**

| |
|---|
| 1: Give me the giraffe |
| 2: Give me the lion |
| 3: Give me the elephant |
| 4: Give me the monkey |
| Score (0/1): |

**2. Command language – give a cup or a pencil to an animal (demonstration first)**

| |
|---|
| 1: Give the cup to the monkey |
| 2: Give the pencil to the giraffe |
| *3:* Give the cup to the elephant |
| 4: Give the pencil to the lion |
| Score (0/1): |

**3. Command language – take an animal to another animal (no demonstration)**

| |
|---|
| 1: Take the monkey to the giraffe |
| 2: Take the giraffe to the lion |
| 3: Take the elephant to the monkey |
| 4: Take the lion to the elephant |
| Score (0/1): |

**4. Modifier Language – color integration (demonstrate colors)**

| |
|---|
| 1: Give me a *red* straw |
| 2: Give me a *green* pencil |
| 3: Give me a *yellow* straw |
| 4: Give me a *blue* pencil |
| Score (0/1): |

**5. Modifier Language – size integration (demonstrate sizes)**

| |
|---|
| 1: Give me a *big* straw |
| 2: Give me a *small* pencil |
| 3: Give me a *big* pencil |
| 4: Give me a *small* straw |
| Score (0/1): |

**6. Modifier Language – color and size integration (no demonstration)**

| |
|---|
| 1: Give me a *big red* straw |
| 2: Give me a *small green* pencil |
| 3: Give me a *big yellow* straw |
| 4: Give me a *small blue* pencil |
| Score (0/1): |

**7. Modifier Language – number integration (demonstrate numbers two and three)**

| |
|---|
| 1: Give me two straws |
| 2: Give me three pencils |
| 3: Give me three straws |
| 4: Give me two pencils |
| Score (0/1): |

**8. Modifier Language – number and color integration (no demonstration)**

| |
|---|
| 1: Give me tree red straws |
| 2: Give me two blue pencils |
| 3: Give me three green straws |
| 4: Give me two red pencils |
| Score (0/1): |

**9. Syntactic Language – stacking cups canonical word order (show an example: blue cup inside red cup)**

| |
|---|
| 1: Put the *green* cup inside the *blue* cup |
| 2: Put the *red* cup inside the *green* cup |
| *3:* Put the *green* inside the *yellow* cup |
| 4: Put the *yellow* inside the *blue* cup |
| Score (0/1): |

**10. Syntactic Language – stacking cups noncanonical word order (no demo)**

| |
|---|
| 1: Inside the *blue* cup, put the *green* cup |
| 2: Inside the *red* cup, put the *yellow* cup |
| 3: Inside the *green* cup, put the *yellow* cup |
| 4: Inside the *blue* cup, put the *red* cup |
| Score (0/1): |

**11. Syntactic Language – spatial prepositions canonical word order (demonstrate *under* and *on* prepositions: Put monkey on/under elephant)**

| |
|---|
| 1: Put the *giraffe* under the *monkey* |
| 2: Put the *elephant* on the *giraffe* |
| 3: Put the *lion* under the *elephant* |
| 4: Put the *monkey* on the *lion* |
| Score (0/1): |

**12. Syntactic Language – spatial prepositions noncanonical word order (no demo)**

| |
|---|
| 1: Under the *giraffe*, put the *elephant* |
| 2: On the *monkey*, put the *elephant* |
| 3: Under the *elephant*, put the *lion* |
| 4: On the *lion*, put the *monkey* |
| Score (0/1): |

**13. Syntactic Language – behind, in front (no demo)**

| |
|---|
| 1: Put the *monkey* next to you |
| 2: Put the *lion* in front of you |
| 3: Put the *elephant* behind you |
| 4: Put the *giraffe* between you and I |
| Score (0/1): |

**14. Syntactic Language – mental reasoning with an object and a subject (no demo)**

| |
|---|
| 1: If a boy washed a girl, who is clean? (CORRECT=girl) |
| 2: If a tiger ate a lion, who has a full belly? (CORRECT=tiger) |
| 3: If a girl hit a boy, who is in pain? (CORRECT=boy) |
| 4: If a girl defeated a boy, who is the winner? (CORRECT=girl) |
| Score (0/1): |

**15. Syntactic Language – mental reasoning with an object and a subject – passive voice (no demo)**

| |
|---|
| 1: If a boy was showered by a girl, who is wet? (CORRECT=boy) |
| 2: If a tiger was eaten by a lion, who has a full belly? (CORRECT=lion) |
| 3: If a girl was pushed by a boy, who fell? (CORRECT=girl) |
| 4: If a girl was defeated by a boy, who is the winner? (CORRECT=boy) |
| Score (0/1): |

| |
|---|
| **TOTAL SCORE (0-15):** |

| Total Score | Language Comprehension Phenotype |
|---|---|
| 0 | No comprehension |
| 1 | Single word comprehension |
| 2 | Pre-command language comprehension |
| 3 - 5 | Command language comprehension phenotype |
| 6 - 11 | Modifier language comprehension phenotype |
| 12-15 | Syntactic language comprehension phenotype |

### General instructions

- LPA is applicable to children 2 to 5 years of age and older children with language delay.
- The evaluation should be conducted in the child's most familiar language (usually the language spoken at home).
- Children can be accompanied by parents/caregivers in order to enhance their comfort and ensure their collaboration. Parents/caregivers should not provide any correct/incorrect feedback to the child.
- You can repeat each instruction several times to ensure the participant's understanding of the instruction.

### Items instructions

Item 1: Before the test, ask the participant to name the animals. Make sure your subject can select all animals after you name them.

Item 2: This item does not require a demonstration. Place animals away from each other, e.g., into four corners of a small table.

Item 3: This item does not require a demonstration. Place animals away from each other, e.g., into four corners of a small table.

Item 4: Before the test, confirm that the participant knows color names of pencils and straws.

Item 5: Before the test, confirm that the participant knows words for size.

Item 6: This item does not require a demonstration.

Item 7: Before the test, confirm that the participant knows numbers two and three.

Item 8: This item does not require a demonstration.

Item 9: Before the test, ask the participant to identify the color of each of the four cups. For two colors and two colors only (e.g., blue and red), demonstrate what it looks like to put one cup inside of the other. Ask the child to repeat after you. Do not use this color combination (blue and red) in the following tasks.

Item 10: This item does not require a demonstration.

Item 11: Before the test, demonstrate to the participant one example of using "on" and one example of using "under." For example, show them a lion *on* a giraffe and say "this is what it looks like when a lion is *on* a giraffe." Then show them a lion *under* a giraffe and say "this is what it looks like when a lion is *under* a giraffe." During the test avoid the combination of animals used in the example (i.e. lion and giraffe).

Item 12, 13: This item does not require a demonstration.

Items 14 and 15: Instruct the participant that they will not be using any physical objects for these tasks, but that it will all be in their heads. In these items, participants must determine the correct answer by using their own mental representations of objects, without the use of physical objects. Nonverbal children can respond by typing the answer or by any other suitable means.

### LPA Scoring

For each task, note correct or incorrect responses. Accuracy of 75% or more equals the item score of 1, while anything below that equals the score of 0.

Calculate the sum of all items. The highest possible score is 15. A score of 15 indicates that the participant successfully completed each task with at least 75% accuracy and thus received the highest possible score. Similarly, a participant with successful completion of seven items would receive a score of 7, a participant with successful completion of no items would receive a score of 0, and so on.

=========End of the LPA form=========

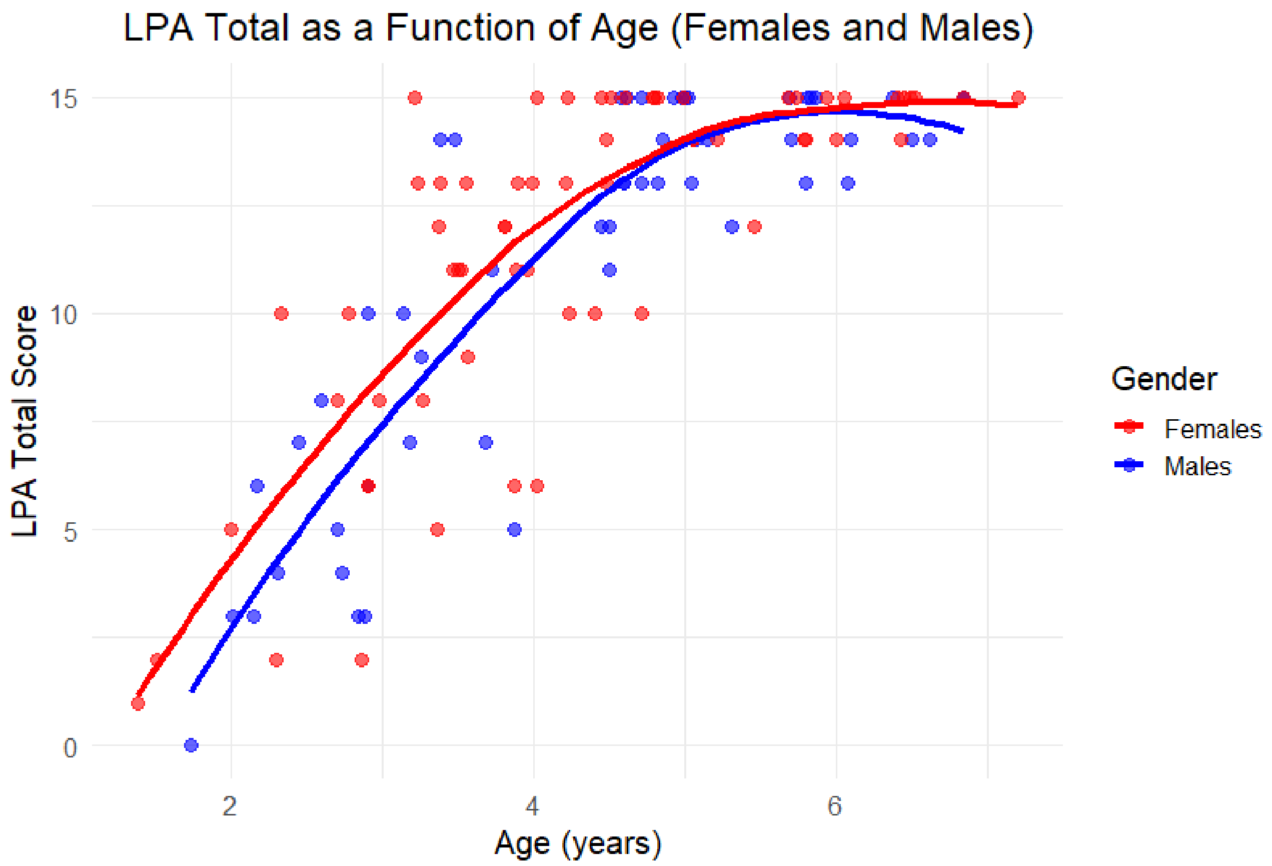

**Figure S2. Clinician-observed total LPA score as a function of age in neurotypical children, with males and females shown separately. Markers represent LPA scores in individual children. Females scored slightly higher than males, although this difference was not statistically significant (Mann-Whitney U test: $p = 0.58$).**

### Percentage of children demonstrating understanding of items in the clinician-observed Language Phenotype Assessment (LPA)

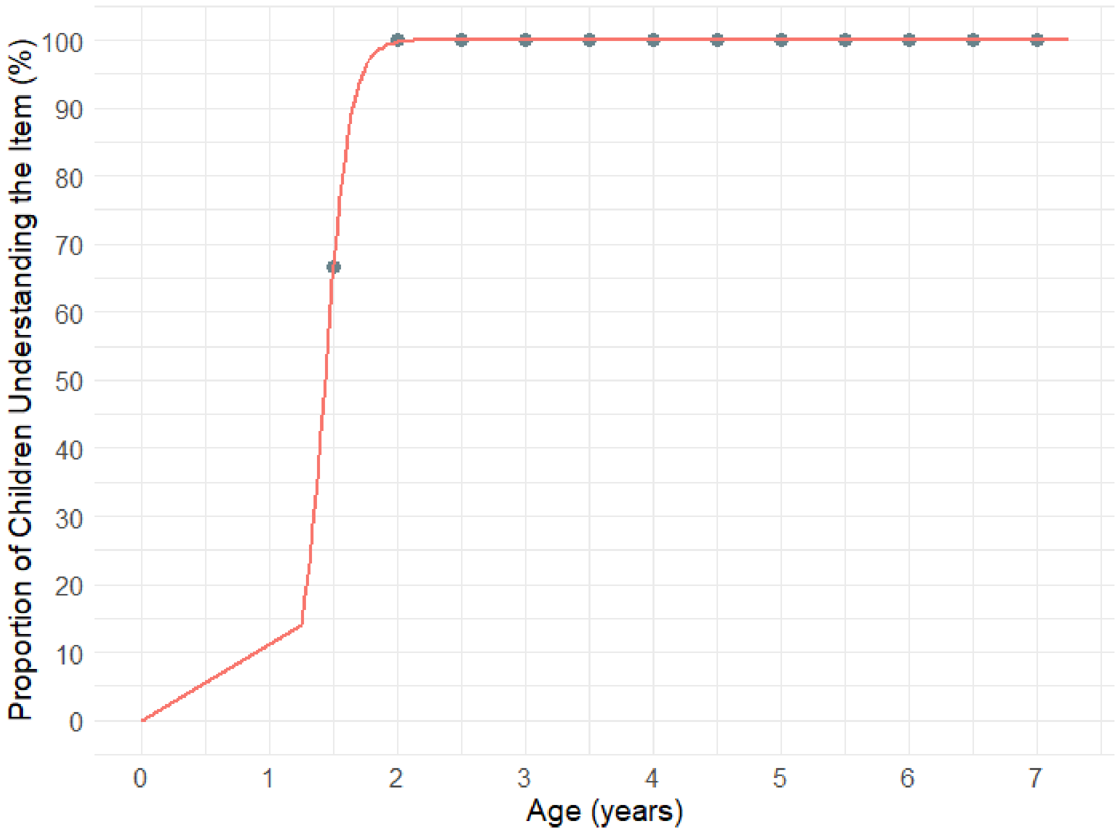

**Figure S3. Percentage of children demonstrating understanding of clinician-observed LPA Item 1: 'Give me an animal.' 50% of children attained this item by 1.4 years. Markers represent the proportion of children who demonstrated understanding of the item, calculated in 0.5-year bins.**

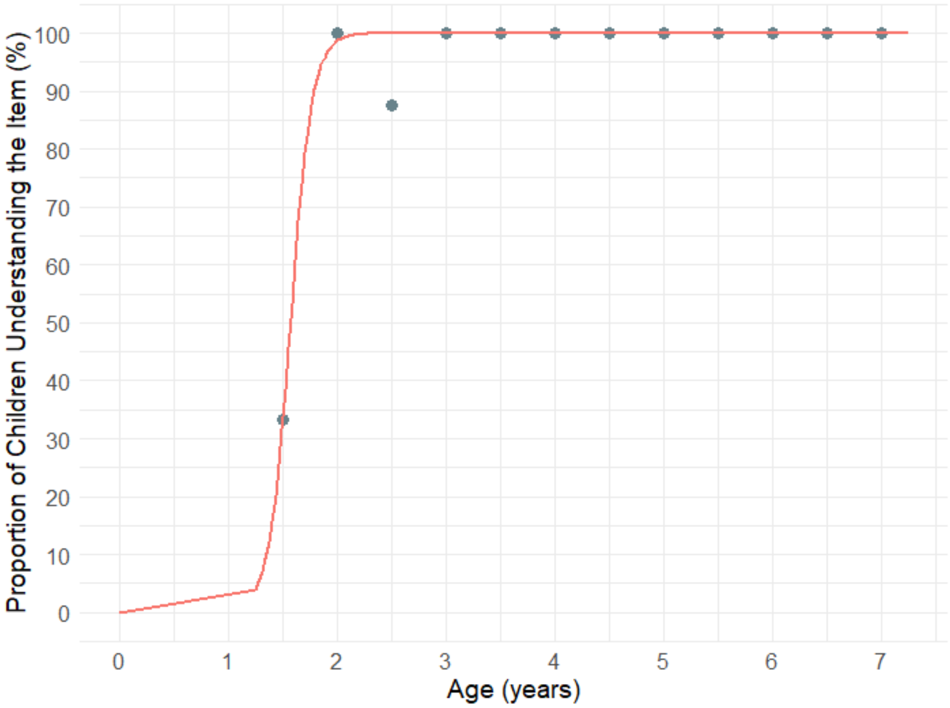

**Figure S4. Percentage of children demonstrating understanding of clinician-observed LPA Item 2: 'Give a cup/pencil to an animal.' 50% of children attained this item by 1.6 years. Markers represent the proportion of children who demonstrated understanding of the item, calculated in 0.5-year bins.**

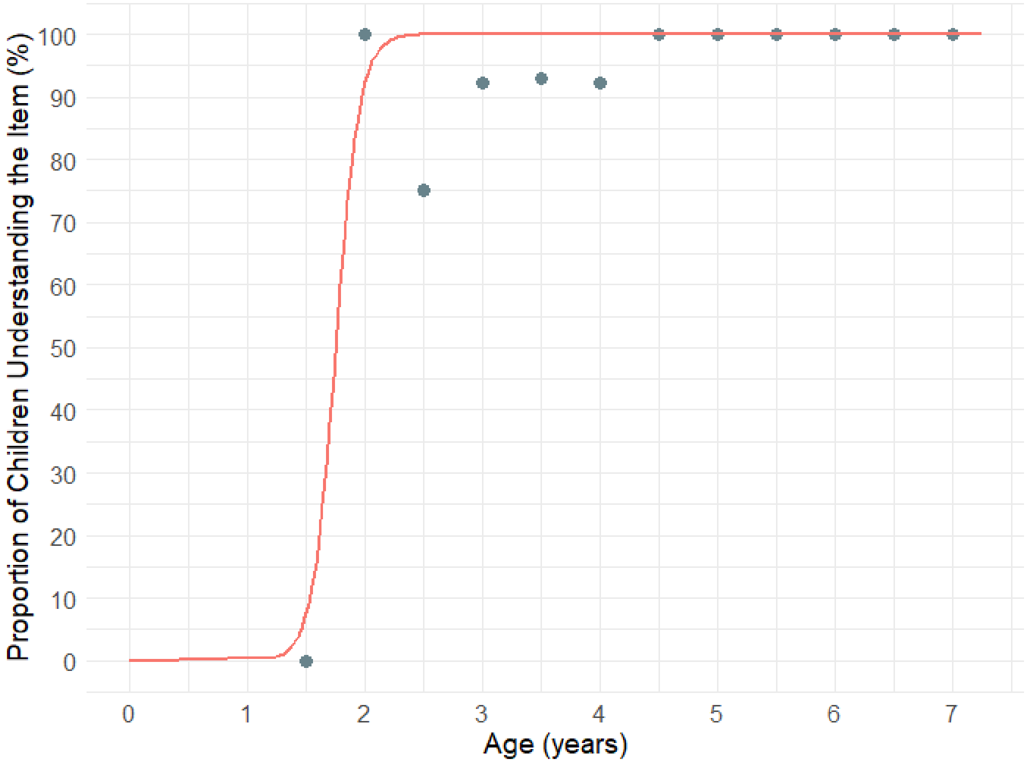

**Figure S5. Percentage of children demonstrating understanding of clinician-observed LPA Item 3: 'Take an animal to another animal.' 50% of children attained this item by 1.8 years. Markers represent the proportion of children who demonstrated understanding of the item, calculated in 0.5-year bins.**

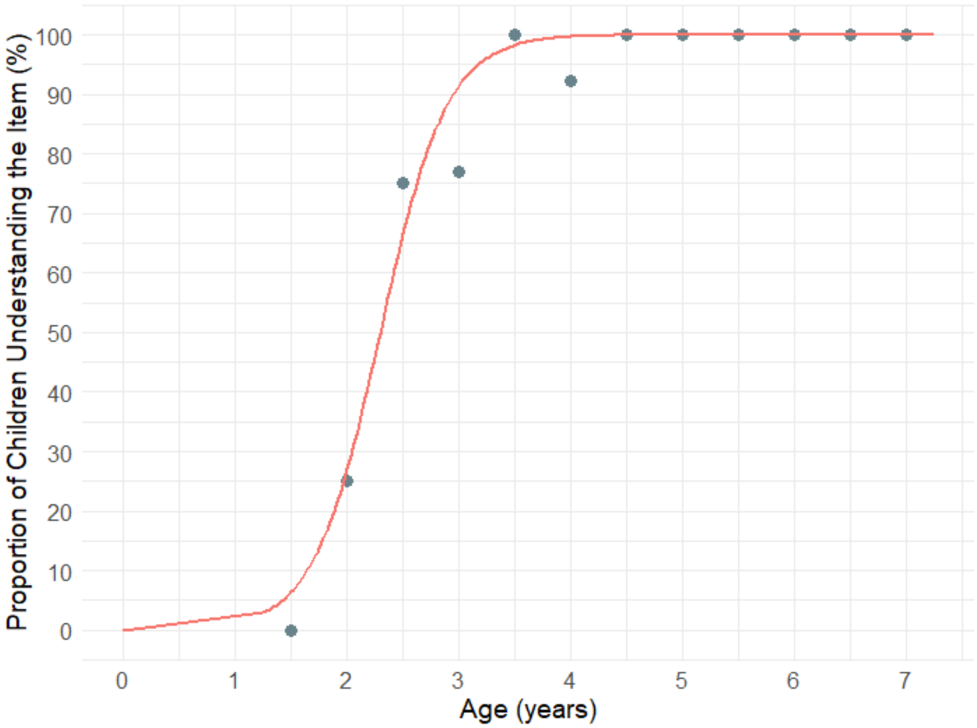

**Figure S6. Percentage of children demonstrating understanding of clinician-observed LPA Item 4: 'Color integration.' 50% of children attained this item by 2.3 years. Markers represent the proportion of children who demonstrated understanding of the item, calculated in 0.5-year bins.**

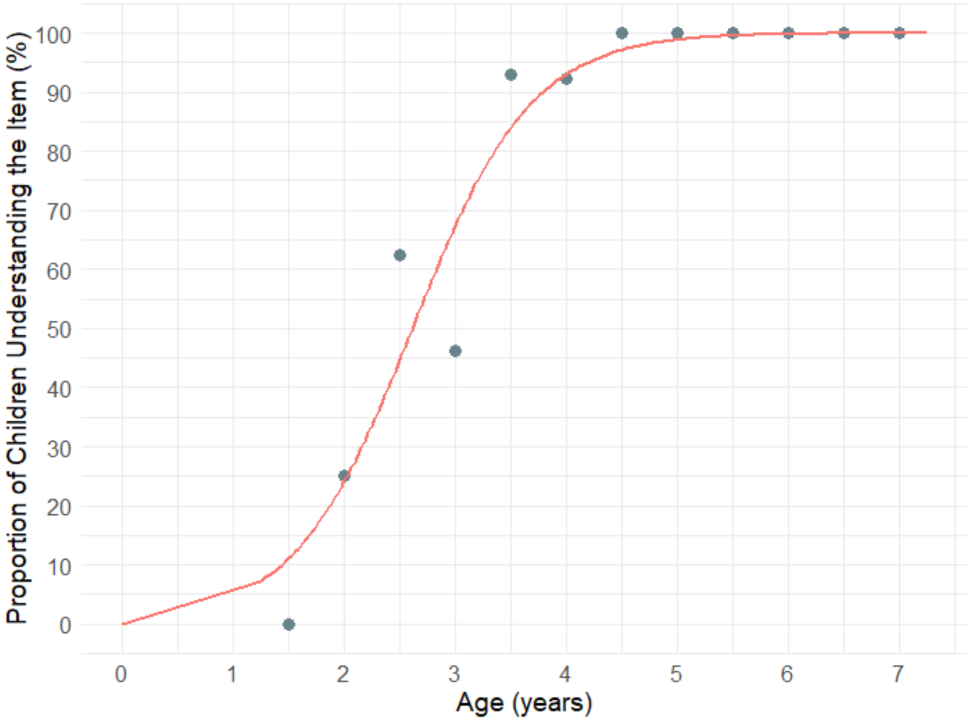

**Figure S7. Percentage of children demonstrating understanding of clinician-observed LPA Item 5: 'Size integration.' 50% of children attained this item by 2.6 years. Markers represent the proportion of children who demonstrated understanding of the item, calculated in 0.5-year bins.**

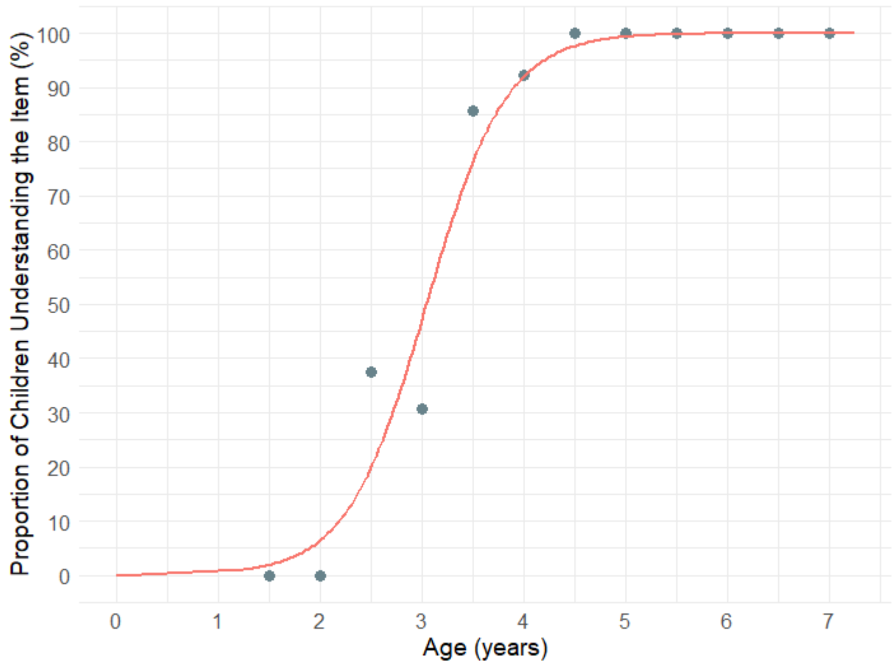

**Figure S8. Percentage of children demonstrating understanding of clinician-observed LPA Item 6: 'Color and size integration.' 50% of children attained this item by 3.0 years. Markers represent the proportion of children who demonstrated understanding of the item, calculated in 0.5-year bins.**

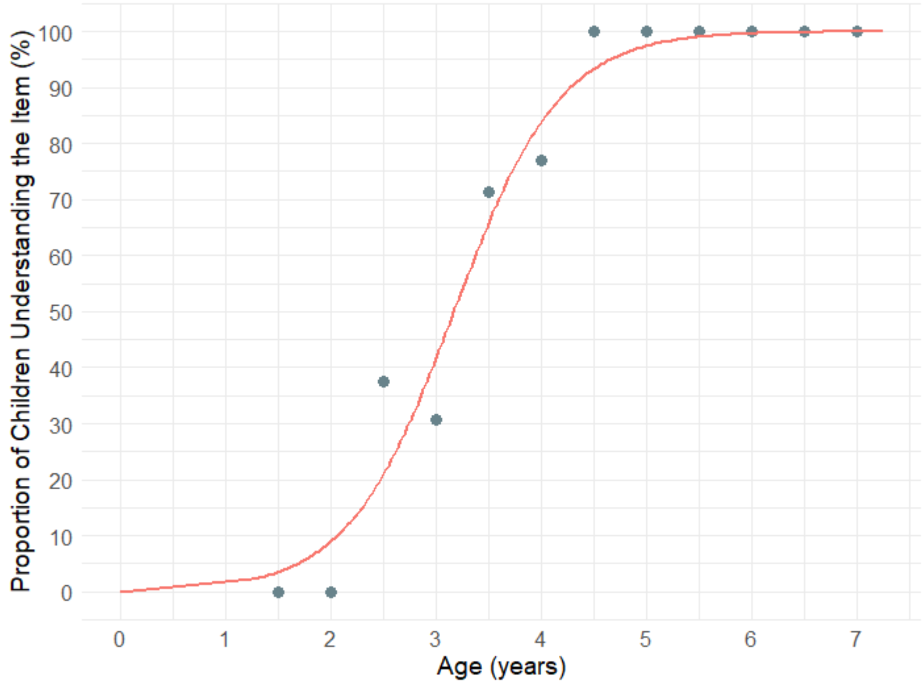

**Figure S9. Percentage of children demonstrating understanding of clinician-observed LPA Item 7: 'Number integration.' 50% of children attained this item by 3.2 years. Markers represent the proportion of children who demonstrated understanding of the item, calculated in 0.5-year bins.**

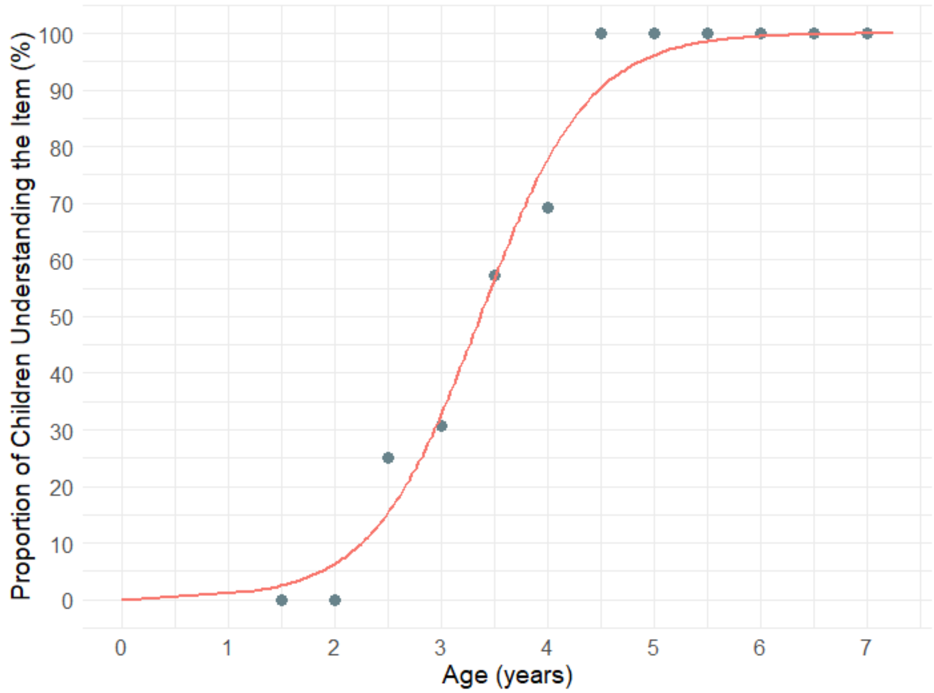

**Figure S10. Percentage of children demonstrating understanding of clinician-observed LPA Item 8: 'Number and color integration.' 50% of children attained this item by 3.4 years. Markers represent the proportion of children who demonstrated understanding of the item, calculated in 0.5-year bins.**

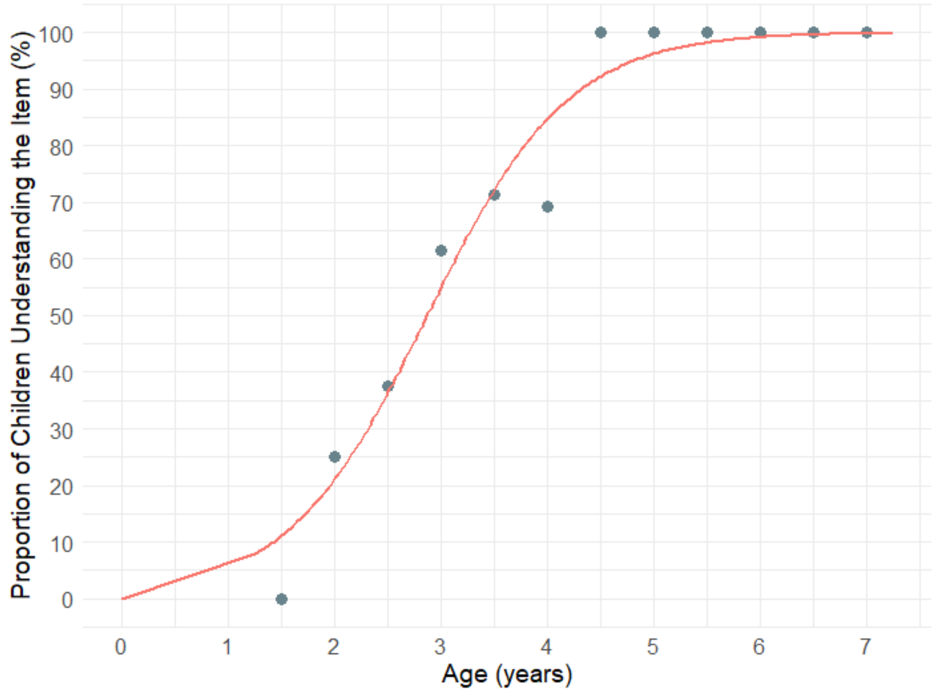

**Figure S11. Percentage of children demonstrating understanding of clinician-observed LPA Item 9: 'Stacking cups canonical word order.' 50% of children attained this item by 2.9 years. Markers represent the proportion of children who demonstrated understanding of the item, calculated in 0.5-year bins.**

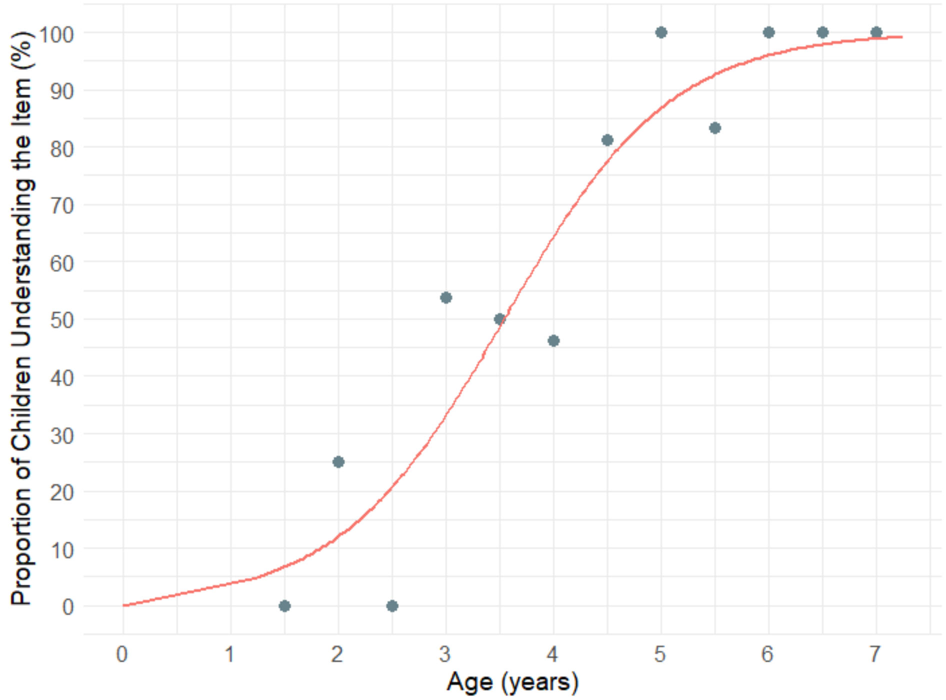

**Figure S12. Percentage of children demonstrating understanding of clinician-observed LPA Item 10: 'Stacking cups noncanonical word order.' 50% of children attained this item by 3.5 years. Markers represent the proportion of children who demonstrated understanding of the item, calculated in 0.5-year bins.**

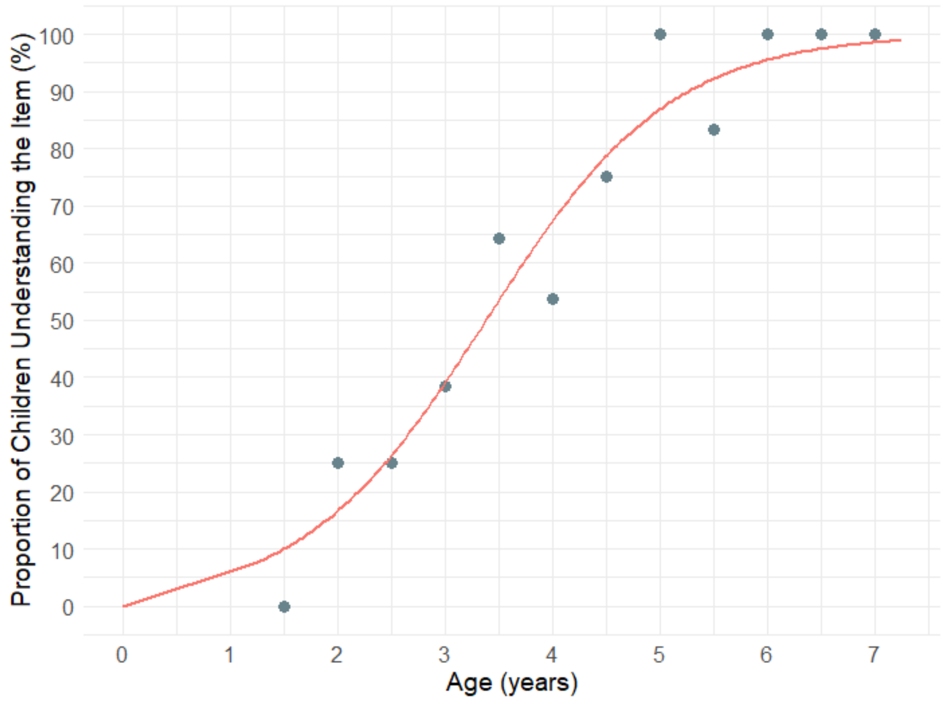

**Figure S13. Percentage of children demonstrating understanding of clinician-observed LPA Item 11: 'Spatial prepositions canonical word order.' 50% of children attained this item by 3.4 years. Markers represent the proportion of children who demonstrated understanding of the item, calculated in 0.5-year bins.**

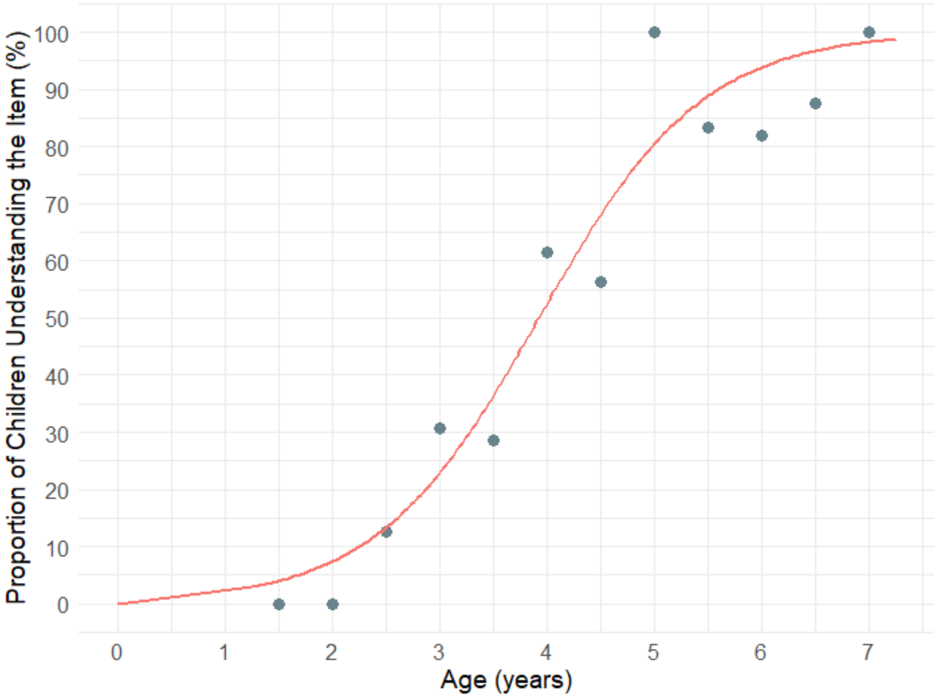

**Figure S14. Percentage of children demonstrating understanding of clinician-observed LPA Item 12: 'Spatial prepositions noncanonical word order.' 50% of children attained this item by 3.9 years. Markers represent the proportion of children who demonstrated understanding of the item, calculated in 0.5-year bins.**

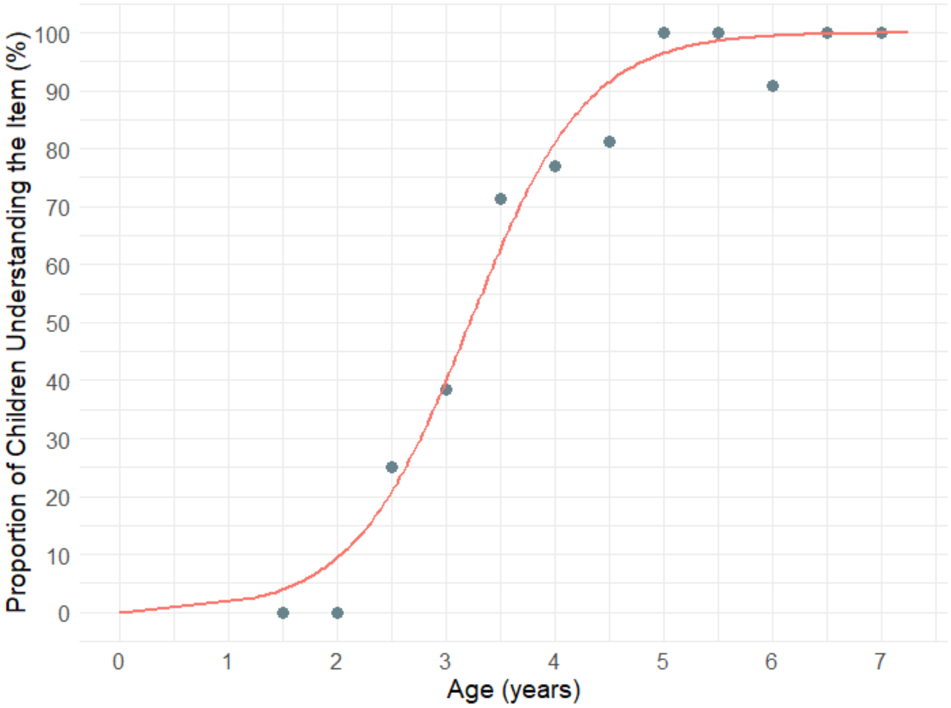

**Figure S15. Percentage of children demonstrating understanding of clinician-observed LPA Item 13: 'Spatial prepositions behind, in front, between.' 50% of children attained this item by 3.2 years. Markers represent the proportion of children who demonstrated understanding of the item, calculated in 0.5-year bins.**

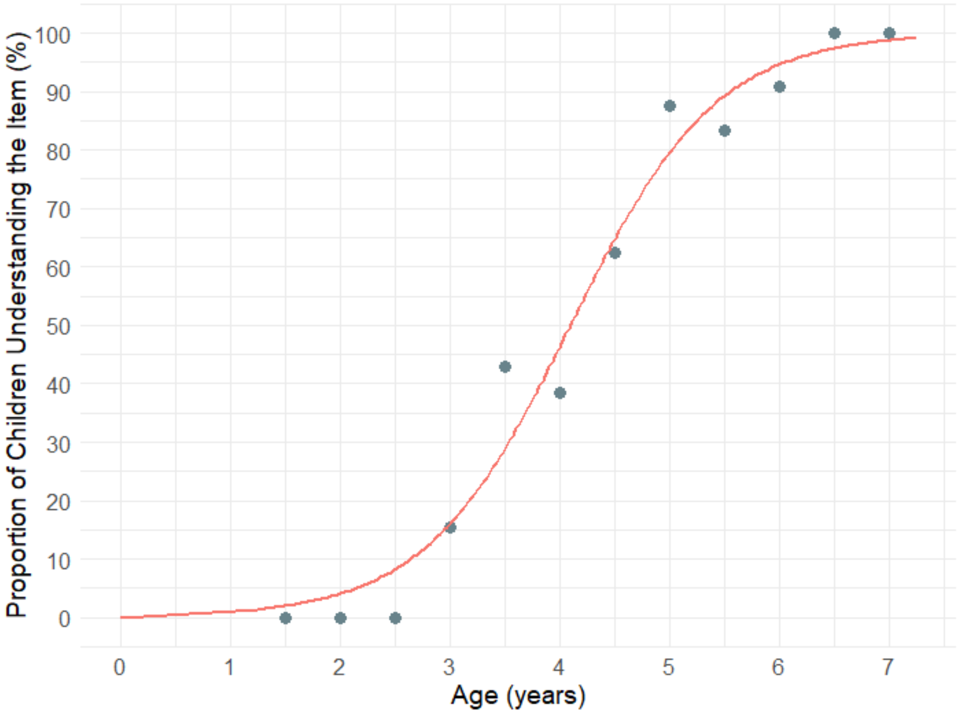

**Figure S16. Percentage of children demonstrating understanding of clinician-observed LPA Item 14: 'Mental reasoning with an object and a subject.' 50% of children attained this item by 4.1 years. Markers represent the proportion of children who demonstrated understanding of the item, calculated in 0.5-year bins.**

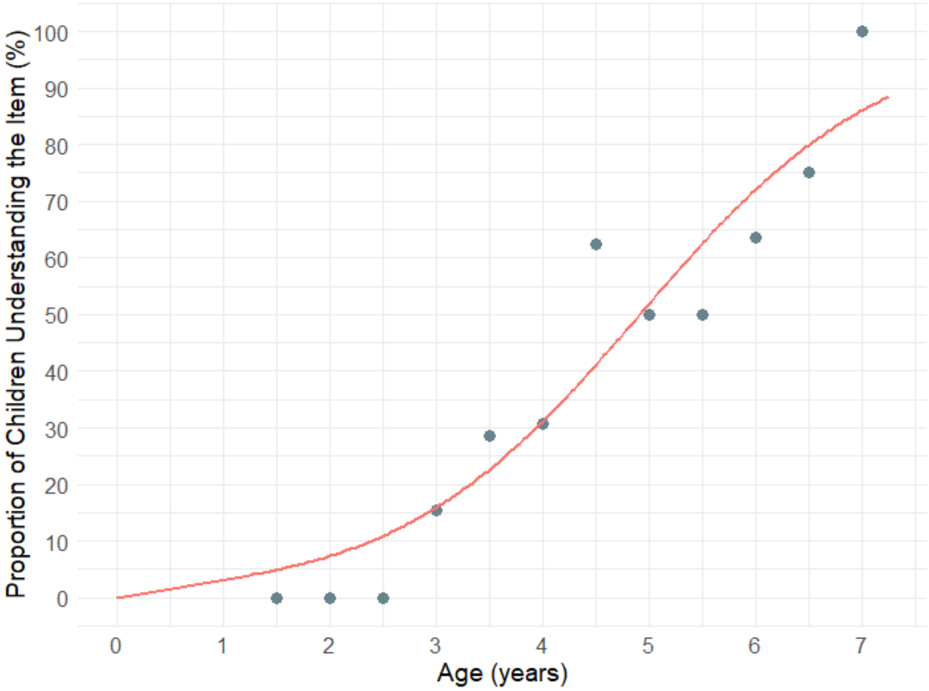

**Figure S18. Percentage of children demonstrating understanding of clinician-observed LPA Item 15: 'Mental reasoning with an object and a subject – passive voice.' 50% of children attained this item by 4.9 years. Markers represent the proportion of children who demonstrated understanding of the item, calculated in 0.5-year bins.**

**Table S1. Median age of attainment for each LPA item (years).**

| Item | Median age of attainment |
|---|---|
| **1. Give me an animal** | 1.4 |
| **2. Give a cup/pencil to an animal** | 1.6 |
| **3. Take an animal to another animal** | 1.75 |
| **4. Color integration** | 2.3 |
| **5. Size integration** | 2.6 |
| **6. Color and size integration** | 3 |
| **7. Number integration** | 3.2 |
| **8. Number and color integration** | 3.4 |
| **9. Stacking cups canonical word order** | 2.9 |
| **10. Stacking cups noncanonical word order** | 3.5 |
| **11. Spatial prepositions canonical word order** | 3.4 |
| **12. Spatial prepositions noncanonical word order** | 3.9 |
| **13. Spatial prepositions behind, in front, between** | 3.2 |
| **14. Mental reasoning with an object and a subject** | 4.1 |
| **15. Mental reasoning with an object and a subject – passive voice** | 4.9 |
| **Command Phenotype (items 1 to 3)** | 1.6 |
| **Modifier Phenotype (items 4 to 8)** | 3.0 |
| **Syntactic Phenotype (items 9 to 15)** | 3.7 |

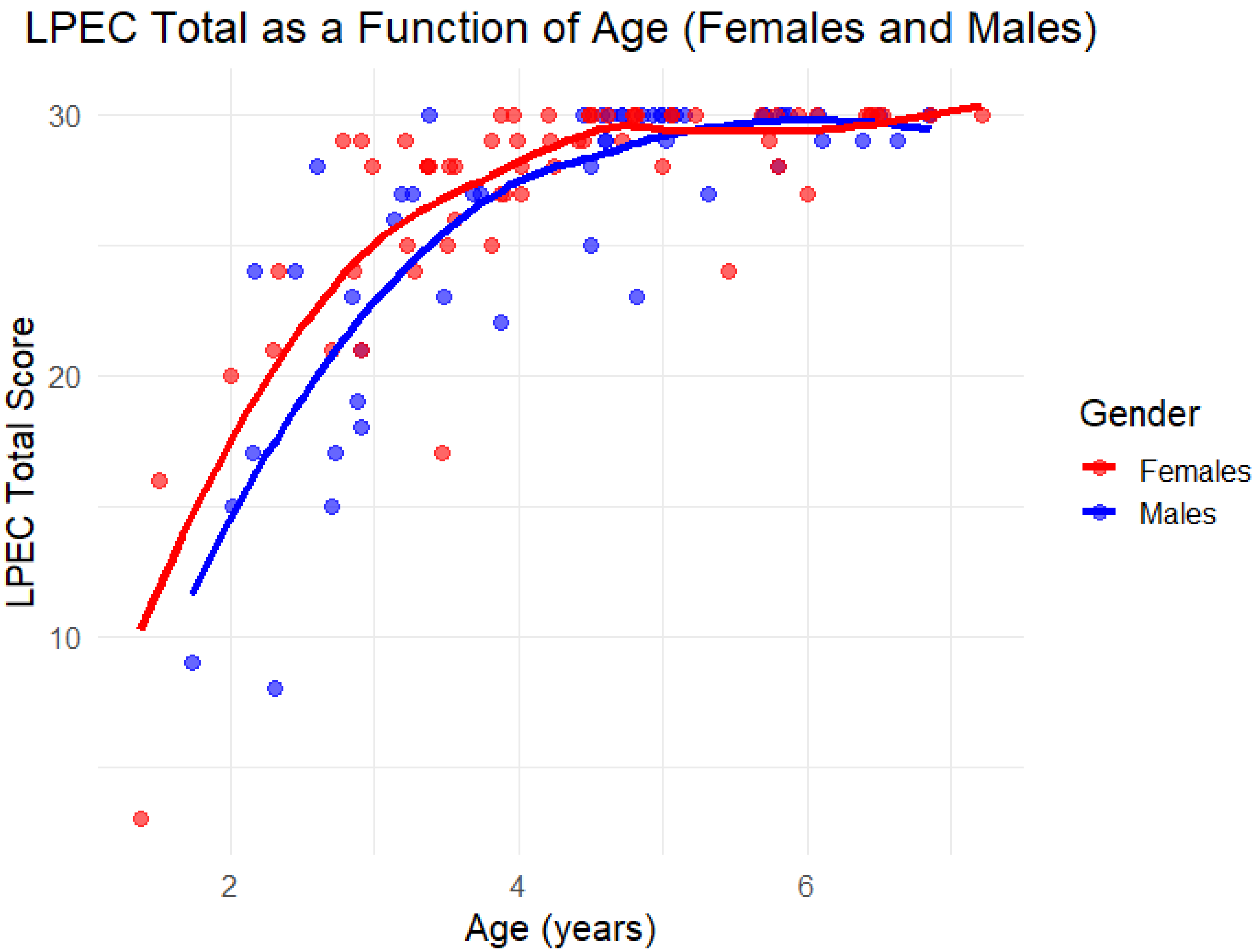

**Figure S19. Parent-reported total LPEC score as a function of age in neurotypical children, with males and females shown separately. Markers represent LPEC scores in individual children. Females scored slightly higher than males (as in the clinician-observed LPA), although this difference was not statistically significant (Mann-Whitney U test: *p* = 0.60).**

### Percentage of children demonstrating understanding of items in the parent-reported Language Phenotype Evaluation Checklist (LPEC)

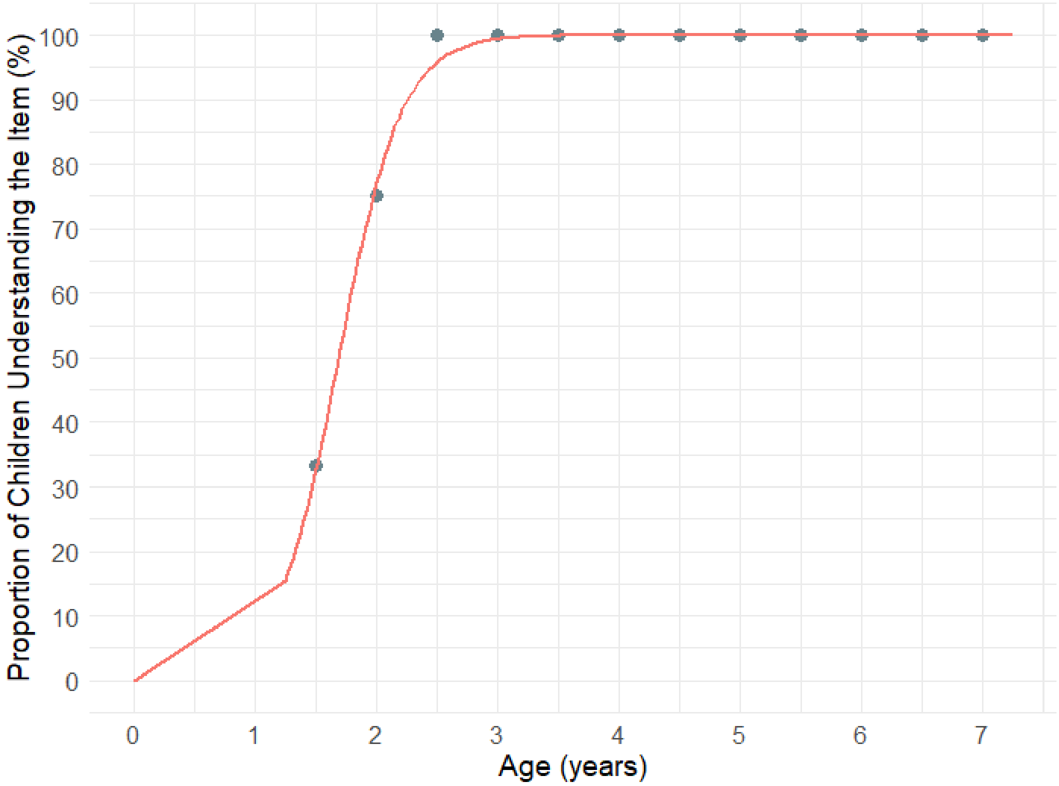

**Figure S20. Percentage of children whose parents responded 'Very True' to LPEC Item 1: Knows names of common objects (cup, chair, car, pencil, etc.). 50% of children attained this item by 1.7 years. Markers represent the proportion of children who demonstrated understanding of the item, calculated in 0.5-year bins.**

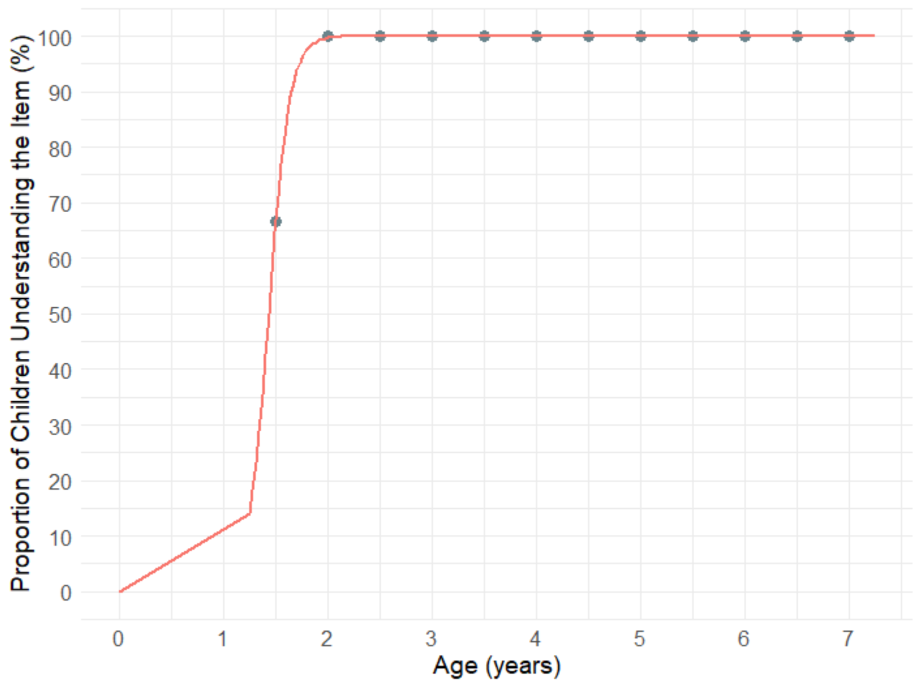

**Figure S21. Percentage of children whose parents responded 'Very True' to LPEC Item 2: Understands commands with pointing (e.g., if you point to the cup and say 'bring me the cup'). 50% of children attained this item by 1.4 years. Markers represent the proportion of children who demonstrated understanding of the item, calculated in 0.5-year bins.**

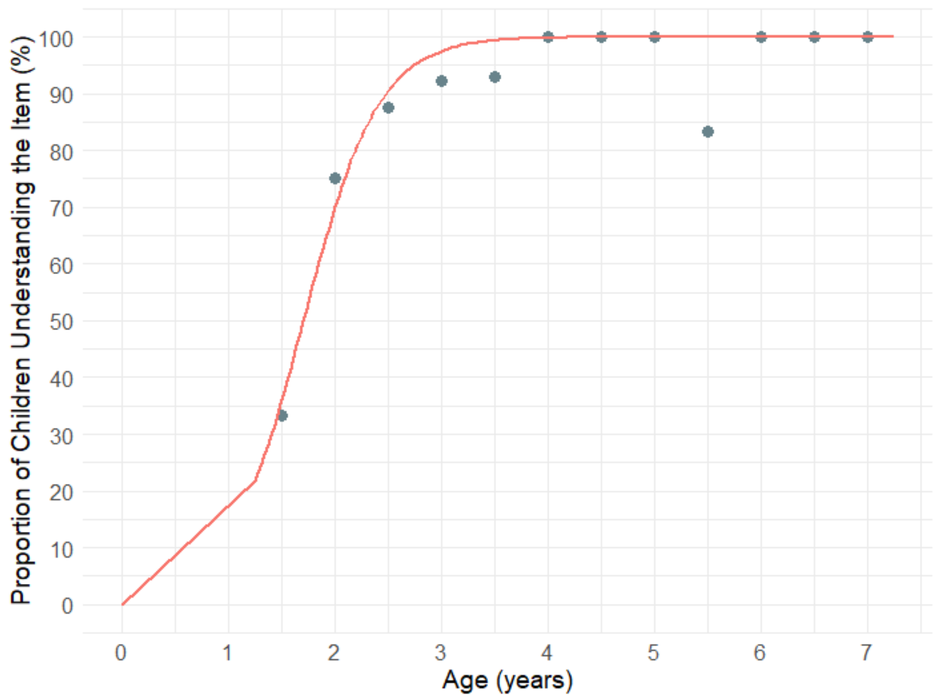

**Figure S22. Percentage of children whose parents responded 'Very True' to LPEC Item 3: Understands commands without pointing (e.g., 'bring me the cup'). 50% of children attained this item by 1.7 years. Markers represent the proportion of children who demonstrated understanding of the item, calculated in 0.5-year bins.**

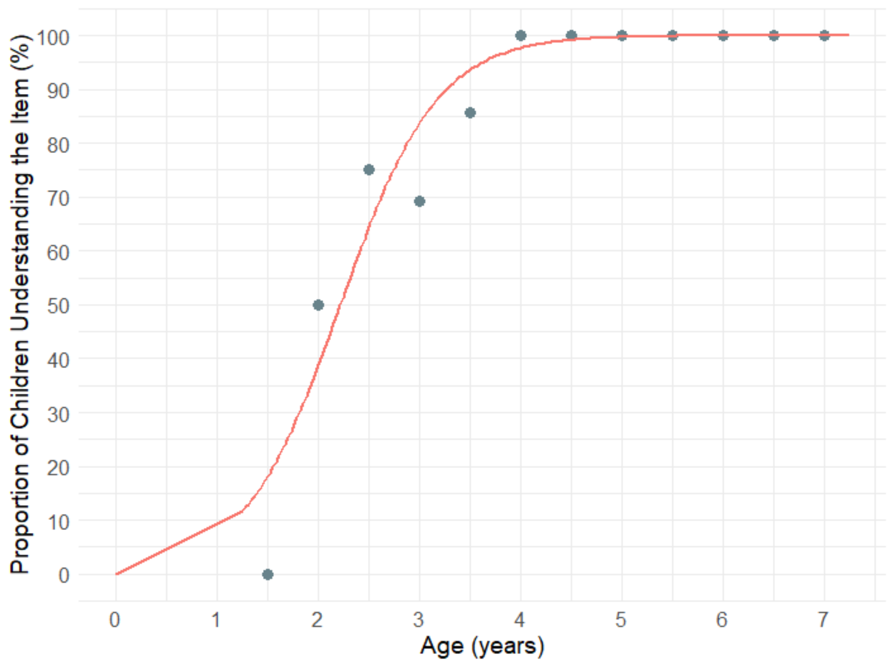

**Figure S23. Percentage of children whose parents responded 'Very True' to LPEC Item 4: Understands color modifiers (e.g., 'green apple' versus 'red apple' versus 'green pencil'). 50% of children attained this item by 2.2 years. Markers represent the proportion of children who demonstrated understanding of the item, calculated in 0.5-year bins.**

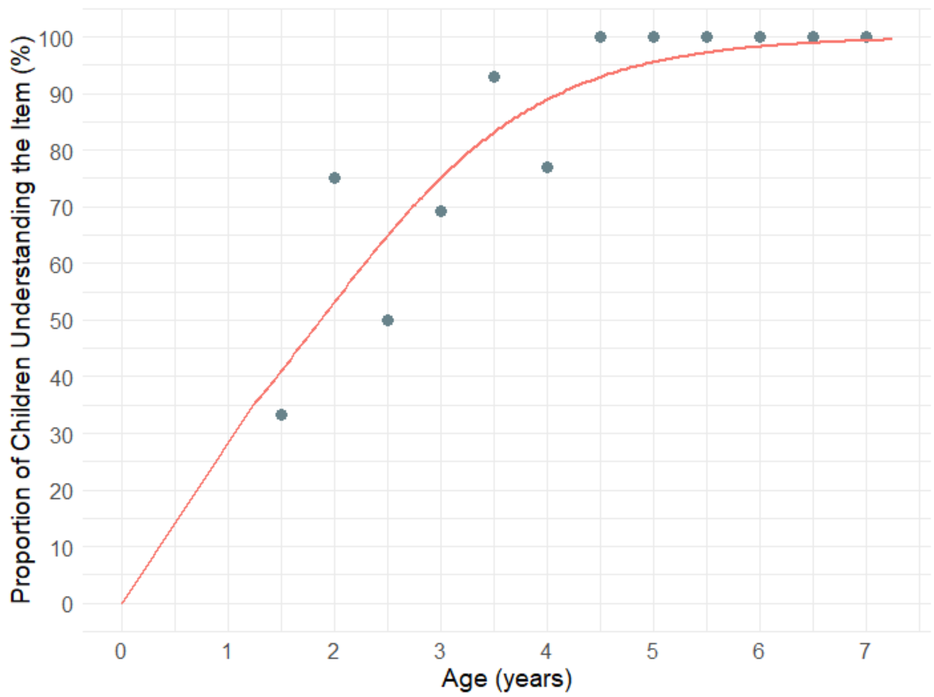

**Figure S24. Percentage of children whose parents responded 'Very True' to LPEC Item 5: Understands size modifiers (e.g., 'big apple' versus 'small apple' versus 'big car'). 50% of children attained this item by 1.9 years. Markers represent the proportion of children who demonstrated understanding of the item, calculated in 0.5-year bins.**

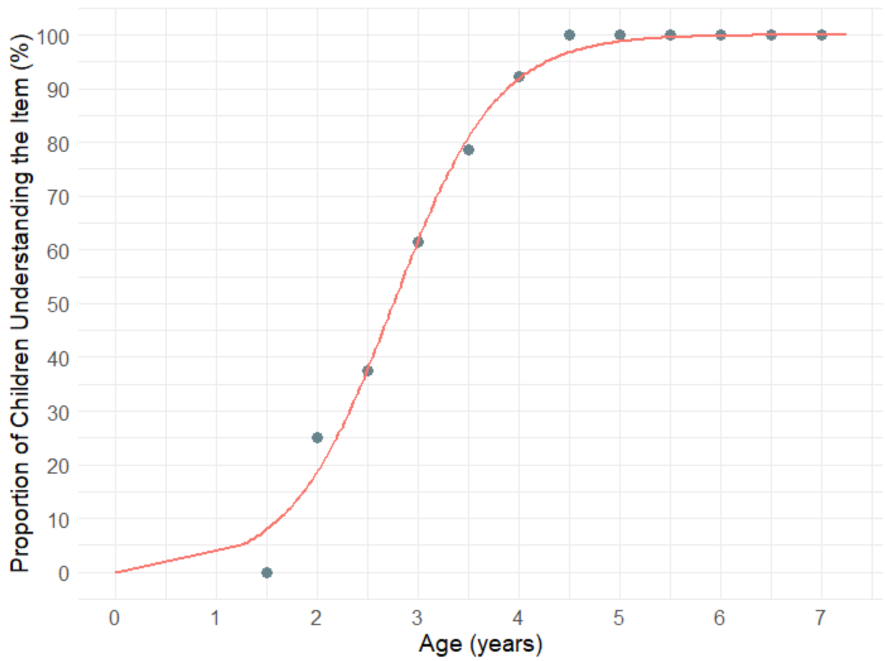

**Figure S25. Percentage of children whose parents responded 'Very True' to LPEC Item 6: Understands several modifiers in a sentence (e.g., 'small green apple'). 50% of children attained this item by 2.8 years. Markers represent the proportion of children who demonstrated understanding of the item, calculated in 0.5-year bins.**

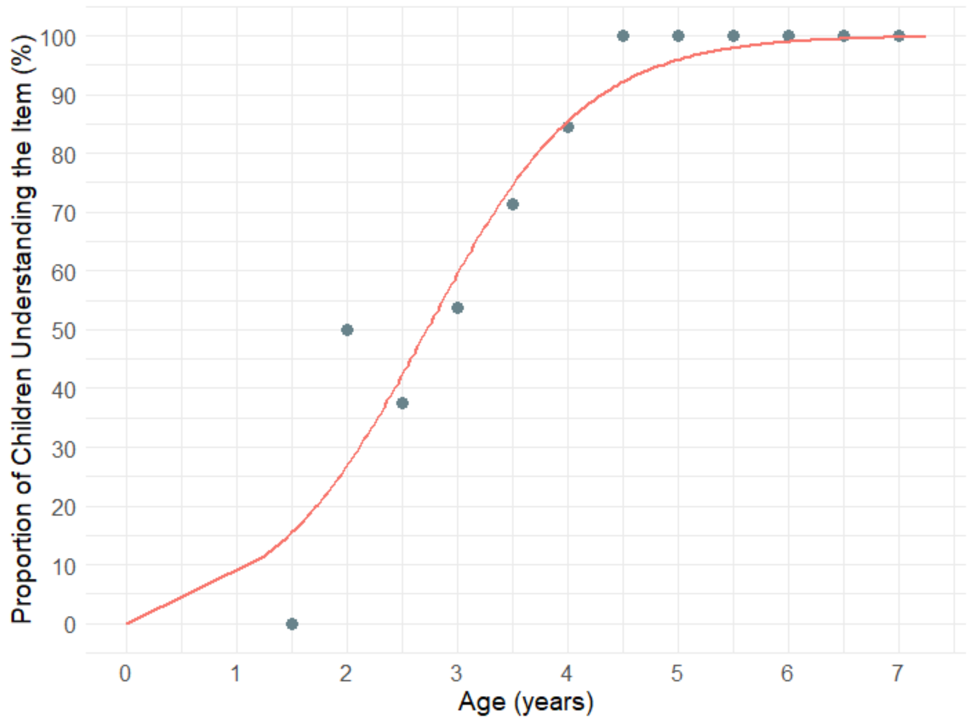

**Figure S26. Percentage of children whose parents responded 'Very True' to LPEC Item 7: Understands size superlatives (can select the largest/smallest object out of a collection of objects). 50% of children attained this item by 2.7 years. Markers represent the proportion of children who demonstrated understanding of the item, calculated in 0.5-year bins.**

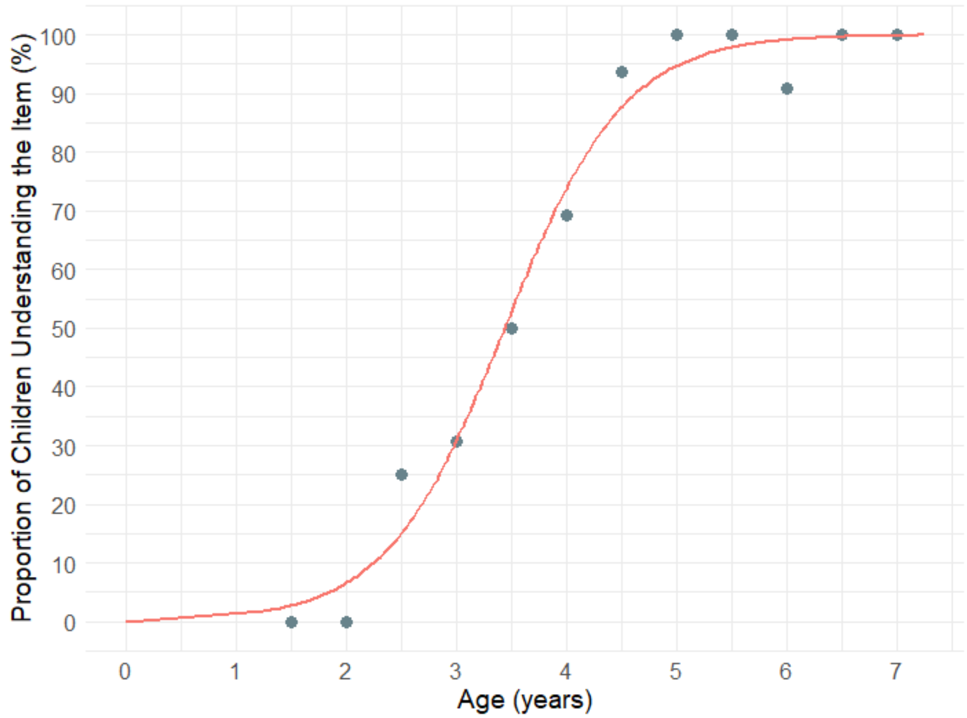

**Figure S27. Percentage of children whose parents responded 'Very True' to LPEC Item 8: Understands NUMBERS (e.g., two apples vs. three apples). 50% of children attained this item by 3.4 years. Markers represent the proportion of children who demonstrated understanding of the item, calculated in 0.5-year bins.**

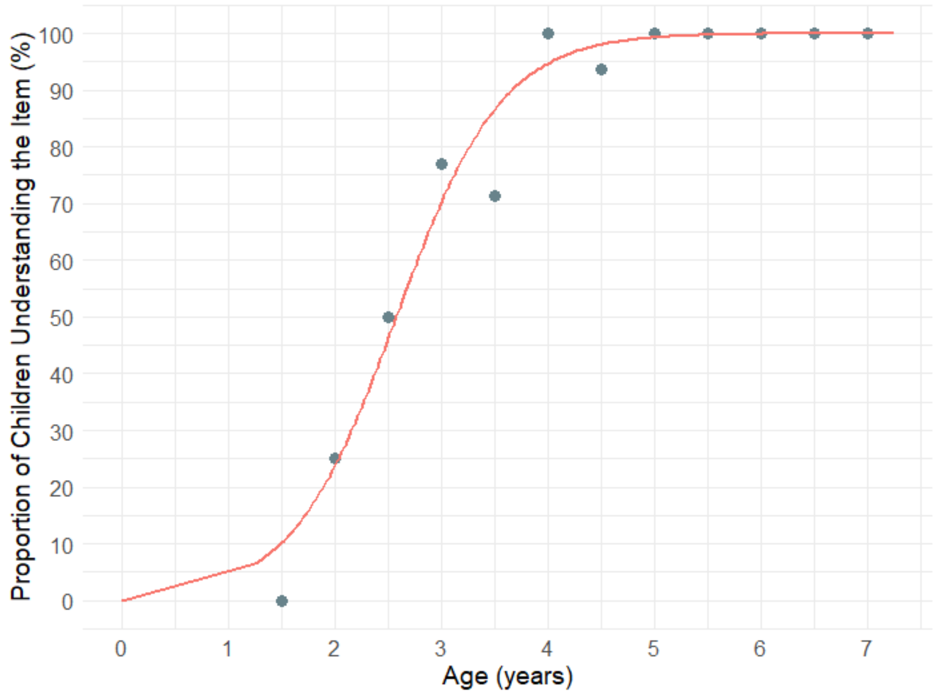

**Figure S28. Percentage of children whose parents responded 'Very True' to LPEC Item 9: Understands possessive pronouns (e.g., your apple vs. her apple). 50% of children attained this item by 2.6 years. Markers represent the proportion of children who demonstrated understanding of the item, calculated in 0.5-year bins.**

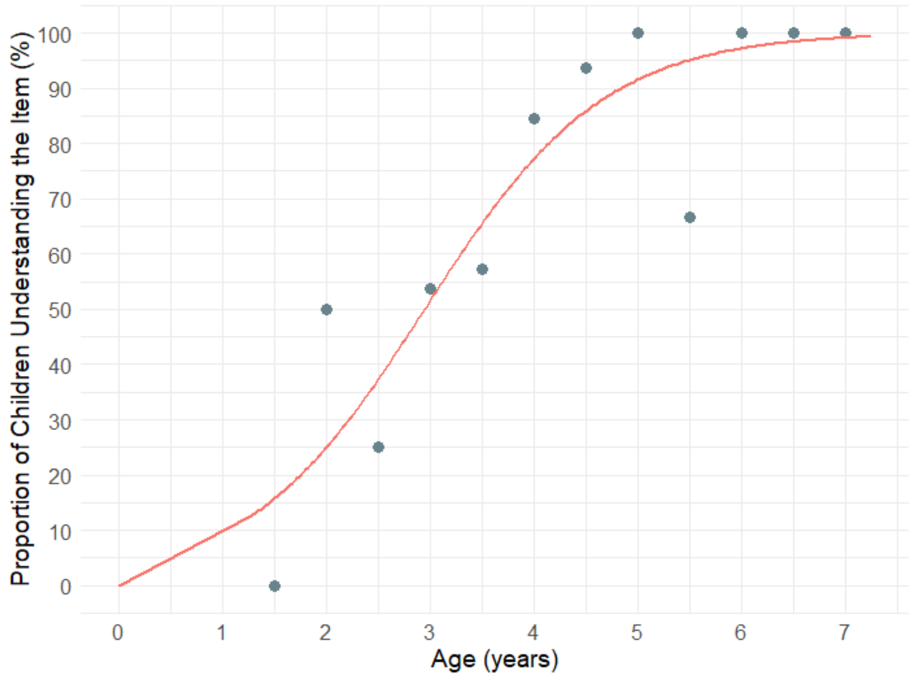

**Figure S29. Percentage of children whose parents responded 'Very True' to LPEC Item 10: Understands spatial prepositions (e.g., put the apple ON TOP of the box versus INSIDE the box vs. BEHIND the box). 50% of children attained this item by 2.9 years. Markers represent the proportion of children who demonstrated understanding of the item, calculated in 0.5-year bins.**

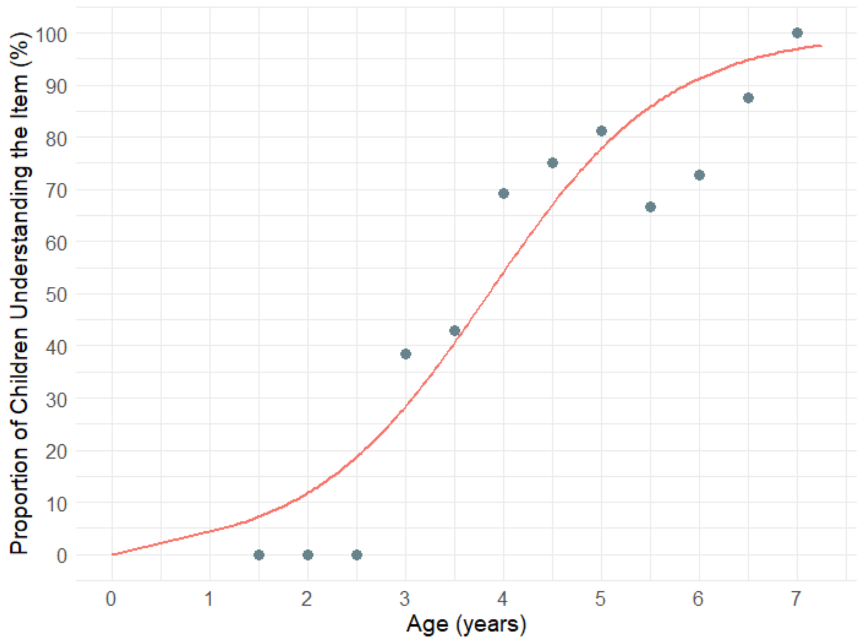

**Figure S30. Percentage of children whose parents responded 'Very True' to LPEC Item 11: Understands verb tenses (e.g., I will eat an apple vs. I ate an apple). 50% of children attained this item by 3.9 years. Markers represent the proportion of children who demonstrated understanding of the item, calculated in 0.5-year bins.**

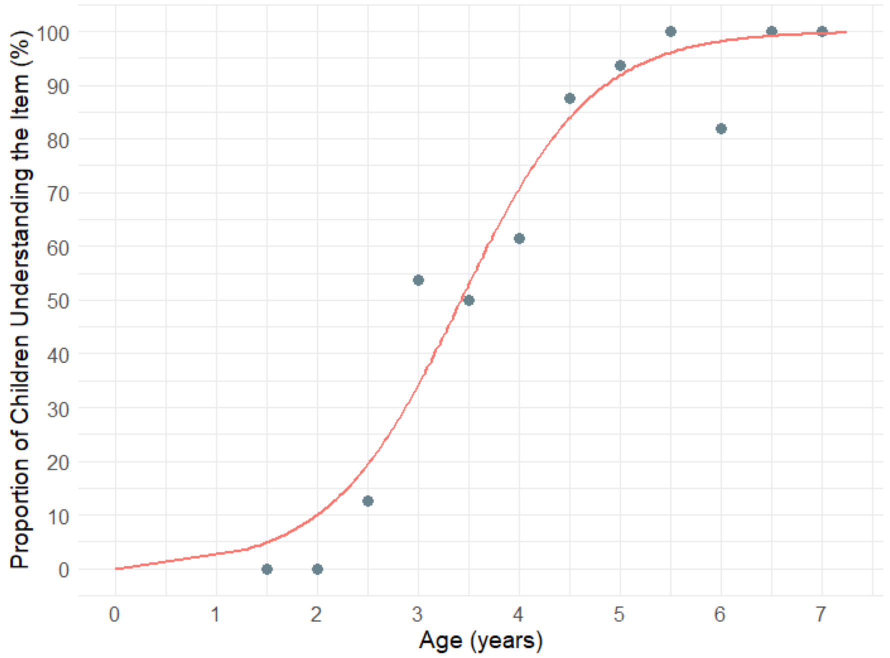

**Figure S31. Percentage of children whose parents responded 'Very True' to LPEC Item 12: Understands the change in meaning when the order of words is changed (e.g., understands the difference between 'a cat ate a mouse' versus 'a mouse ate a cat'). 50% of children attained this item by 3.4 years. Markers represent the proportion of children who demonstrated understanding of the item, calculated in 0.5-year bins.**

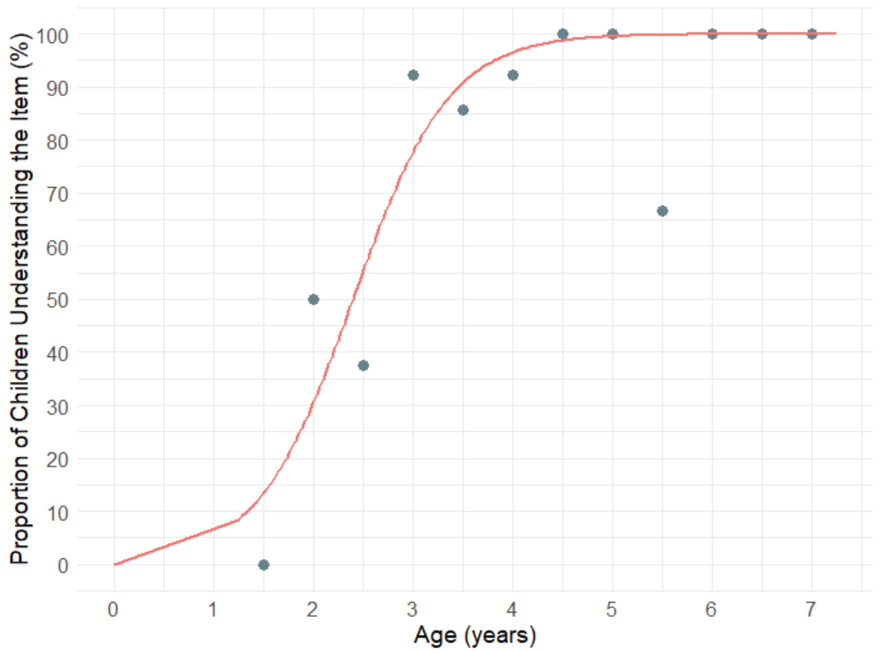

**Figure S32. Percentage of children whose parents responded 'Very True' to LPEC Item 13: Understands simple stories that are read aloud. 50% of children attained this item by 2.4 years. Markers represent the proportion of children who demonstrated understanding of the item, calculated in 0.5-year bins.**

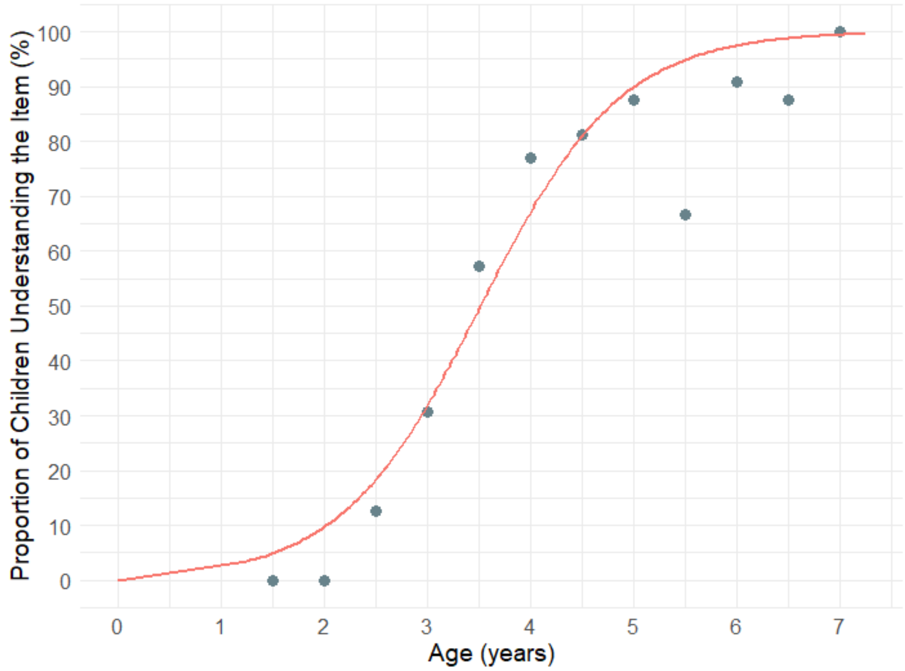

**Figure S33. Percentage of children whose parents responded 'Very True' to LPEC Item 14: Understands elaborate fairy tales that are read aloud (i.e. stories describing FANTASY creatures). 50% of children attained this item by 3.5 years. Markers represent the proportion of children who demonstrated understanding of the item, calculated in 0.5-year bins.**

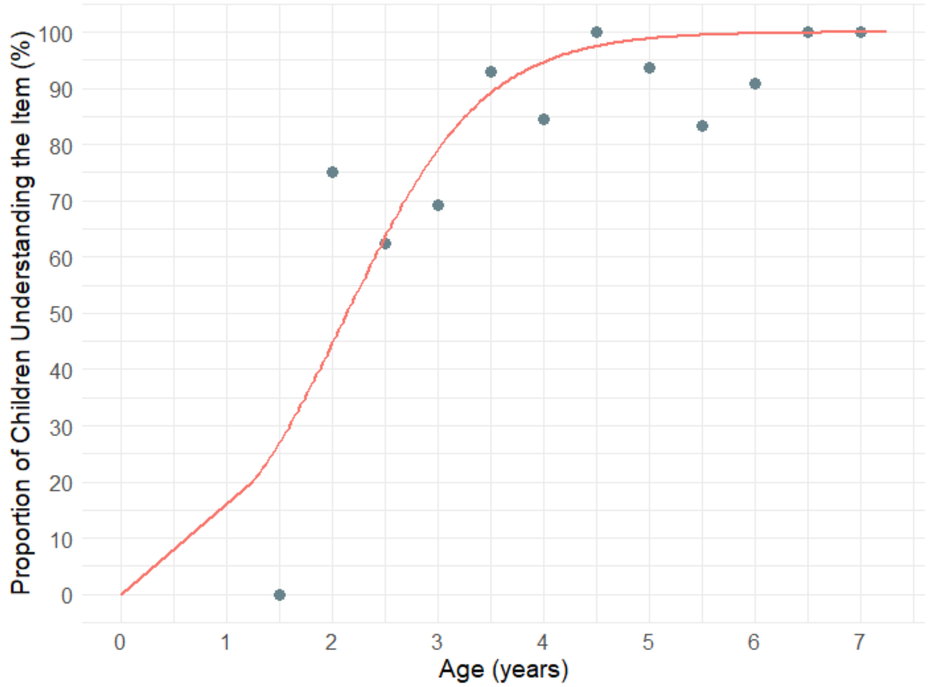

**Figure S34. Percentage of children whose parents responded 'Very True' to LPEC Item 15: Understands explanations about people, objects or situations beyond the immediate surroundings (e.g., "Mom is walking the dog," "The snow has turned to water"). 50% of children attained this item by 2.1 years. Markers represent the proportion of children who demonstrated understanding of the item, calculated in 0.5-year bins.**

**Table S2. Median age of attainment for each LPEC item (years).**

| Item | Median age of attainment |
|---|---|
| 1. Knows names of common objects (cup, chair, car, pencil, etc.) | 1.7 |
| 2. Understands commands with pointing (e.g., if you point to the cup and say 'bring me the cup') | 1.4 |
| 3. Understands commands without pointing (e.g., 'bring me the cup') | 1.7 |
| 4. Understands color modifiers (e.g., 'green apple' versus 'red apple' versus 'green pencil') | 2.2 |
| 5. Understands size modifiers (e.g., 'big apple' versus 'small apple' versus 'big car') | 1.9 |
| 6. Understands several modifiers in a sentence (e.g., 'small green apple') | 2.8 |
| 7. Understands size superlatives (can select the largest/smallest object out of a collection of objects) | 2.7 |
| 8. Understands NUMBERS (e.g., two apples vs. three apples) | 3.4 |
| 9. Understands possessive pronouns (e.g., your apple vs. her apple) | 2.6 |
| 10. Understands spatial prepositions (e.g., put the apple ON TOP of the box versus INSIDE the box vs. BEHIND the box) | 2.9 |
| 11. Understands verb tenses (e.g., I will eat an apple vs. I ate an apple) | 3.9 |
| 12. Understands the change in meaning when the order of words is changed (e.g., understands the difference between 'a cat ate a mouse' versus 'a mouse ate a cat') | 3.4 |
| 13. Understands simple stories that are read aloud | 2.4 |
| 14. Understands elaborate fairy tales that are read aloud (i.e. stories describing FANTASY creatures) | 3.5 |
| 15. Understands explanations about people, objects or situations beyond the immediate surroundings (e.g., "Mom is walking the dog," "The snow has turned to water") | 2.1 |
| Command Phenotype (items 1 to 3) | 1.6 |
| Modifier Phenotype (items 4 to 8) | 2.7 |
| Syntactic Phenotype (items 9 to 15) | 3.0 |

### Additional normative data

In addition to 15-item LPEC, parents provided answers to nine items listed in Table S1. These items represent important development milestones indirectly related to syntactic language acquisition and therefore are presented here. Two items assess representational drawing (Table S3, items 1 and 2), which has been shown to be associated with the syntactic-language-comprehension-phenotype [1]. One item evaluates pretend play (Table S3, item 3), which is a known precursor to syntactic language; lack of pretend play in children with ASD is a strong indicator of challenges in acquisition of the syntactic-language-comprehension-phenotype [2–5]. Additionally, six items measure understanding of complex recursion through arithmetic (Table S3, items 4-9). At an early level, arithmetic is an extension of syntactic-language [6,7]. Interpretation of syntactic sentences requires a degree of reasoning that is similar to that of arithmetic. Compare the following two sentences: 1) “The lion lives under the monkey, who lives under the dog,” and 2) “Mom had five flowers; she gave two flowers to Dad; how many flowers does Mom have now?” While the first sentence could come from a fairy tale and the second from an arithmetic book, the two instructions involve the same executive function that can be characterized as reasoning, syntactic logic, or interpreting complex recursive sentences. The possible answers to each item are: very true (2 points), somewhat true (1 point), not true (0 points).

**Table S3. Additional parent-reported items (LPEC-extension).**

| |
|---|
| 1. Draws a VARIETY of RECOGNIZABLE images (objects, people, animals, etc.) |
| 2. Can draw a NOVEL image following YOUR description (e.g. a three-headed horse) |
| 3. Engages in a VARIETY of make-believe activities (such as: playing house, playing with toy soldiers, building forts and castles, etc.) |
| 4. Can perform simple arithmetic: 2 + 3 = ? |
| 5. Can add larger numbers: 7 + 6 = ? |
| 6. Can perform simple subtraction: 3 – 2 = ? |
| 7. Can subtract larger numbers: 15 – 7 = ? |
| 8. Can perform simple multiplication: 2 × 2 = ? |
| 9. Can multiply larger numbers: 6 × 7 = ? |

### Percentage of children demonstrating understanding of items in the parent-reported LPEC-extension

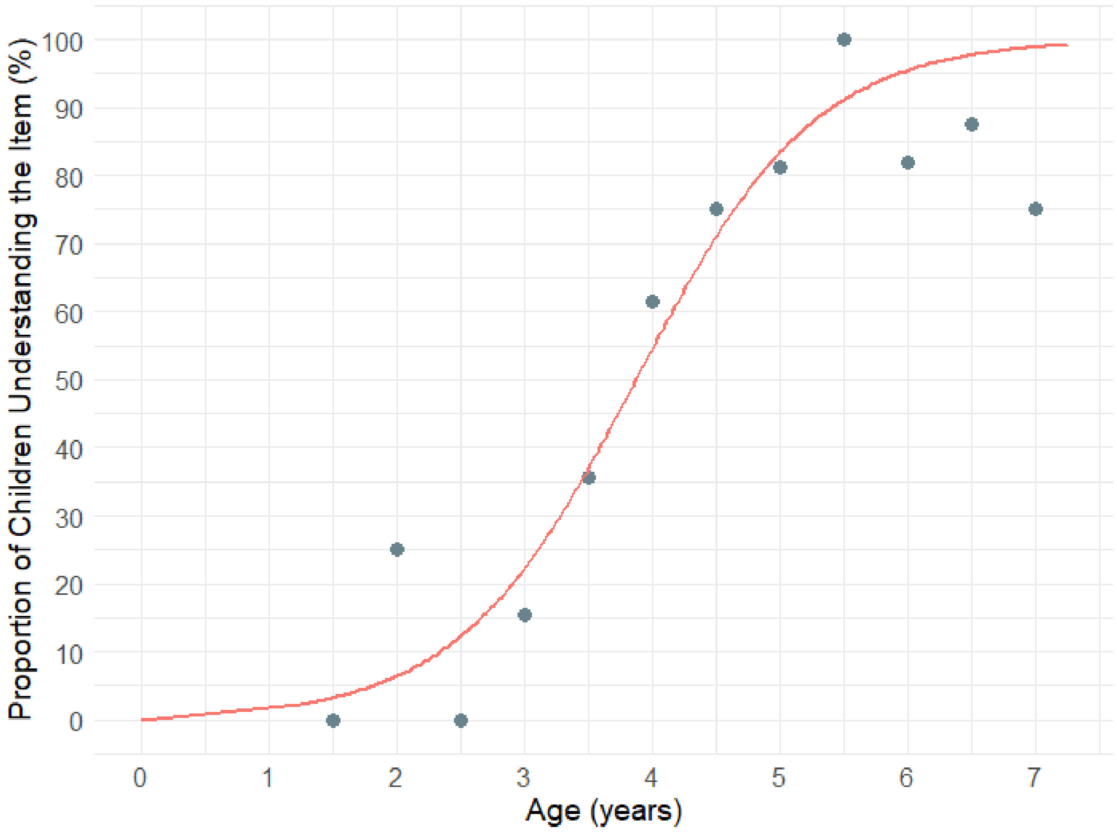

**Figure S35. Percentage of children whose parents responded 'Very True' to Item 1 in Table S3: Draws a VARIETY of RECOGNIZABLE images (objects, people, animals, etc.). 50% of children attained this item by 3.9 years. Markers represent the proportion of children who demonstrated understanding of the item, calculated in 0.5-year bins.**

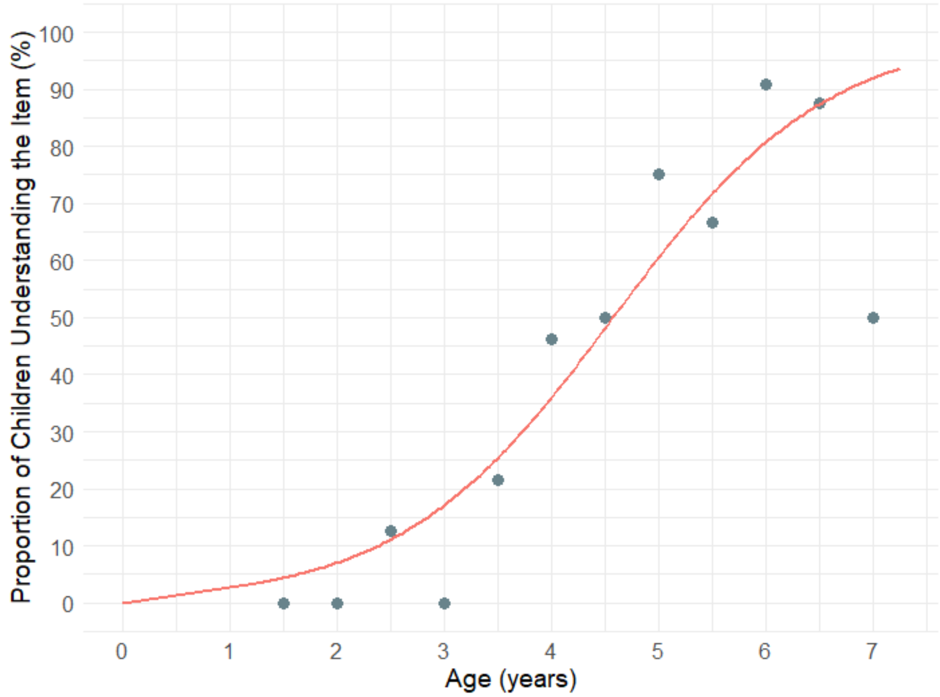

**Figure S36. Percentage of children whose parents responded 'Very True' to Item 2 in Table S3: Can draw a NOVEL image following YOUR description (e.g. a three-headed horse). 50% of children attained this item by 4.6 years. Markers represent the proportion of children who demonstrated understanding of the item, calculated in 0.5-year bins.**

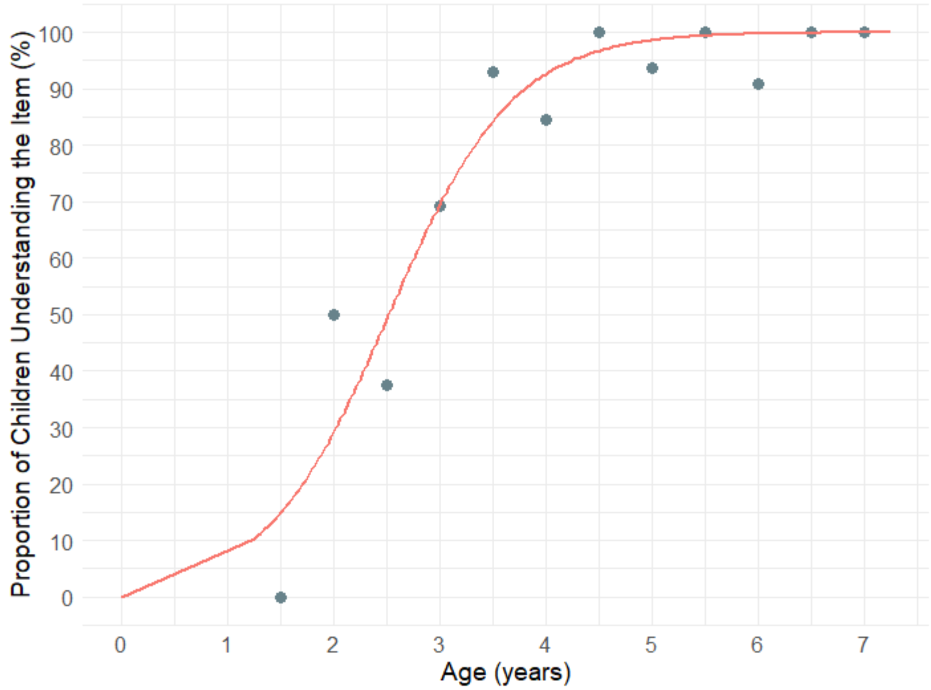

**Figure S37. Percentage of children whose parents responded 'Very True' to Item 3 in Table S3: Engages in a VARIETY of make-believe activities (such as: playing house, playing with toy soldiers, building forts and castles, etc.). 50% of children attained this item by 2.5 years. Markers represent the proportion of children who demonstrated understanding of the item, calculated in 0.5-year bins.**

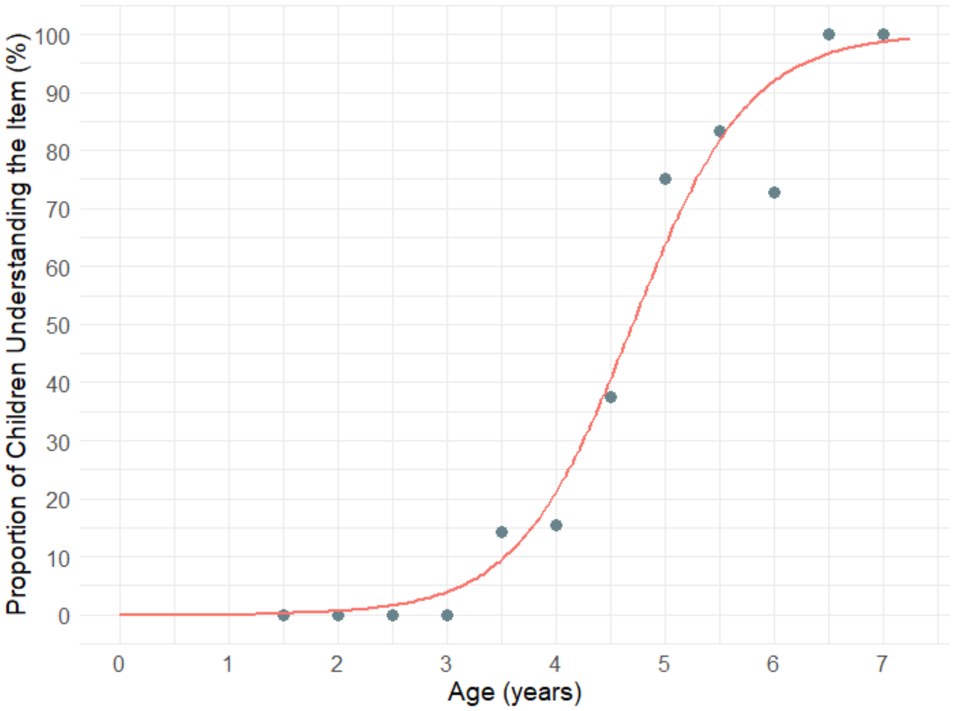

**Figure S38. Percentage of children whose parents responded 'Very True' to Item 4 in Table S3: Can perform simple arithmetic: 2 + 3 = ?. 50% of children attained this item by 4.7 years. Markers represent the proportion of children who demonstrated understanding of the item, calculated in 0.5-year bins.**

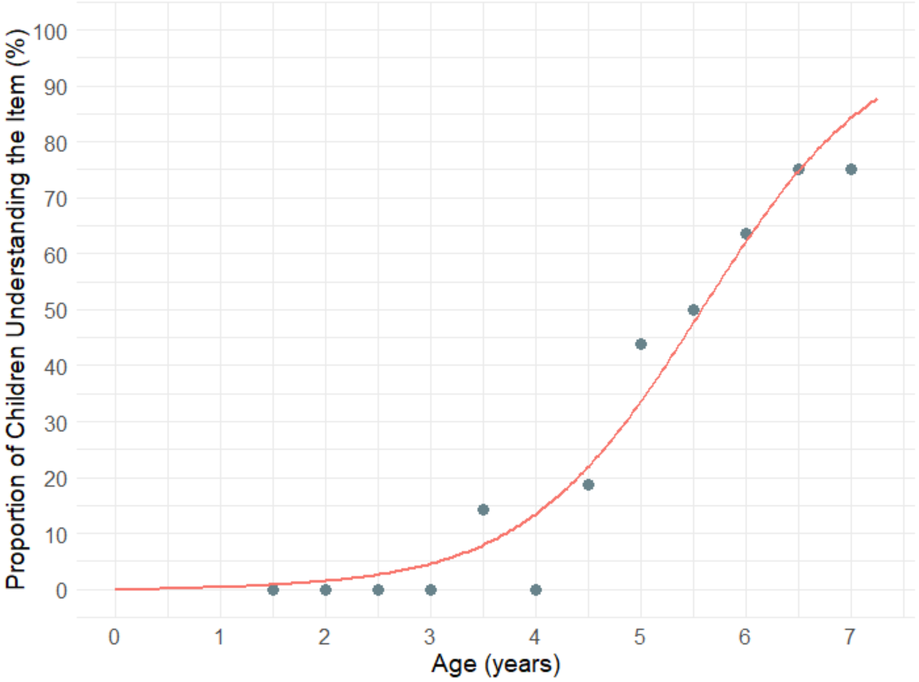

**Figure S39. Percentage of children whose parents responded 'Very True' to Item 5 in Table S3: Can add larger numbers: 7 + 6 = ?. 50% of children attained this item by 5.6 years. Markers represent the proportion of children who demonstrated understanding of the item, calculated in 0.5-year bins.**

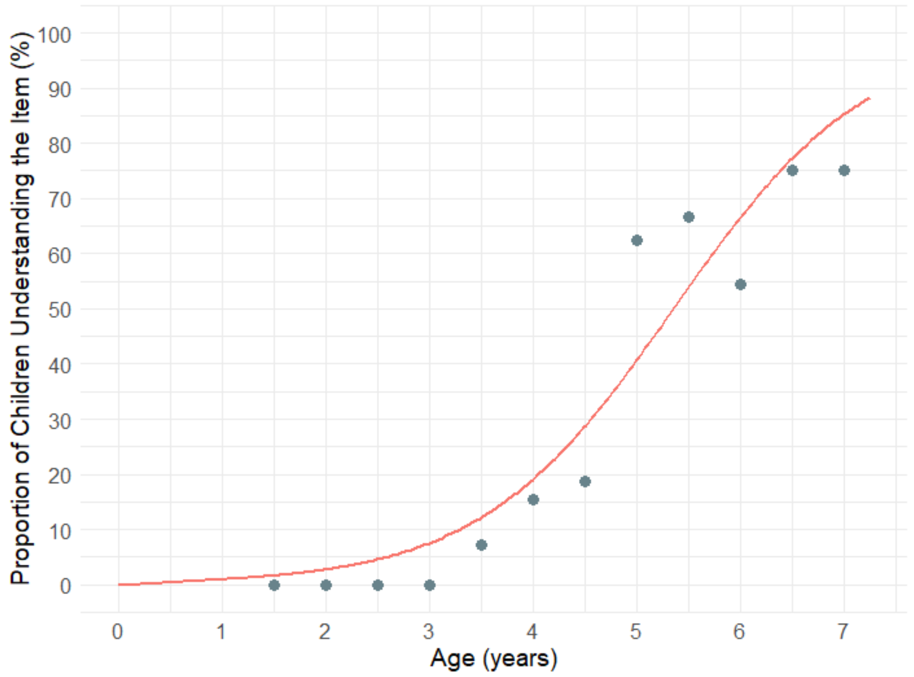

**Figure S40. Percentage of children whose parents responded ‘Very True’ to Item 6 in Table S3: Can perform simple subtraction: 3 – 2 = ?. 50% of children attained this item by 5.4 years. Markers represent the proportion of children who demonstrated understanding of the item, calculated in 0.5-year bins.**

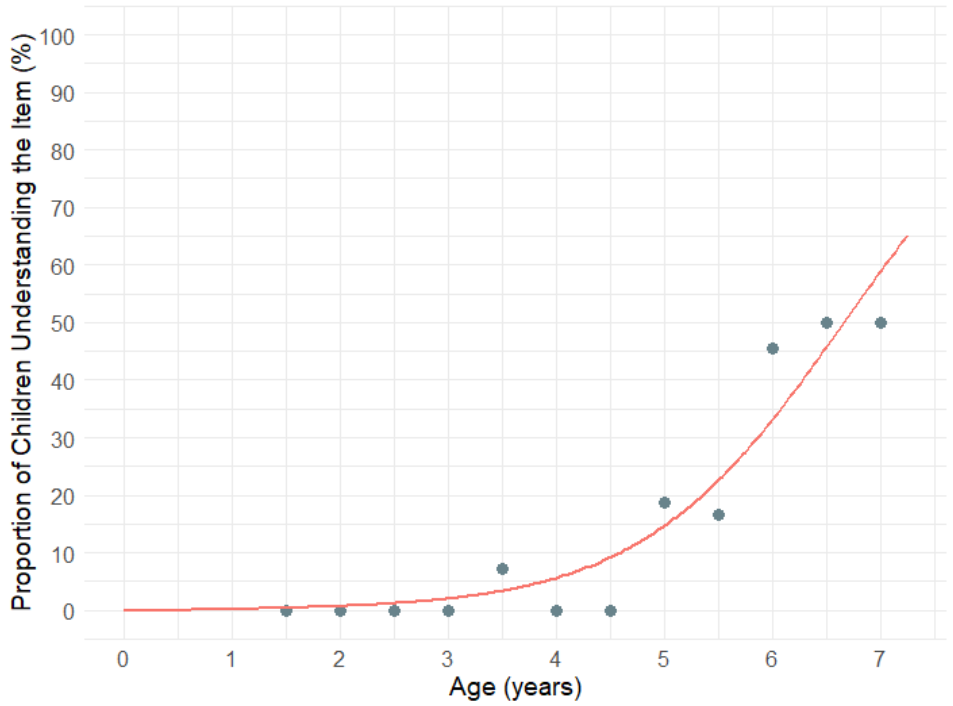

**Figure S41. Percentage of children whose parents responded ‘Very True’ to Item 7 in Table S3: Can subtract larger numbers: 15 – 7 = ?. 50% of children attained this item by 6.7 years. Markers represent the proportion of children who demonstrated understanding of the item, calculated in 0.5-year bins.**

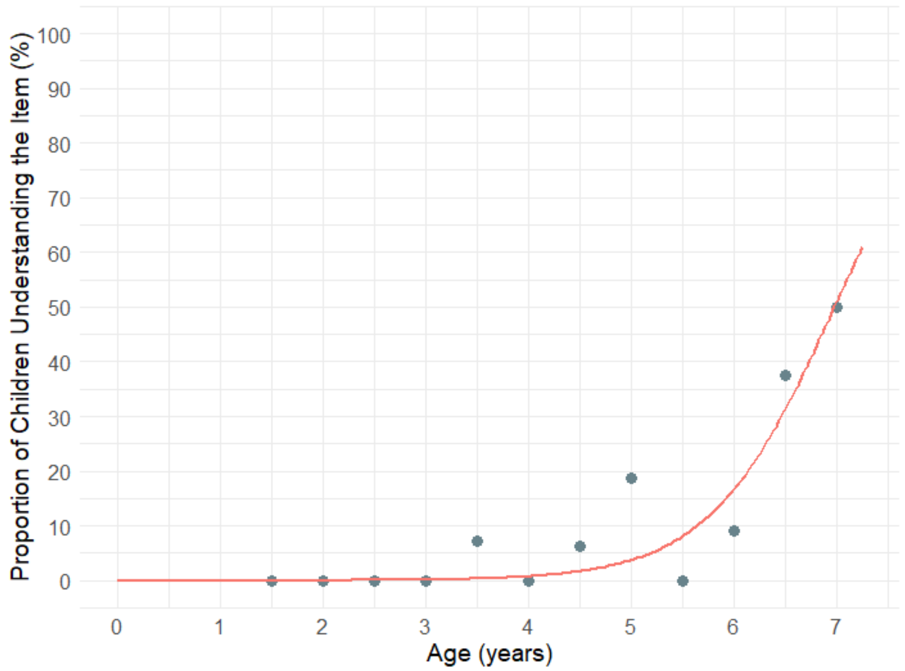

**Figure S42. Percentage of children whose parents responded 'Very True' to Item 8 in Table S3: Can perform simple multiplication: 2 × 2 = ?. 50% of children attained this item by 7.0 years. Markers represent the proportion of children who demonstrated understanding of the item, calculated in 0.5-year bins.**

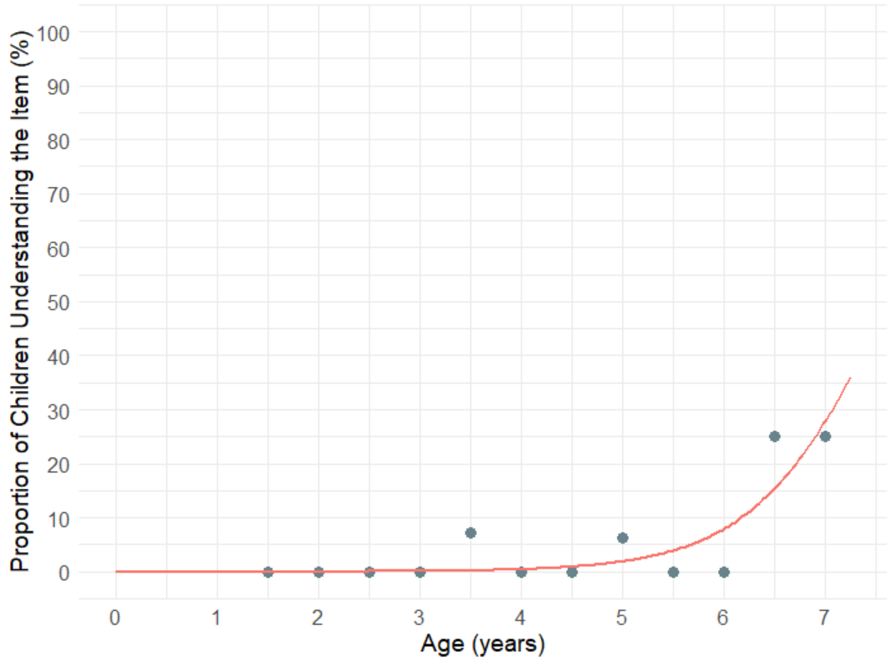

**Figure S43. Percentage of children whose parents responded 'Very True' to Item 9 in Table S3: Can multiply larger numbers: 6 × 7 = ?. 50% of children attained this item by 7.6 years. Markers represent the proportion of children who demonstrated understanding of the item, calculated in 0.5-year bins.**

**Table S4. Median age of attainment for each LPEC-extension item (years).**

| Item number | Median age of attainment |
|---|---|
| 1. Draws a VARIETY of RECOGNIZABLE images (objects, people, animals, etc.) | 3.9 |
| 2. Can draw a NOVEL image following YOUR description (e.g. a three-headed horse) | 4.6 |
| 3. Engages in a VARIETY of make-believe activities (such as: playing house, playing with toy soldiers, building forts and castles, etc.) | 2.5 |
| 4. Can perform simple arithmetic: 2 + 3 = ? | 4.7 |
| 5. Can add larger numbers: 7 + 6 = ? | 5.6 |
| 6. Can perform simple subtraction: 3 – 2 = ? | 5.4 |
| 7. Can subtract larger numbers: 15 – 7 = ? | 6.7 |
| 8. Can perform simple multiplication: 2 × 2 = ? | 7 |
| 9. Can multiply larger numbers: 6 × 7 = ? | 7.6 |

### Percentile curves for the 24-item parent-reported LPEC-extended

Given that Items 1 to 3 in Table S3 assess pre-syntactic abilities, while Items 4 to 9 extend the LPEC to cover post-syntactic abilities, it may be beneficial to report normative data for the 24-item LPEC-Extended Scale, which incorporates all 15 LPEC items from Table 1 along with all 9 items in Table S3 (LPEC-extension).

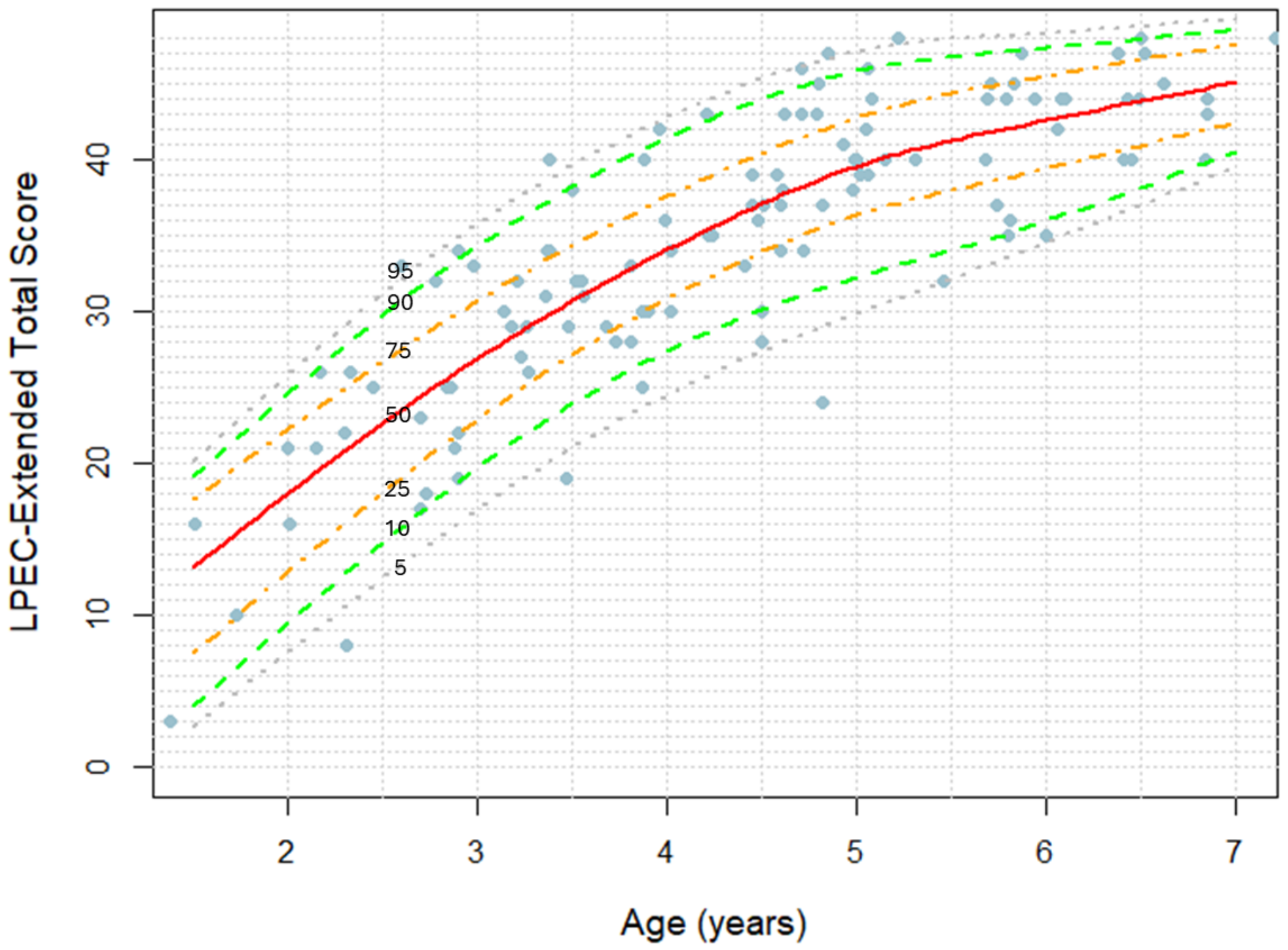

**Figure S44. Parent-reported 24-item LPEC-Extended score as a function of age. Markers represent LPEC-Extended scores in individual children.**

### Percentile curves for the 20-item parent-reported Mental Synthesis Evaluation Checklist (MSEC)

The LPEC-Extended Scale is the superset of the previously developed Mental Synthesis Evaluation Checklist (MSEC) [8–10], Table S5. In the MSEC, a lower score indicates better language comprehension ability: not true (2 points), somewhat true (1), very true (0). The subscale score ranges from 0 to 40 points. For the sake of continuity, we present MSEC normative data is Figure S42.

**Table S5: Mental Synthesis Evaluation Checklist (MSEC) [8].**

| |
|---|
| 1. Understands simple stories that are read aloud |
| 2. Understands elaborate fairy tales that are read aloud (i.e. stories describing FANTASY creatures) |
| 3. Draws a VARIETY of RECOGNIZABLE images (objects, people, animals, etc.) |
| 4. Can draw a NOVEL image following YOUR description (e.g. a three-headed horse) |
| 5. Engages in a VARIETY of make-believe activities (such as: playing house, playing with toy soldiers, building forts and castles, etc.) |
| 6. Understands some simple modifiers (i.e. green apple vs. red apple or big apple vs. small apple) |
| 7. Understands several modifiers in a sentence (i.e. small green apple) |
| 8. Understands size (can select the largest/smallest object out of a collection of objects) |
| 9. Understands possessive pronouns (i.e. your apple vs. her apple) |
| 10. Understands spatial prepositions (i.e. put the apple ON TOP of the box vs. INSIDE the box vs. BEHIND the box) |
| 11. Understands verb tenses (i.e. I will eat an apple vs. I ate an apple) |
| 12. Understands the change in meaning when the order of words is changed (i.e. understands the difference between 'a cat ate a mouse' vs. 'a mouse ate a cat') |
| 13. Understands NUMBERS (i.e. two apples vs. three apples) |
| 14. Can perform simple arithmetic: 2 + 3 = ? |
| 15. Can add larger numbers: 7 + 6 = ? |
| 16. Can perform simple subtraction: 3 – 2 = ? |
| 17. Can subtract larger numbers: 15 – 7 = ? |
| 18. Can perform simple multiplication: 2 × 2 = ? |
| 19. Can multiply larger numbers: 6 × 7 = ? |
| 20. Understands explanations about people, objects or situations beyond the immediate surroundings (e.g., “Mom is walking the dog,” “The snow has turned to water”) |

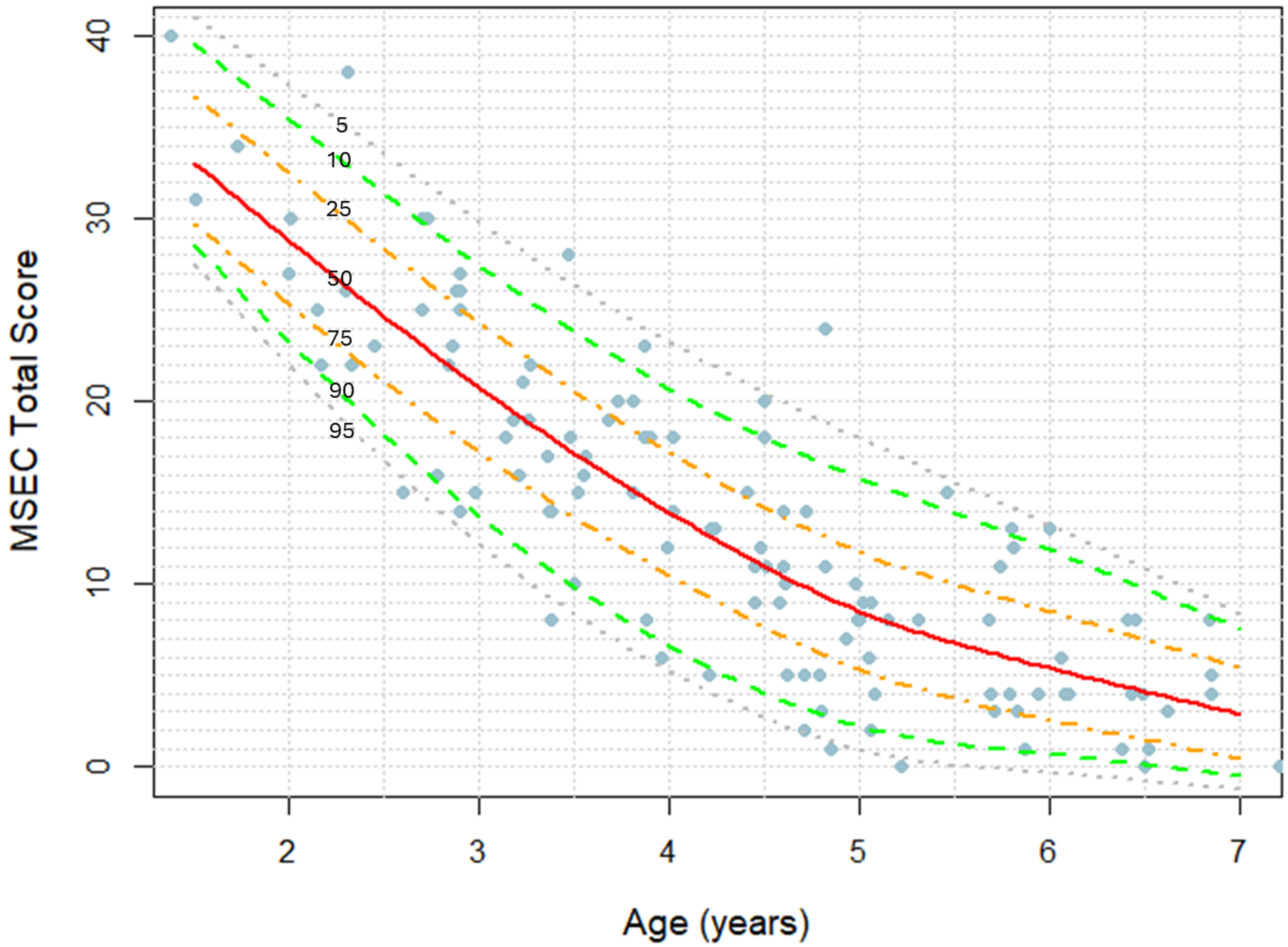

**Figure S45. Parent-reported 20-item MSEC score as a function of age. Markers represent LPEC-Extended scores in individual children. A lower score indicates better language comprehension ability.**

| LPA Score | LPEC Score | Language comprehension phenotype |
|---|---|---|
| 1 | 1 | Single word comprehension |
| 2 | 2 | Pre-command language comprehension |
| 3 - 5 | 3 - 5 | Command Language Comprehension Phenotype |
| 6 - 11 | 6 - 11 | Modifier Language Comprehension Phenotype |
| 12-15 | 12-15 | Syntactic Language Comprehension Phenotype |

**Table S6. Relationship between the LPA and LPEC scores and language comprehension phenotypes.**

### Participants diagnosed with Autism Spectrum Disorder (ASD)

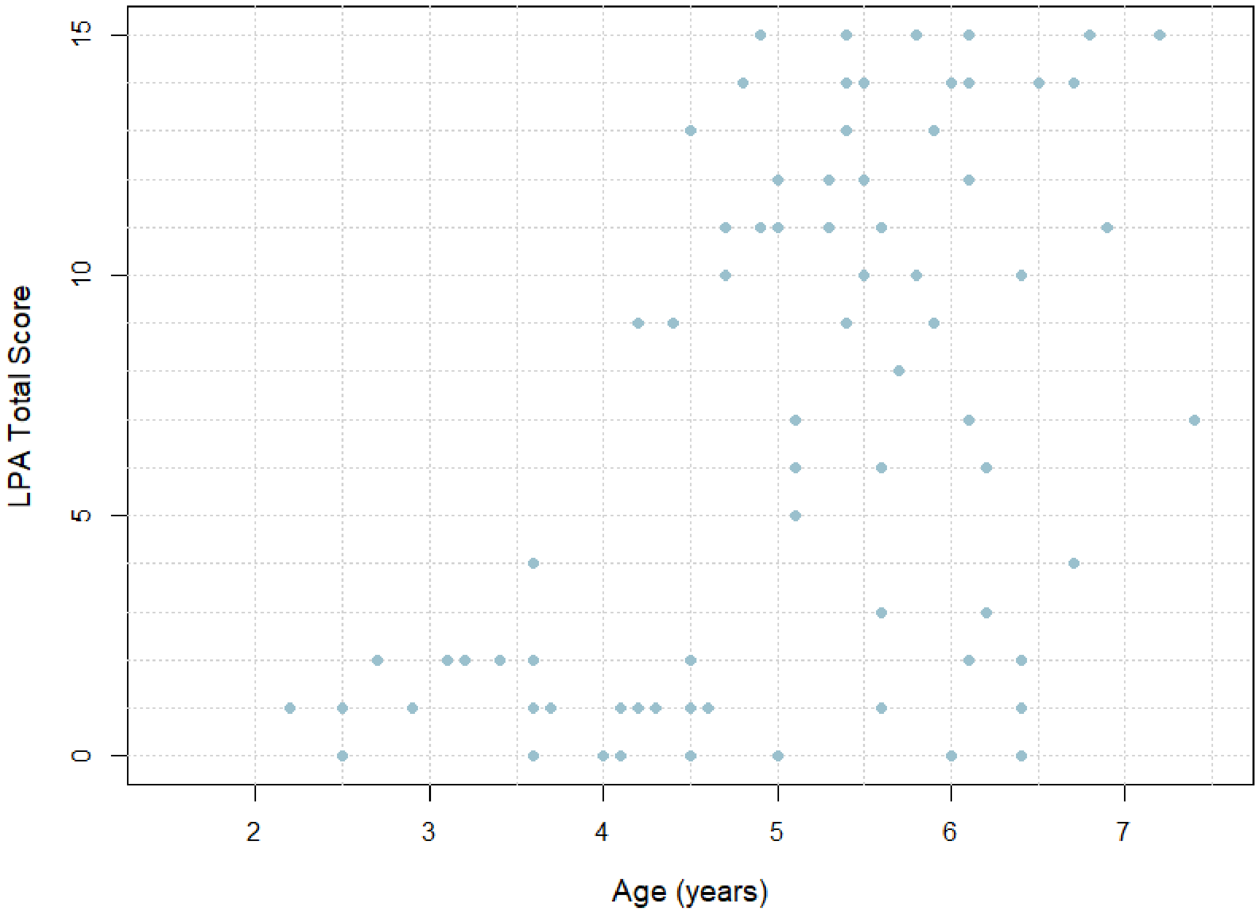

**Figure S46. Participants diagnosed with ASD: clinician-observed total LPA score as a function of age. Markers represent total LPA score in individual children.**

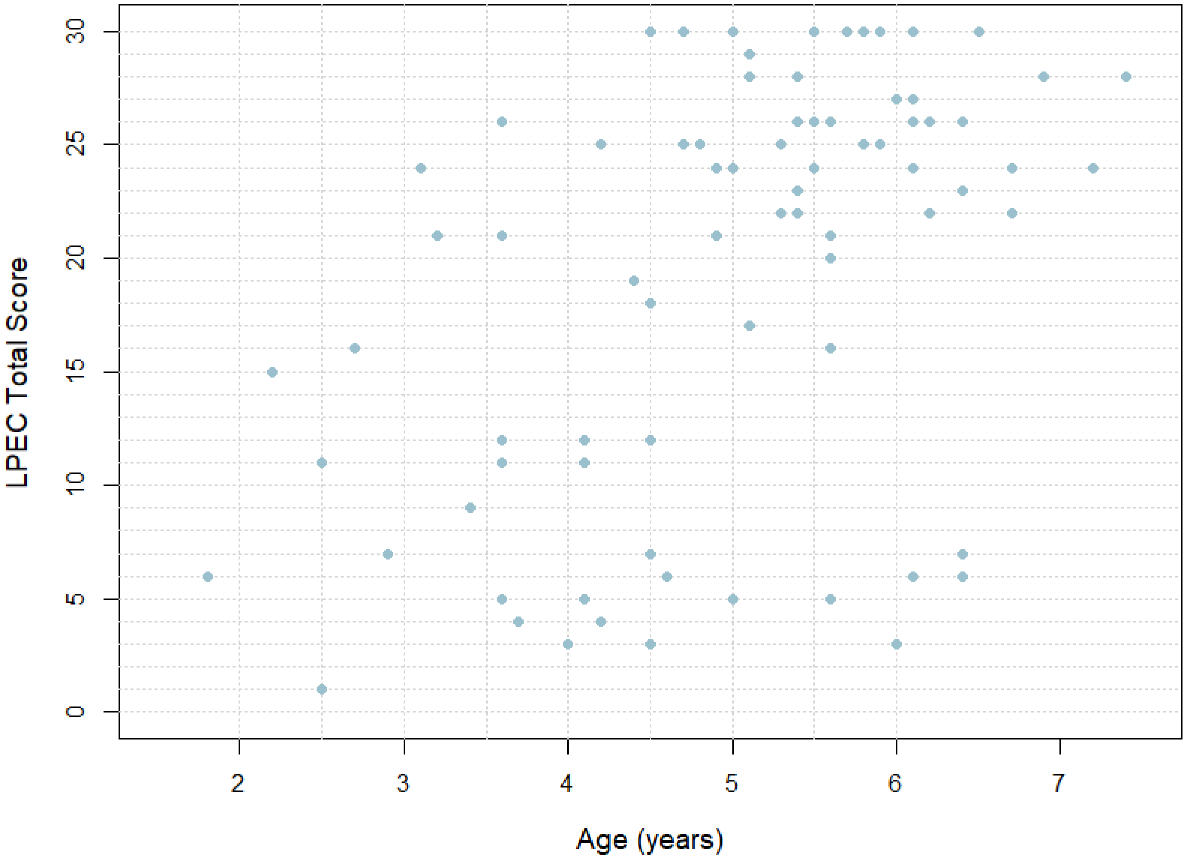

**Figure 47. Participants diagnosed with ASD: parent-reported total LPEC score as a function of age. Markers represent total LPEC score in individual children.**

**Table S7. Language comprehension phenotype in participants aged 5 years or older diagnosed with ASD (N=46). Compare to neurotypical participant: all neurotypical participants aged 5 year or more attained the Syntactic Phenotype.**

| Language Comprehension Phenotype | Determined by clinician-observed LPA (%) | Determined by parent-reported LPEC (%) |
|---|---|---|
| **Syntactic** | 39 | 48 |
| **Modifier** | 37 | 30 |
| **Command** | 13 | 9 |
| **Pre-command** | 11 | 13 |

**Table S8. Language comprehension phenotype in participants aged 4 years or older diagnosed with ASD (N=65). Compare to neurotypical participant: all neurotypical participants aged 4 year or more attained the Modifier Phenotype.**

| Language Comprehension Phenotype | Determined by clinician-observed LPA (%) | Determined by parent-reported LPEC (%) |
|---|---|---|
| **Modifier** | 66 | 68 |
| **Command** | 11 | 14 |
| **Pre-command** | 23 | 18 |

**Table S9. Language comprehension phenotype in participants aged 3 years or older diagnosed with ASD (N=74). Compare to neurotypical participant: all neurotypical participants aged 3 year or more attained the Command Phenotype.**

| Language Comprehension Phenotype | Determined by clinician-observed LPA (%) | Determined by parent-reported LPEC (%) |
|---|---|---|
| **Command** | 74 | 81 |
| **Pre-command** | 26 | 19 |

### References

1. Vyshedskiy, A., Venkatesh, R. & Khokhlovich, E. Representational drawing ability is associated with the syntactic language comprehension phenotype in autistic individuals. (2024) doi:10.1101/2024.07.26.24310995.
2. Vyshedskiy, A. & Khokhlovich, E. Pretend play predicts language development in young children with Autism Spectrum Disorder. *Int. J. Play* **12**, 403–419 (2023).
3. Kim, S. Pretend play and language development among preschool children: A meta-analysis. (2018).
4. Lillard, A. S., Pinkham, A. M. & Smith, E. Pretend play and cognitive development. *Wiley-Blackwell Handb. Child. Cogn. Dev.* **32**, 285 (2011).
5. Stagnitti, K. & Unsworth, C. The importance of pretend play in child development: An occupational therapy perspective. *Br. J. Occup. Ther.* **63**, 121–127 (2000).
6. Guerrero, D. Recursion in Language and Number: Is There a Relationship? (2020).
7. Guerrero, D. & Park, J. Arithmetic thinking as the basis of children's generative number concepts. *Dev. Rev.* **67**, 101062 (2023).
8. Braverman, J., Dunn, R. & Vyshedskiy, A. Development of the Mental Synthesis Evaluation Checklist (MSEC): A Parent-Report Tool for Mental Synthesis Ability Assessment in Children with Language Delay. *Children* **5**, 62 (2018).
9. Arnold, M. & Vyshedskiy, A. Combinatorial language parent-report score differs significantly between typically developing children and those with Autism Spectrum Disorders. *J. Autism Dev. Disord.* (2022) doi:/10.1007/s10803-022-05769-8.
10. Netson, R. *et al.* A Comparison of Parent Reports, the Mental Synthesis Evaluation Checklist (MSEC) and the Autism Treatment Evaluation Checklist (ATEC), with the Childhood Autism Rating Scale (CARS). *Pediatr. Rep.* **16**, 174–189 (2024).
